# Pan-UK Biobank GWAS improves discovery, analysis of genetic architecture, and resolution into ancestry-enriched effects

**DOI:** 10.1101/2024.03.13.24303864

**Authors:** Konrad J. Karczewski, Rahul Gupta, Masahiro Kanai, Wenhan Lu, Kristin Tsuo, Ying Wang, Raymond K. Walters, Patrick Turley, Shawneequa Callier, Nirav N. Shah, Nikolas Baya, Duncan S. Palmer, Jacqueline I. Goldstein, Gopal Sarma, Matthew Solomonson, Nathan Cheng, Sam Bryant, Claire Churchhouse, Caroline M. Cusick, Timothy Poterba, John Compitello, Daniel King, Wei Zhou, Cotton Seed, Hilary K. Finucane, Mark J. Daly, Benjamin M. Neale, Elizabeth G. Atkinson, Alicia R. Martin

## Abstract

Large biobanks, such as the UK Biobank (UKB), enable massive phenome by genome-wide association studies that elucidate genetic etiology of complex traits. However, individuals from diverse genetic ancestry groups are often excluded from association analyses due to concerns about population structure introducing false positive associations. Here, we generate mixed model associations and meta-analyses across genetic ancestry groups, inclusive of a larger fraction of the UKB than previous efforts, to produce freely-available summary statistics for 7,266 traits. We build a quality control and analysis framework informed by genetic architecture. Overall, we identify 14,676 significant loci (p < 5 x 10^-8^) in the meta-analysis that were not found in the EUR genetic ancestry group alone, including novel associations for example between *CAMK2D* and triglycerides. We also highlight associations from ancestry-enriched variation, including a known pleiotropic missense variant in *G6PD* associated with several biomarker traits. We release these results publicly alongside FAQs that describe caveats for interpretation of results, enhancing available resources for interpretation of risk variants across diverse populations.

## Introduction

Paired genetic and phenotypic data have grown explosively over the last decade, particularly with the maturity of massive global biobank efforts^1,2^. These data have led to the identification of over 275,000 associations between genetic loci and human traits and diseases to date^3^. However, genome-wide association studies (GWAS) tend to be vastly Eurocentric, limiting their generalizability to ancestrally and globally diverse populations^4,5^. Imbalanced data generation is the primary cause of this issue, but a secondary contributor is that GWAS tend to analyze the largest ancestry group in a dataset and exclude underrepresented groups to avoid potential false positives arising due to population stratification. Using the ancestrally diverse data already available is critical for several reasons, including increasing the applicability of genetic findings across populations, as well as increasing power for gene discovery due to increased genetic diversity.

Underrepresented populations also disproportionately contribute to genomic discoveries: for example, African ancestry and Hispanic/Latin American groups only comprise 2.4% and 1.3% of individuals represented in the GWAS catalog, respectively, but contribute 7% and 4.3% of associations overall^6^. In comparison, 78% of individuals have primarily European ancestry but contribute only 54% of associations.

### Box 1

**Genetic ancestry in the Pan-UKB**

At its core, a GWAS tests whether differences in allele frequencies correlate with trait variation. Differences in allele frequencies have arisen throughout human history, mainly driven by genetic drift (i.e., random changes in frequency over time). Failure to control for these differences (i.e. population stratification) can induce confounding, where mean differences in traits spuriously correlate with differences in allele frequencies, resulting in false associations. We evaluated two approaches to include as many individuals as possible into the GWAS: splitting based on genetic similarity (estimated by principal component analysis) then meta-analyzing, versus mega-analyzing all individuals in one model. We found that meta-analyzing genetic ancestry groups showed less evidence of cryptic stratification, reducing false positives and improving the robustness of associations. However, these groups are ultimately pragmatic, based on a variety of historical factors in how reference data were collected and analysis conventions were set; they do not imply the existence of discrete, biological ancestral populations (Discussion). We emphasize that genetic ancestry is a continuum with more genetic variation within these ancestry groups than differences between them.^7^ Interpreting genetic ancestry groups as monikers of race or ethnicity is unwarranted as it grossly oversimplifies the complexities of human demographic history and the diversity in ethnicity and identity present worldwide.^8^ The genetic ancestry groups used in this study were defined using a pair of well-known reference panels (HGDP+1kGP, see Supplementary Information). Throughout this manuscript, we refer to UK Biobank ancestry assignments using the ancestry labels provided by HGDP+1kGP: EUR (European), CSA (Central/South Asian), AFR (African), EAS (East Asian), MID (Middle Eastern), and AMR (Admixed American - an imprecise label introduced by the 1kGP to describe individuals with recent admixture from multiple continents including Amerindigenous ancestry), describing genetic similarity in accordance with recent reports that explored population descriptors in much greater detail^9,10^, e.g. HGDP+1kGP-AFR-like, which we shorten throughout the rest of the text to the three-letter population codes, e.g. AFR. More details on these and related points can be found in the Frequently Asked Questions (Supplementary Information).

Many trait- and disease-specific consortia have conducted multi-ancestry GWAS to increase sample sizes and investigate generalizability, which have yielded deep insights into biology^11^. For most traits and diseases, a large number of variants--each with a small effect size--contribute to phenotypic variation. As a general rule, causal variant effect sizes tend to be largely consistent across populations^12–14^, showing very low evidence of heterogeneity when accounting for differences in allele frequency and linkage disequilibrium (LD) across populations. Multi-ancestry studies, however, have highlighted some clear examples where ancestry-enriched variants (e.g. variants at 10-fold higher frequency in an ancestry group compared to others) provide power for genetic discovery that would not be readily identified only in European ancestry studies. Some examples include associations between *HNF1A* and type 2 diabetes identified in Latin American populations^15^; loss-of-function variant associations in *PCSK9* and low LDL cholesterol identified in African Americans^16^; associations between inflammatory bowel disease and variants enriched in East Asian ancestries^17^; associations between the Duffy-null allele and malaria identified in sub-Saharan African populations^18^; and associations between *APOL1* and resistance to trypanosomes but also chronic kidney disease in African and African diaspora populations^19–21^. Several of these associations have clinical implications that benefit individuals from all backgrounds, such as the *PCSK9* association which led to one of the first genetically informed therapies to prevent heart disease.

In addition to genomic discovery, multi-ancestry genetic analyses are also critical for resolving, interpreting, generalizing, and translating GWAS results. For example, diverse GWAS provide greater resolution into the identification of putative causal variation via fine-mapping due to: 1) a joint analysis of different patterns of LD across diverse populations^22,23^, and 2) larger sample sizes with more diverse ancestral recombination history^24–28^, improving identification of actionable targets using GWAS results. Another key application of GWAS is the construction of polygenic risk scores, with numerous potential clinical implications including disease risk stratification^29^. Genetic prediction accuracy closely reflects the composition of the study cohorts from which such models are derived, leading to the widely-replicated observation that Eurocentric discovery GWAS produce polygenic scores whose predictive powers are reduced by several fold in poorly represented genetic ancestry groups^5,30–33^. Multi-ancestry studies have already begun improving genetic prediction accuracy in underrepresented populations for some phenotypes^2,12,13,34,35^. Biobank-scale genetic correlation analyses and phenome-wide association studies are additional approaches that have aided our understanding of molecular and epidemiological relationships across a wide variety of traits; however, these have also tended to be Eurocentric in representation due to imbalances in GWAS^36–39^ and more diverse representation is needed particularly to disentangle ancestry-enriched genetic and environmental factors that contribute to disease risk.

The UK Biobank (UKB)^40,41^ is one of the most impactful biobanks to date, due to its large number of participants, depth and breadth of phenotyping, consistency in data generation, and uniquely open and straightforward access model; despite this, analyses have largely focused on only European ancestry participants. While most (95%) UKB participants fall into the EUR group (the genetic ancestry group with predominantly European ancestries), more than 20,000 participants have primarily non-European ancestries (**Box 1**). The UKB therefore provides opportunities to conduct some of the largest genetic studies to date of thousands of phenotypes in diverse continental ancestries.

Here, we describe the Pan-UK Biobank Project (https://pan.ukbb.broadinstitute.org/), a multi-ancestry genetic analysis of thousands of phenotypes. We extend previous phenome-wide association resources, adding 14,676 independent associations using meta-analysis of multiple ancestry groups rather than only the EUR subset. We highlight discoveries enabled by multi-ancestry analysis, including an association between *CAMK2D* and triglycerides. We also show how ancestry-enriched variation highlights interesting biology, including a pleiotropic association between *G6PD* and a number of biomarker traits, primarily accessible in the AFR genetic ancestry group. We describe and release pipelines for a robust analytic framework to facilitate future multi-phenotype multi-ancestry genetic analyses to improve gene discovery.

## Results

### A resource of multi-ancestry association results from 7,266 phenotypes in the UKB

To maximize genomic discovery in the UKB, we performed a multi-ancestry analysis of 7,266 phenotypes, with analysis of up to 441,331 total individuals from up to 6 genetic ancestry groups followed by meta-analysis. We assigned each individual to a genetic ancestry group by conducting principal components analysis (PCA) on a diverse reference panel consisting of the Human Genome Diversity Panel (HGDP) and 1000 Genomes Project genotype data^42,43^ (**Supplementary Table 1**), then projected UKB individuals into this space using their genotype data using a random forest (probability > 0.5) to partition the dataset into six genetic ancestry groups (**Extended Data Fig. 1, Supplementary Figs. 1-13, Supplementary Tables 2-5**). After initial assignments, to further reduce stratification, we removed ancestry outliers based on multidimensional centroid distances from average PC values (**Supplementary Information**). The ancestry groups follow broad expected trends based on self-reported ethnicity (as defined by the UK), continental birthplaces, and country of birth although there is notably still considerable within-group diversity (**Supplementary Tables 6-7, Supplementary Fig. 14**); while principle components correlate with these concepts, correlations deviating from 1 reflect the fact that genetic ancestry is a distinct concept from these other identity- and geography-based descriptors (see **Box 1** and **Supplementary Information**, FAQ). For example, a person assigned to some genetic ancestry group may or may not report having a corresponding ethnic identity^44–46^. From these data, we performed sample and variant quality control (QC) to remove sample outliers and low-quality and ultra-rare variants (**Methods**). This resulted in 20,800 individuals assigned to non-EUR ancestry groups with 42,535 remaining unclassified, most of whom were preliminarily assigned with similarity to EUR and then subsequently removed as outliers (**Supplementary Tables 2 and 4**).

We used a two-step approach for genetic association testing, first performing GWAS within each genetic ancestry group for a given trait using a generalized mixed model approach (SAIGE^47^), and then performing fixed-effect inverse-variance weighted meta-analysis across all within-ancestry GWAS performed for that trait. Consistent with previous studies^48^, we observed similar results between mega- and meta-analyses, with the meta-analysis approach reducing stratification and type 1 error rate compared to an alternative single-step “mega-analysis” approach that included all individuals in a single mixed model (**Extended Data Table 1**). Specifically, we observed genomic control statistics closer to 1 using our approach, as well as comparable and in some cases improved statistical power with discovery of additional genome-wide significant loci using our approach.

Using this two-step approach, we performed association testing across 10-23 million SNPs (**Supplementary Table 8**) for all ancestry-trait pairs for quantitative phenotypes, and all binary traits with at least 50 cases observed in an ancestry group (except EUR, 100 cases, given the larger sample size; **Fig. 1a**; **Supplementary Fig. 15**). Altogether, this resulted in an analysis of 16,528 ancestry-trait pairs across 7,266 traits, of which 922 were run in all six genetic ancestry groups (**Fig. 1b**). These traits include thousands of newly-analyzed phenotypes, including aggregate disease combinations (i.e., phecodes^37^), prescription drug status, and continuous updates to the COVID-19 phenotypes^49^. We developed a summary statistics QC protocol to remove low-confidence variants and associations (**Supplementary** Figs. 16-19).

**Figure 1.**
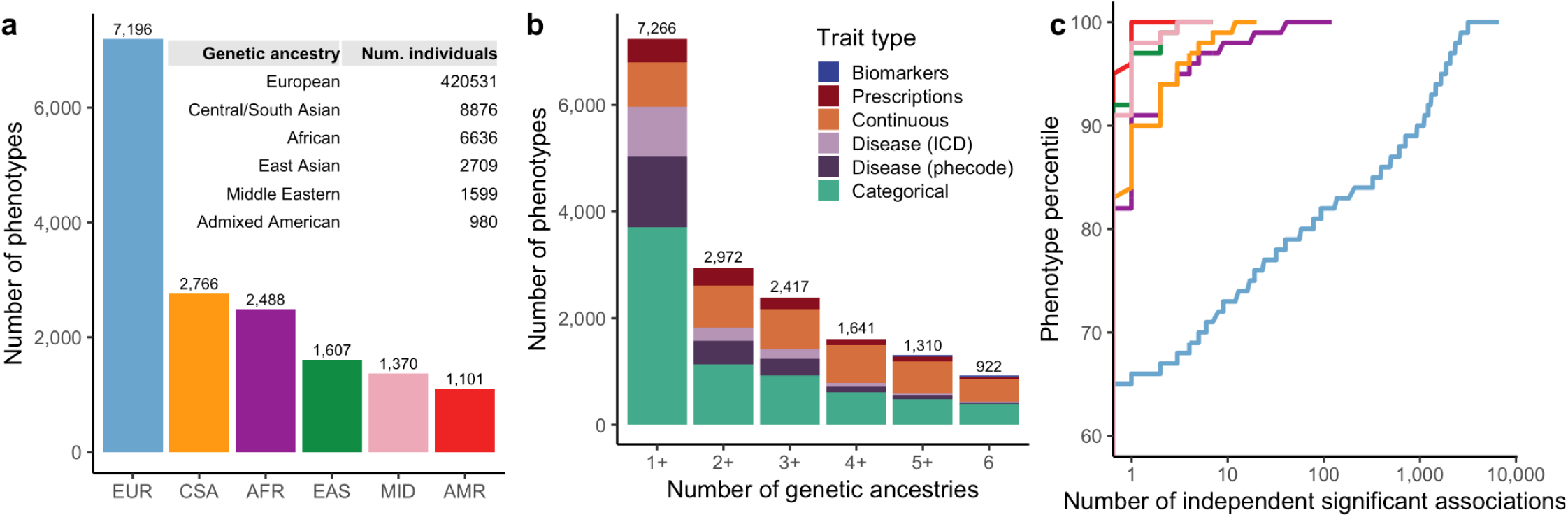
Pan-UK Biobank GWAS resource facilitates multi-ancestry multi-trait analyses. **a-b**, The number of phenotypes with GWAS computed across: **a**, genetic ancestry groups and **b**, number of genetic ancestry groups stratified by trait type; within each bar, trait types are ordered by total number of traits. A full list of phenotypes can be found in **Supplementary Dataset 2**. **c**, A cumulative distribution function showing the number of independent genome-wide significant (p < 5 x 10^-8^) associations across all phenotypes analyzed within each genetic ancestry group. Independence was defined using clumping in plink^50^, including an *r*^2^ threshold of 0.1 for ancestry-matched reference panels (Supplementary Information). Colors are consistent in **a** and **c**.

To avoid double counting variants in LD, we determined a set of LD-independent loci within each ancestry-trait pair using in-sample reference panels (**Supplementary Figure 20**). We extended this to our meta-analysis by constructing reference panels of 5,000 individuals matched by genetic ancestry proportions. For 922 traits for which association analysis was performed for all six ancestry groups, we discovered a mean of 2.26 independent loci in non-EUR genetic ancestry groups, demonstrating that we have sufficient power for novel discovery in these understudied groups for which GWAS have not previously been run at scale in the UKB. Owing to the substantially larger sample size in EUR, we find more genome-wide significant associations per phenotype in EUR: 25% of phenotypes have >18 associated loci in EUR while fewer than 10% of phenotypes have 3 or more associated loci across all non-EUR populations (**Fig. 1c**). Despite the lower sample size but consistent with previous GWAS catalog summaries^6^, we find a higher mean number of significant regionally-independent associations in AFR compared to CSA, potentially suggesting increased power from higher heterozygosity (**Extended Data Table 2**).

### A framework for identifying high-quality phenotypes in diverse ancestries with massively imbalanced sample sizes

Previous phenome-wide studies of the UK Biobank have performed association testing of thousands of traits; however, these have used linear models^51^ and/or only analyzed a single ancestry group^47,51^. In our analysis of data from multiple ancestry groups, we identified extensive challenges due to extreme imbalances of sample sizes, leading to unreliability of some association tests. Here, we propose a framework for testing the reliability of GWAS of multiple ancestries.

Specifically, we summarized properties of genotype-phenotype associations, including genomic control (λ_GC_), heritability, and residual population stratification. We estimated SNP-heritability (*h*^2^*_SNP_*) for each ancestry-trait pair through several strategies. For a set of pilot phenotypes in EUR (**Supplementary Table 9**), we confirmed a high concordance for two methods (**Extended Data Fig. 2**; S-LDSC^52,53^ and RHE-mc^54^), as well as between our results and previously reported results (**Supplementary** Figs. 21-24). To balance computational considerations with power gains, we used S-LDSC for all traits in EUR, and RHE-mc for all other genetic ancestry groups (**Extended Data Fig. 2b; Supplementary Information**). Across groups, the number of traits with significant heritability (*z* ≥ 4) correlates with sample size of genetic ancestry group (**Extended Data Fig. 3a**; **Supplementary Fig. 25**). We identified traits with significant out-of-bounds (i.e. outside 0-1) heritability point estimates, deflated λ_GC_ estimates, and/or elevated S-LDSC ratio statistics, particularly in traits that are prone to population stratification (e.g. country of birth, dietary preferences; **Supplementary Fig. 26**).

To increase confidence in analyses across traits and ancestries, we devised a quality control strategy to systematically flag traits with potentially problematic GWAS results while retaining GWAS of heritable traits passing QC in two or more populations (**Fig. 2a, Supplementary** Figs. 27-29, and Supplementary Information). Overall, we pruned 16,528 ancestry-trait pairs with available GWAS to 1,091 ancestry-trait pairs that passed all filters spanning 452 traits (**Fig. 2a**). Of the ancestry-trait GWAS pairs that passed, the majority were shared between the two largest ancestry groups (EUR and CSA), with 147 phenotypes found in three or more genetic ancestry groups (**Supplementary Fig. 29**). As many phenotypes in the biobank are correlated with each other, we pruned to a maximal set of independent phenotypes with pairwise r^2^ < 0.1, resulting in a set of 151 phenotypes (**Supplementary Fig. 30**), and computed polygenicity estimates for all high quality traits (**Supplementary Fig. 31**). We note that these genome-wide summaries aid in prioritizing phenotypes with broad heritable components, although the failure of a phenotype in this framework does not necessarily preclude true individual SNP-level associations.

**Figure 2.**
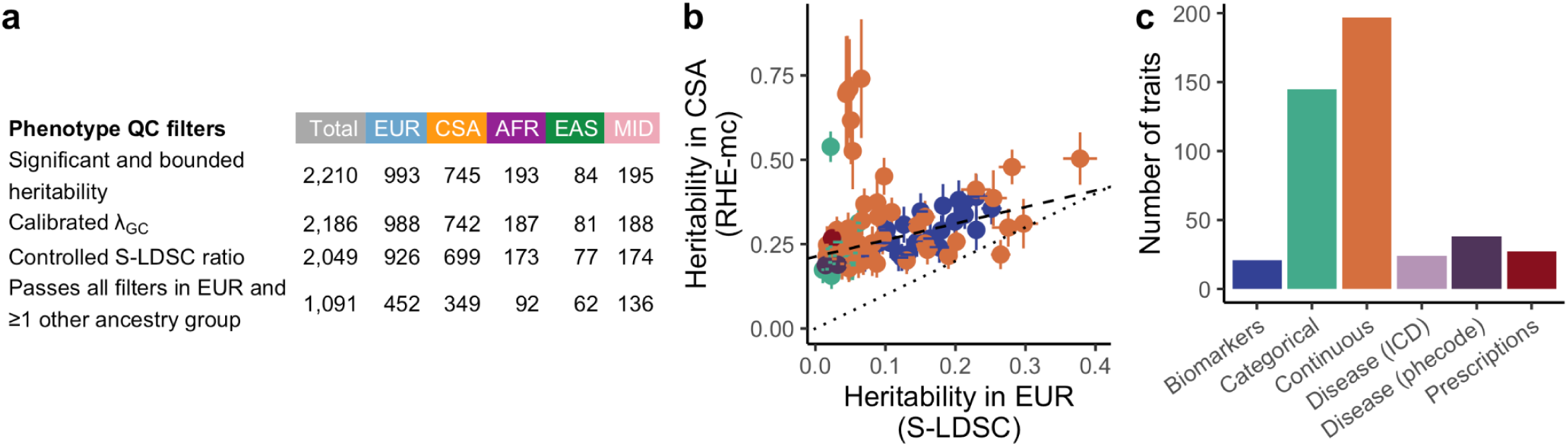
Heritability informs robustness of GWAS across ancestry-trait pairs. **a**, To balance large differences in sample size across genetic ancestry groups, we developed a stepwise series of phenotype QC filters applied based on heritability estimates, genomic control (λ_GC_), evidence of residual stratification (S-LDSC ratio), and high-quality data in multiple ancestry groups (see also **Supplementary** Figs. 25-27 for more detail). **b**, Comparison of heritability estimates across EUR and CSA genetic ancestry groups. Note that two different heritability estimation methods are used to balance computational efficiency (S-LDSC in EUR) versus precision in small sample sizes (RHE-mc in CSA). Binary phenotype heritability estimates are reported on the observed scale due to highly variable prevalences and liability scaling at smaller sample sizes. Error bars indicate standard errors. The dotted line shows y=x, while the dashed line is a fitted York regression (slope = 0.49, intercept = 0.21, p = 5 x 10^-12^). A version with less filtering is shown in Extended Data Fig. 2c. **c**, The number of traits passing final QC filters by trait type. Colors are consistent in **b, c**.

Among high-quality GWAS results that passed our broad-scale QC of GWAS results, we identified consistency in the relative magnitude of heritability estimates among populations. For example, 113 independent phenotypes passed QC in both EUR and CSA genetic ancestry groups, and the heritability estimates for these traits had a significant positive correlation (York regression p = 5 x 10^-12^; **Fig. 2b, Extended Data Fig. 2c**); the heritability estimates in this set were systematically higher in the CSA, likely reflecting a combination of winner’s curse from selection of phenotypes with significant estimates in CSA and of residual population stratification. Biomarkers and continuous phenotypes tended to have the highest heritability estimates (EUR average h^2^ = 0.19 and 0.16), whereas disease and prescription phenotypes tended to have the lowest heritability estimates (EUR average h^2^ = 0.021 and 0.016, **Extended Data Fig. 3b**).

### Genomic discoveries powered by Pan-UK Biobank analysis

We next investigated the extent to which broader inclusion of ancestrally diverse participants identifies specific biological signals through meta-analysis, compared to EUR-only analysis. In particular, for 452 high-quality phenotypes, we compared p-values for the multi-ancestry meta-analysis to those from the EUR GWAS, which were overall highly correlated (**Fig. 3a**; **Supplementary Fig. 32**; r^2^ = 0.999; p < 10^-100^). We identified 237,360 significant (p < 5 x 10^-8^), LD-independent associations in the largest meta-analysis available across 431 phenotypes, with LD defined as above (and **Supplementary Figure 20**). Of these, 14,676 (6.2%) were not significant in EUR, representing new biology discovered solely by analyzing already-available data from diverse populations.

**Figure 3.**
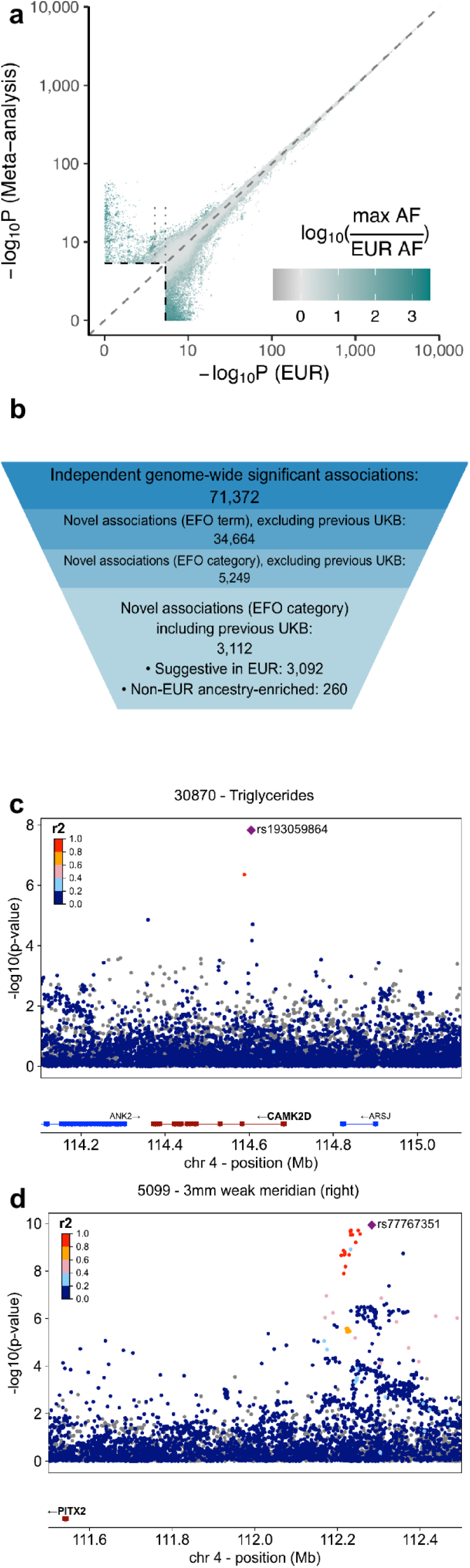
Biobank-wide analysis improves genetic discovery. **a**, Comparison of GWAS significance in EUR GWAS only versus meta-analysis across ancestries. Variants with p < 10^-10^ in either analysis are shown. Colors highlight that variants with higher non-EUR allele frequencies (teal) are more likely to be identified in the meta-analysis. Dashed gray line shows y=x, and the scale is logarithmic for p from 1 to 10^-10^, and log-log for p < 10^-10^. Dotted lines indicate variants suggestive in EUR (5 x 10^-8^ < p < 10^-6)^, but significant in meta-analysis (p < 5 x 10^-8^; see Fig. 3b). **b,** Summary of regionally-independent genome-wide significant associations. The total number, number of novel associations (at the EFO term and category level), excluding and not excluding previous UK Biobank multi-phenotype analyses^47,51^. The final category shows the number of associations that are suggestive in EUR alone (p < 10^-6^), and where the frequency of the variant is enriched in a non-EUR genetic ancestry group by at least 10-fold (247 that were both suggestive and enriched). **c-d**, Locuszoom plots of (**c**) *CAMK2D* (rs193059864) and triglycerides (p = 1.5 x 10^-8^; N = 416,764; allele frequency in AFR = 0.016, EUR = 1.4 x 10^-4^) and (**d**) *PITX2* (rs77767351) of keratometry (3mm weak meridian - right; p = 1.2 x 10^-10^; N = 89,664), with lead variants indicated by the purple diamond and LD (r^2^) for neighboring variants derived from a weighted reference panel (see Supplementary Information). Both lead variants (**c-d**) have info score > 0.9 and heterogeneity p-value > 0.1.

In order to compare to prior trait-specific and systematic analyses in the UK Biobank^47,51^ and other datasets, we created a framework to quantify associations added by the Pan-UK Biobank analysis compared to existing associations in Open Targets Genetics^55^. Two challenges with phenome-wide comparisons is the pervasiveness of pleiotropy coupled with mapping phenotypes across GWAS, which may be coded slightly differently and have subtle differences in phenotype definitions^56^. Through semi-manual curation, we mapped 3,047 (42%) of our traits to terms in the Experimental Factor Ontology (EFO), of which 2,566 matched a study in the GWAS Catalog; our ability to cross-reference phenotypes varied by trait type (**Supplementary Table 10**). Furthermore, as LD-independent associations are challenging (e.g. even r^2^ < 0.1 can result in spurious “independent” signals when in LD with extremely significant signals with □^2^ > 300), we computed distance-based independent associations in order to assess novelty conservatively, and compared to known associations accordingly.

Specifically, we defined a variant as novel only if no known associations for the same EFO broad category are present within 1 Mb. We identified 71,372 regionally-independent associations that mapped to an EFO category, of which 68,260 (96%) were previously reported for the same EFO category. 3,112 (4%) were not previously identified (**Fig. 3b**), with this rate varying by EFO category (**Extended Data Fig. 4**). The X chromosome contributes an outsized proportion of this novelty, with 573 of 2,448 (23%) associations not previously reported for the same EFO category. This disproportionate contribution is likely due to exclusion of the X chromosome in many previous GWAS^57^.

Newly-significant associations arise due to a combination of factors: bolstered associations that crossed the genome-wide significance threshold with mixed models from previously sub-threshold associations in standard regression, increased sample size of EUR participants, and inclusion of participants with non-EUR ancestries that either added support or contributed outsized significance due to ancestry-enriched variation. Of the 3,112 novel meta-analysis associations, 3,092 show suggestive signals (e.g. 5 x 10^-8^ < p < 10^-6^) in EUR-only analyses where the multi-ancestry meta-analysis results in genome-wide significance (p < 5 x 10^-8^). Allele frequencies were enriched by at least 10-fold in at least one genetic ancestry group over EUR in at least one genetic ancestry group for 260 associations (247 both suggestive in EUR and enriched outside EUR; **Fig. 3b**). For instance, we find a significant association between triglycerides and *CAMK2D* (meta-analysis p = 1.5 x 10^-8^; EUR p = 0.0017; **Fig. 3c**), at a variant (rs193059864) that is 114-fold enriched in AFR (AFR frequency 1.6%, EUR frequency 1.4 x 10^-4^). Another variant in this gene has recently been implicated in heart failure^58^; however, rs193059864 is in low LD with the variant identified in this study (rs17620390; r^2^ = 0.001), indicating that these likely represent independent associations.

We investigated broad patterns of gene function closest to each association to assess biological relevance. We find that 66% (124/187) of haploinsufficient genes are near a novel significant association (**Extended Data Fig. 5a**; all hits shown in **Supplementary Fig. 33**), compared to 34% of all genes (5,969/17,428). The associations near haploinsufficient genes often correspond to broadly similar phenotype categories as the OMIM annotation of the gene. For instance, we find associations between SNP rs77767351 (near *PITX2*) and several keratometry measurements, including 3mm weak meridian-right (meta-analysis p-value = 1.2 x 10^-10^; **Fig. 3d**). Previous studies have identified a crucial role for *PITX2* in embryonic development and tissue formation^59^ and implicated mutations in this gene with rare Mendelian eye-related diseases, such as Axenfeld-Rieger syndrome^60,61^. To our knowledge, this SNP has not been significantly associated in any GWAS, and indicates a potential allelic series, in which the intermediate eye phenotype associated with the common variant can provide context for the molecular mechanisms underlying the dominant condition caused by high-impact variants in *PITX2*.

### Inclusion of multiple genetic ancestries improves genetic discovery

To further explore potential reasons for associations only significant in the meta-analysis, we compared effect sizes to allele frequencies by ancestry. For quantitative traits where effect sizes are directly comparable, we broadly observed the characteristic inverse relationship between minor allele frequency (MAF) and effect size, in which rarer variants tend to have larger effect sizes than common variants^62,63^ (**Fig. 4a**). We identified genome-wide significant associations through meta-analysis and not in the EUR GWAS in some cases due to higher ancestry-specific MAFs, which rendered some variants with larger effect sizes accessible to discovery despite the smaller sample size (**Fig. 4b**), although winner’s curse may inflate some effect size estimates^64^.

**Figure 4.**
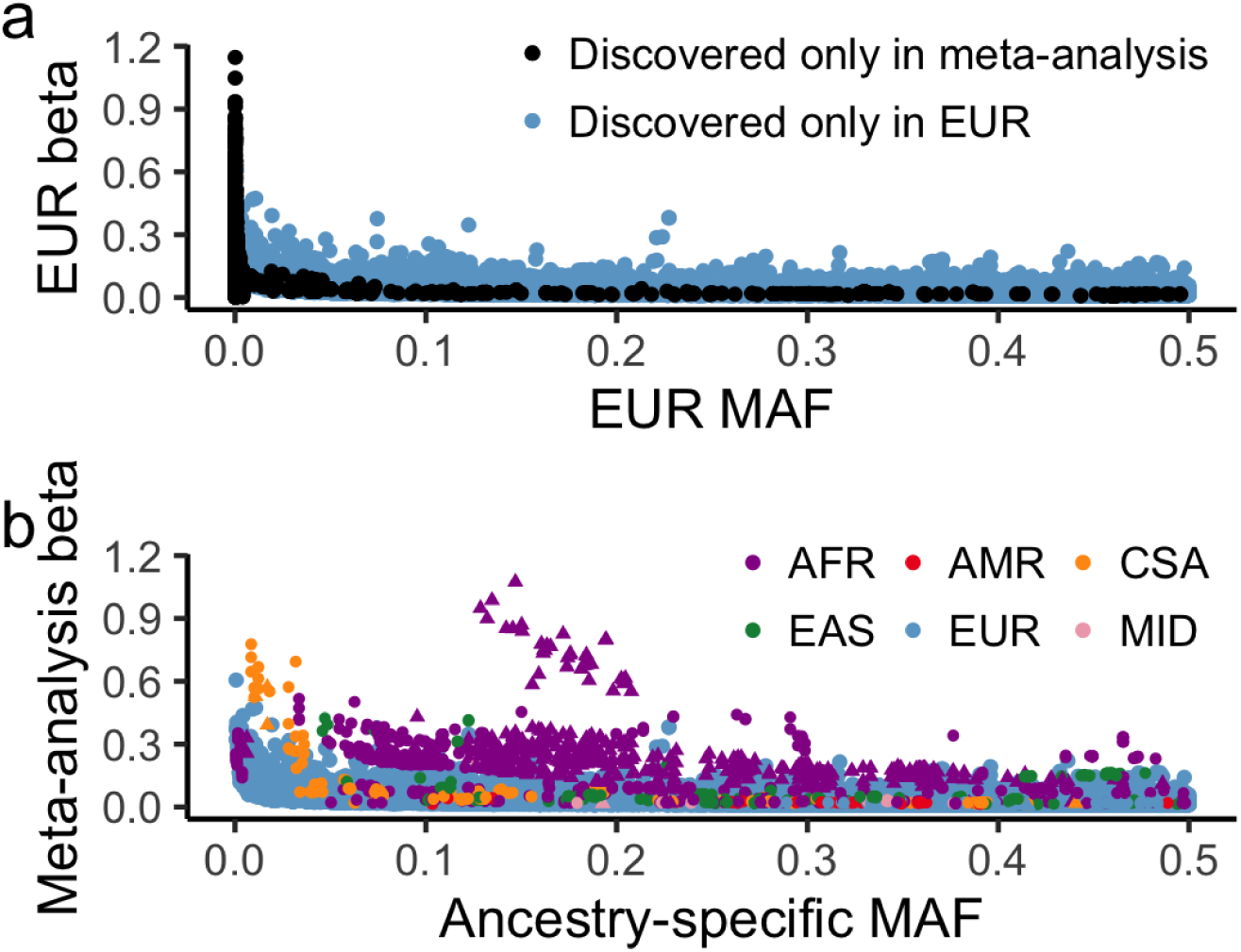
Differences in allele frequencies across ancestries yield novel genetic discoveries. **a**, Significantly associated variants for a set of 140 quantitative traits identified in the EUR genetic ancestry group (blue) versus those discovered only in the meta-analysis (black), with allele frequencies and effect sizes in EUR shown. **b**, The same significantly associated variants as shown in **a**, but with ancestry-specific frequencies and effect sizes estimated from the multi-ancestry meta-analysis. Associations on the X chromosome (e.g. *G6PD*) are denoted with triangles. Contrasting **a-b** highlights the importance of higher allele frequencies in underrepresented ancestry groups for empowering associations.

We thus investigated the most extreme differences (i.e. associations significant in the meta-analysis, but not in the EUR GWAS, i.e. those in the upper left quadrant of **Fig. 3a**). We find that these associations were more likely to be found at high-quality variants (**Supplementary Information**) and were 6-fold enriched for variants more common in AFR (4-fold in any non-EUR ancestry; **Extended Data Fig. 6**), indicating that these associations are likely enriched for universally tagging or putatively causal variants in multiple populations. The most extreme differences in p-values stemmed from extreme frequency differences, such as between variants in/near *G6PD* and a number of traits (**Fig. 5**), where a higher frequency in AFR increased power for association. However, perhaps paradoxically, variants with p-values more significant in EUR-only GWAS compared to meta-analysis were also 2.4-fold enriched for those with higher AFR frequencies (**Extended Data Fig. 6**). These variants are most likely not causal, but tagging variants with differential LD patterns across ancestries leading to apparent heterogeneity in meta-analysis. For instance, a variant in LD with a causal variant in EUR, but only partial LD in AFR with increased frequency will be identified as heterogeneous with little or no effect in AFR. Indeed, heterogeneity (Cochran’s Q p < 0.01) was 2.5-fold enriched for variants with reduced significance in the meta-analysis compared to the EUR GWAS alone (**Extended Data Fig. 6; Supplementary Fig. 32**).

**Figure 5.**
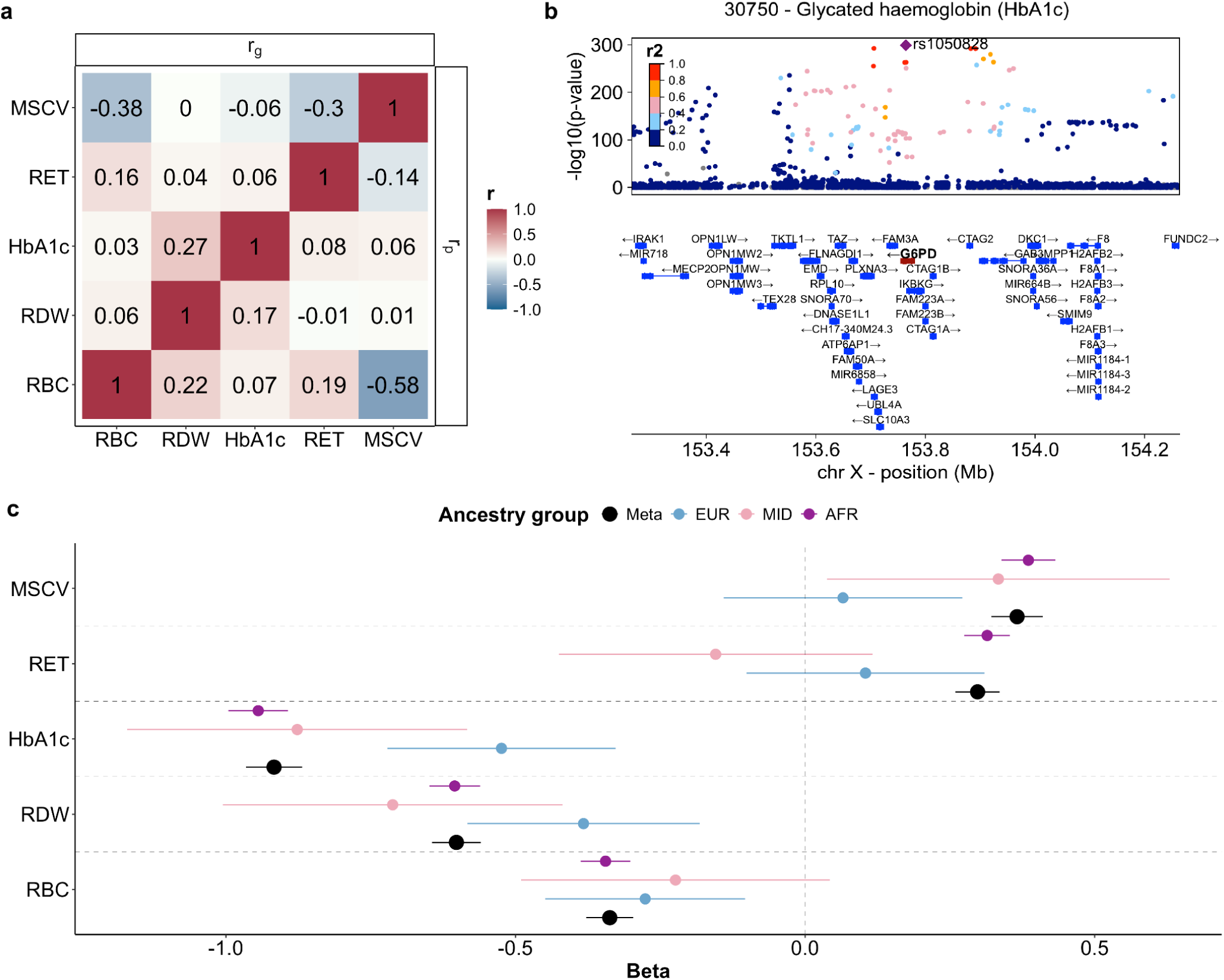
Meta-analysis identifies pleiotropic signals from non-European populations. **a,** Correlation matrix of the top five phenotypes significantly (meta-analysis p < 5 x 10^-8^) associated with rs1050828 (chrX:153764217), a missense variant in *G6PD* (all p-values are shown in Supplementary Table 11). The upper and lower triangles of the matrix represent the genetic and phenotypic correlations, respectively. (RET: High light scatter reticulocyte count; RDW: Red blood cell (erythrocyte) distribution width; RBC: Red blood cell (erythrocyte) count; MSCV: Mean sphered cell volume; HbA1c: Glycated hemoglobin) **b**, Locuszoom plot of a 1Mb region around the lead SNP rs1050828 (purple diamond) for the meta-analysis result of glycated hemoglobin (p = 1.1 x 10^-299^; N = 408,539), as in Fig. 3c**-d**. **c**, Forest plot showing association beta for each phenotype for rs1050828, including a meta-analysis across all available ancestry groups (full results shown in **Supplementary Fig. 34**). rs1050828 was low-frequency in CSA and EAS and thus, association statistics were not computed. Error bars correspond to 95% confidence intervals.

### Refining biological signals through ancestry enriched variation

Among the most extreme differences we identified between the meta-analysis and EUR-only results was a missense variant (rs1050828) in *G6PD*, which had significant associations with five phenotypes in the AFR group (allele frequency = 16%; **Fig. 5a**) but not in EUR (allele frequency = 1.5 x 10^-4^), including glycated hemoglobin (HbA1c; **Fig. 5b**), high light scatter reticulocyte count, red blood cell count, red blood cell distribution width, and mean sphered cell volume (**Fig. 5c, Supplementary Fig. 34, Supplementary Table 11**). Our results replicate previous associations between variants in/near *G6PD* and HbA1c^65^, and further validate the pleiotropic effect of this missense variant. We performed ancestry-specific fine-mapping of these signals (**Extended Data Fig. 7a**), which confirm the likely functional nature of this variant; however, fine-mapping of meta-analyzed summary statistics presented a significant challenge (**Extended Data Fig. 7b**), particularly due to highly imbalanced sample sizes.

To further explore the influence of ancestral haplotypes on trait associations, we performed a pilot study running *Tractor*^66^ for a subset of traits on a similar set of individuals to the AFR SAIGE tests. We find further improved false positive control and accurate group-level effect size estimation compared to SAIGE (**Extended Data Fig. 8; Supplementary Dataset 7; Supplementary Information**), highlighting the utility of further refining ancestry to more robustly evaluate genetic associations with traits

## Discussion

We present a multi-ancestry genetic study and resource that extends previous analyses focused only on EUR participants to a wider set of UKB participants from understudied genetic ancestry groups. Here, we build a new framework, including pipelines and best practices for multi-phenotype multi-ancestry analysis, that we recommend be adopted by groups performing similar analyses for the UK Biobank and other diverse biobanks, such as the *All of Us* project. Our quality control framework provides a blueprint to identify phenotypes with robust GWAS, particularly when sample sizes are imbalanced and/or with stratification. Our results show that diverse analysis is critical for maximizing biological discovery even with imbalanced sample sizes, and that full data release is critical to broad comparisons across datasets. We provide summary statistics for 16,528 GWAS in scalable and per-phenotype form, as well as reference data (including LD scores/matrices and sample metadata returned to the UK Biobank) to reduce the barrier for future multi-ancestry analyses of these existing and new phenotypes, respectively. Indeed, this work contributed continuously updated summary statistics to the COVID-19 host genetics initiative^49^, as well as broader efforts such as the Global Biobank Meta-analysis Initiative^67^.

In this work, we highlight challenges in performing multi-phenotype analyses of thousands of phenotypes, particularly around quantifying novelty and fine-mapping in the presence of imbalanced sample sizes. We identify 3,112 genome-wide significant associations that were not previously found in Open Targets Genetics^55^. These discoveries arise from ancestry-enriched variants (e.g. *CAMK2D* rs193059864 with frequency enriched in AFR) as well as previously sub-significant signals (5 x 10^-8^ < p < 10^-6^; e.g. rs77767351 in *PITX2*), which are significantly associated here due to a combination of larger sample sizes in the European ancestry group, inclusion of participants with primarily non-European ancestries, and use of more advanced mixed models that increase statistical power for discovery. Some associations demonstrate allelic series, in which variants in the same gene have different levels of effects on related phenotypes: common variants in *PITX2* are associated with variation in traits (keratometry traits), whereas rare pLoF variants in ClinVar are related to severe diseases (e.g. Axenfeld-Rieger syndrome).

We additionally highlight a set of pleiotropic associations for a common missense variant in *G6PD* in the AFR genetic ancestry group that is rare and thus inaccessible in other genetic ancestry groups. While some of these phenotypes were filtered out by our QC pipeline in AFR, we note that these broad filters do not preclude individual true associations. Fine-mapping of this locus in AFR shows sensible credible sets, whereas fine-mapping across a meta-analyzed cohort leads to instability, particularly in this case where a smaller ancestry group contributes an outsized fraction of the variance explained (i.e. 2pq*β^2^). These results demonstrate a clear need for cohorts with more balanced sample sizes across ancestry groups, such as the burgeoning *All of Us* Research Program cohort^68^. More diverse cohorts will require care when running GWAS to retain high quality association data, particularly as statistical methods often assume homogeneity that can be easily violated when analyzing genomes with high degrees of recent admixture. Scaling methods that consider recent admixture^66^ may improve these analyses further. Additionally, diverse cohorts provide new opportunities to understand heterogeneity in effects, to characterize the influence of variable allele frequency, LD patterns, and environmental factors. While analyzing increasingly diverse GWAS data can be challenging, it is scientifically imperative to accurately identify novel associations, resolve which variants are most likely to be causal, and increase accuracy in polygenic score analysis for all.

In our study, we assigned individuals into distinct ancestry groups. This is a pragmatic approach for statistical analysis that reflects legitimate tradeoffs between controlling type 1 error due to population stratification using meta-analysis versus enhancing statistical power in mega-analyses, which we evaluated empirically. The ideal strategy has not been fully adjudicated in the field, as it is rare that both approaches are equally feasible. Previous studies have shown that results from both approaches are highly comparable.^69,70^ We find outsized benefits to meta-analysis in controlling type 1 error which is especially important when analyzing thousands of traits with widely varying degrees of stratification, and an added benefit is providing ancestry-specific effect size estimates from GWAS that enables more straightforward downstream comparative analyses of genetic effects with datasets from different ancestries. However, this approach simplifies the complex and continuous concept of ancestry into discrete categories and thus risks reifying incorrect assumptions about biological essentialism to racial and ethnic groupings. Readers should be cautious not to infer that these ancestry groups reflect rigid or discrete biological entities; the boundaries between assigned population identifiers are arbitrary and do not necessarily align with biological realities. As such, our use of population grouping should be viewed simply as a methodological convenience. In the future, methods that use continuous measures of genetic diversity or more nuanced approaches to ancestry may provide additional insights and avoid oversimplification. Please refer to our supplementary FAQ for additional context; especially the section ‘Why do you analyze ancestry groups separately?’.

Moreover, the biology of causal variants is mostly shared across populations^14^. In GWAS, differences in MAF and LD due to distinct demographic histories across populations will drive differences in statistical power for discovery, given that the vast majority of GWAS top loci are tagging variants rather than causal. Our detection of GWAS loci unique to particular populations are thus most likely due to improved statistical power for detection in that population, rather than a true causal variant that has distinct effects across ancestry backgrounds. GWAS associations identified only in one ancestry group should thus not be taken as evidence of biological differences between groups. By including multiple ancestral populations in our analyses, we empower identification of loci meaningful to everyone’s health, regardless of ancestry.

Conducting systematic analyses across a massive range of traits raises sensitivities that users of this resource will attempt to make inappropriate comparisons across ancestry groups. Previous work has shown that comparing polygenic score distributions across ancestries is not scientifically meaningful, and we discourage use of this resource to make trait comparisons on the basis of race or ethnicity. We encourage consulting a resource of FAQs we carefully developed as part of this project (https://pan.ukbb.broadinstitute.org/) when using this resource to make any population-based comparisons and evaluating risks versus benefits.

## Acknowledgments

We thank Peter Kraft and Julie-Alexia Dias for helpful discussions. This work was supported by the Novo Nordisk Foundation (NNF21SA0072102), NIH grants R37MH107649, R00MH117229, K01MH121659, F31HL167378, and F30AG074507, and BroadIgnite funding. This study used data from the UK Biobank. UK Biobank has approval from the North West Multi-centre Research Ethics Committee (MREC) as a Research Tissue Bank (RTB) approval. This approval means that researchers do not require separate ethical clearance and can operate under the RTB approval. These analyses were performed under UK Biobank applications 31063 and 95179.

## Data availability

All data are available at https://pan.ukbb.broadinstitute.org/, and sample metadata is available in the UK Biobank showcase under return number 2442: https://biobank.ndph.ox.ac.uk/ukb/dset.cgi?id=2442.

## Extended Data Figures

**Extended Data Figure 1.**
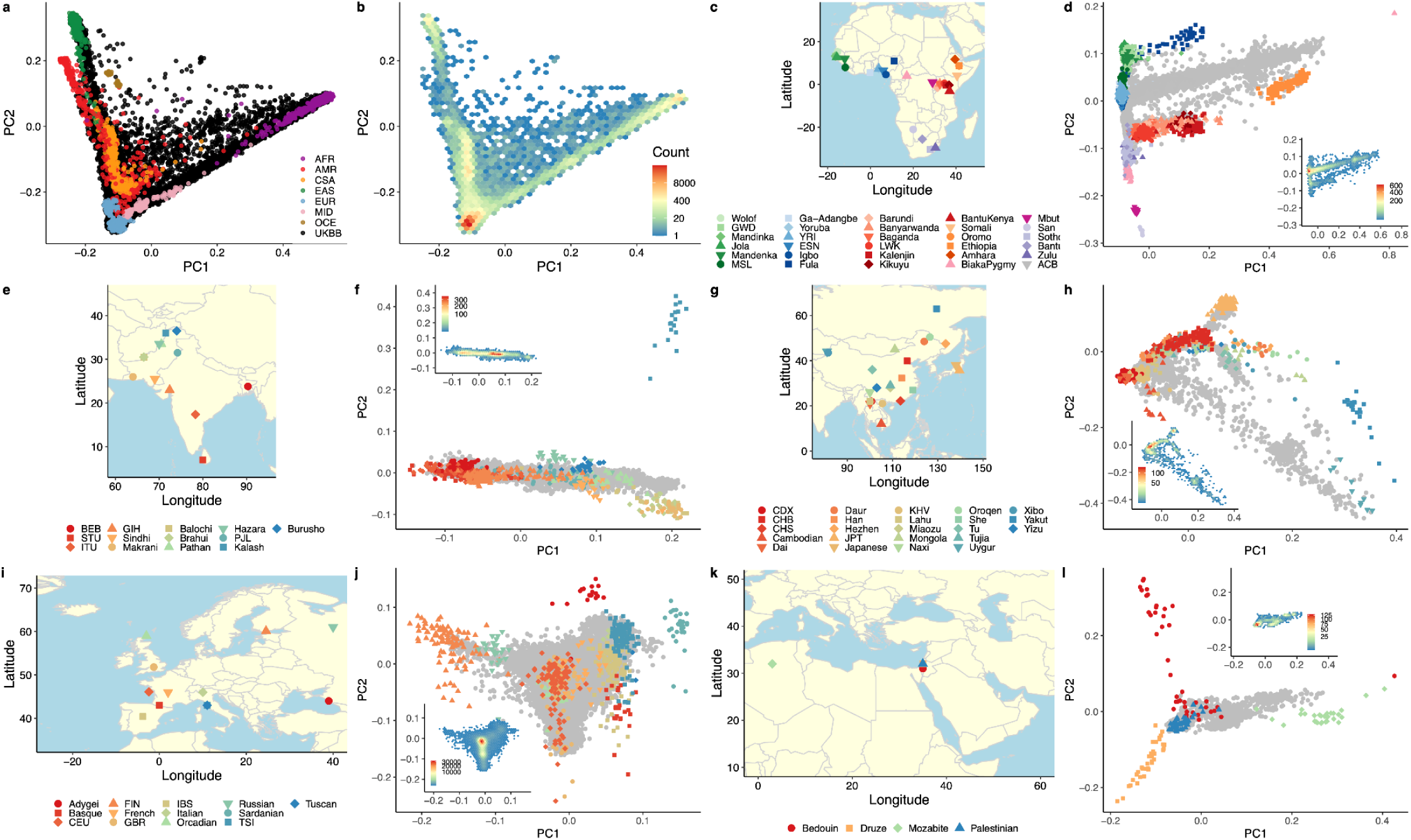
Global and subcontinental PCA. **a**, Global PCA projection of UKBB into PCs 1-2 defined by HGDP and 1000 Genomes Project reference panel, which are shown in colored dots on top of UKBB in black. **b**, Global PCA density plot of UKBB points only, excluding reference panel. **c**, Map of HGDP, 1000 Genomes Project, and AGVP reference used to define AFR PC space. **d**, PCs 1-2 within AFR, reference panel colored, UKBB in grey. Inset shows density of UKBB samples assigned to AFR using a random forest. **c-d**, colors and shapes are consistent across panels. **e**, Map of HGDP and 1000 Genomes Project reference used to define CSA PC space. **f**, PCs 1-2 within CSA, reference panel colored, UKBB in grey. Inset shows density of UKBB samples assigned to CSA using a random forest. **e-f**, colors and shapes are consistent across panels. **g**, Map of HGDP and 1000 Genomes Project reference used to define EAS PC space. **h**, PCs 1-2 within EAS, reference panel colored, UKBB in grey. Inset shows density of UKBB samples assigned to EAS using a random forest. **g-h**, colors and shapes are consistent across panels. **i**, Map of HGDP and 1000 Genomes Project reference used to define EUR PC space. **j**, PCs 1-2 within EUR, reference panel colored, UKBB in grey. Inset shows density of UKBB samples assigned to EUR using a random forest. **i-j**, colors and shapes are consistent across panels. **k**, Map of HGDP and 1000 Genomes Project reference used to define MID PC space. **l**, PCs 1-2 within MID, reference panel colored, UKBB in grey. Inset shows density of UKBB samples assigned to MID using a random forest. **k-l**, colors and shapes are consistent across panels.

**Extended Data Table 1.** Comparison of λ_1000_ and λ_GC_ for five phenotypes across three association study paradigms. λ_GC_ is the genomic inflation factor, which is a measure used to assess the extent of inflation in association test statistics by measuring the median of the observed chi-square test statistics divided by the expected chi-square distribution under the null hypothesis. Similarly, **λ_1000_** is the genomic inflation factor adjusted for sample size to match the scale of a study of 1,000 cases and 1,000 controls with equivalent inflation. For both metrics, values > 1 indicate inflation from polygenic signal, population stratification, or other confounding. We show these metrics for EUR (the European genetic ancestry group alone), mega-analysis (a single association test across all samples), and meta-analysis (across all available population-specific association results). We show comparisons at full sample sizes as well as an equivalent sample size to EUR, with an equivalent number of EUR individuals removed to assess the effect of sample size. λ_1000_ values are all very close to 1, consistent with no significant inflation indicating that our test statistics are in line with the null hypothesis. λ_GC_ values are systematically lower for the meta-analysis versus mega-analysis; since the power of these two approaches should be nearly identical at a given sample size, the lower λ_GC_ indicates that we have likely controlled for population stratification better in the meta-analysis. The number of independent genome-wide significant loci for a range of traits is similar in the meta-analysis versus mega-analysis, indicating that we are not losing appreciable power to identify significant associations. As expected, we are gaining power in the multi-ancestry meta-analysis compared to the EUR-only GWAS, as evidenced by the increase in new loci identified, due to larger sample size and increased diversity. The number of independent GWAS significant loci is computed with an LD r^2^ cutoff of 0.1 and a GWAS significance threshold of 5 x 10^-8^.

| Phenocode |  | 30000 | 30060 | 250.2 | 411 | 495 |
| --- | --- | --- | --- | --- | --- | --- |
| Phenotype |  | White blood cell count | Mean corpuscular haemoglobin concentration | Type 2 Diabetes | Ischemic Heart Disease | Asthma |
| $\lambda_{1000}$ | EUR (full) | 1.0016 | 1.0010 | 1.0038 | 1.0018 | 1.0025 |
|  | Mega-analysis (EUR equivalent sample size) | 1.0016 | 1.0014 | 1.0031 | 1.0022 | 1.0023 |
|  | Meta-analysis (EUR equivalent sample size) | 1.0012 | 1.0006 | 1.0022 | 1.0014 | 1.0014 |
|  | Mega-analysis (full) | 1.0016 | 1.0014 | 1.0028 | 1.0023 | 1.0021 |
|  | Meta-analysis (full) | 1.0012 | 1.0006 | 1.0019 | 1.0012 | 1.0014 |
| $\lambda_{GC}$ | EUR (full) | 1.3360 | 1.2037 | 1.1655 | 1.1225 | 1.1414 |
|  | Mega-analysis (EUR equivalent sample size) | 1.3316 | 1.2841 | 1.1403 | 1.1508 | 1.1341 |
|  | Meta-analysis (EUR equivalent sample size) | 1.2495 | 1.1245 | 1.1027 | 1.0976 | 1.0802 |
|  | Mega-analysis (full) | 1.3368 | 1.3009 | 1.1337 | 1.1650 | 1.1318 |
|  | Meta-analysis (full) | 1.2582 | 1.1327 | 1.0896 | 1.0867 | 1.0837 |
| Number of independent genome-wide significant loci | EUR (full) | 1,375 | 393 | 107 | 74 | 111 |
|  | Mega-analysis (EUR equivalent sample size) | 1,477 | 470 | 121 | 84 | 112 |
|  | Meta-analysis (EUR equivalent sample size) | 1,442 | 440 | 116 | 79 | 112 |
|  | Mega-analysis (full) | 1,580 | 523 | 128 | 86 | 116 |
|  | Meta-analysis (full) | 1,551 | 469 | 124 | 82 | 125 |

**Extended Data Table 2.** Number of significant associations for height in AFR and CSA. The number of variants associated with height at p < 10^-4^ for AFR and CSA are shown (Total), as well as filtered to independent SNPs based on distance-based windows (1 Mb). Despite the smaller sample size, the AFR ancestry group identified 15% more significant associations than CSA (1,261 vs 1,089).

|  | AFR (N: 6,556) |  | CSA (N: 8,657) |  |
| --- | --- | --- | --- | --- |
|  | Total | Regionally-independent | Total | Regionally-independent |
| **Significant variants** | 4,682 | 1,380 | 4,284 | 1,212 |
| **High-quality significant variants** | 3,016 | 1,261 | 2,695 | 1,089 |

**Extended Data Figure 2.**
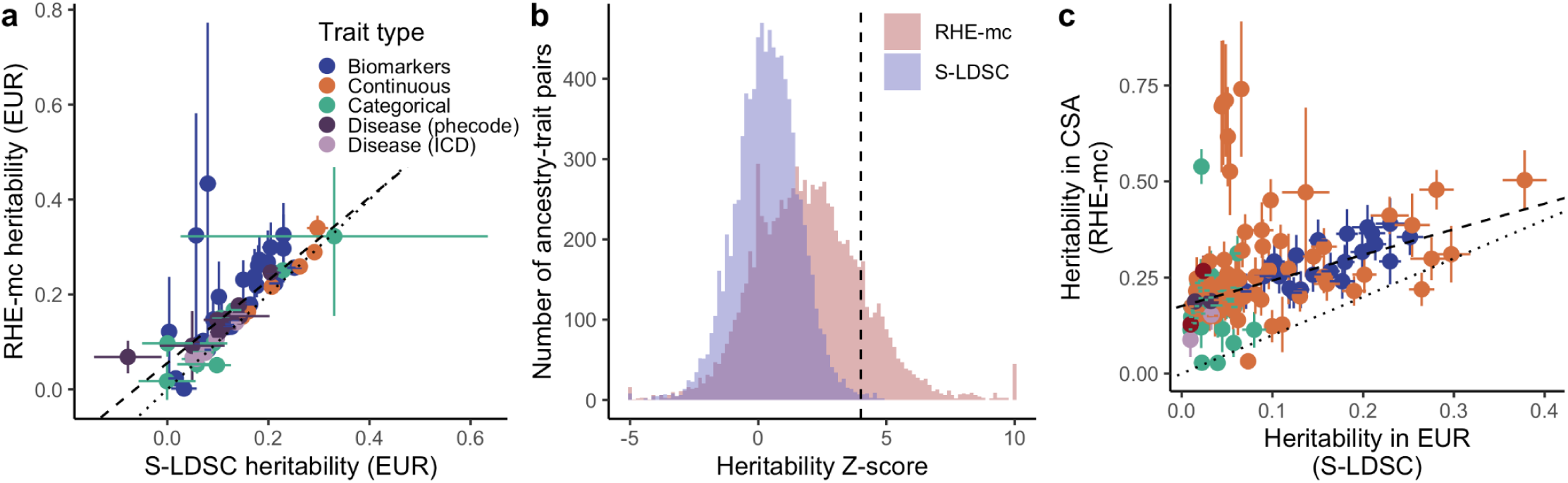
Heritability informs the robustness of GWAS across ancestry-trait pairs. **a,** heritability estimates are generally concordant in the EUR genetic ancestry group across 64 pilot phenotypes (**Supplementary Table 9**) and two statistical methods. RHE-mc uses a randomized multi-component version of classical Haseman-Elston regression with a genetic relatedness matrix^54^, whereas S-LDSC uses GWAS summary statistics^53^. For binary phenotypes, heritability estimates are reported on the liability scale. All pilot phenotypes are shown, except for sepsis which had negative heritability estimates by both methods. The dotted line shows y=x, while the dashed line is a fitted linear regression (slope = 0.87, intercept = 0.05, p = 7 x 10^-13^). **b**, Across the same non-EUR ancestry-trait pairs, heritability estimated with RHE-mc have higher z-scores due to the smaller standard errors compared to S-LDSC. Dashed line at Z=4 was used as a QC filter. **c**, As in Fig. 2b, without filtering to phenotypes passing QC, but instead only filtering to EUR z > 4 and defined heritability in both genetic ancestry groups. Dotted line shows y = x and dashed line shows York regression fit (slope = 0.66, intercept = 0.17, p < 10^-100^).

**Extended Data Figure 3.**
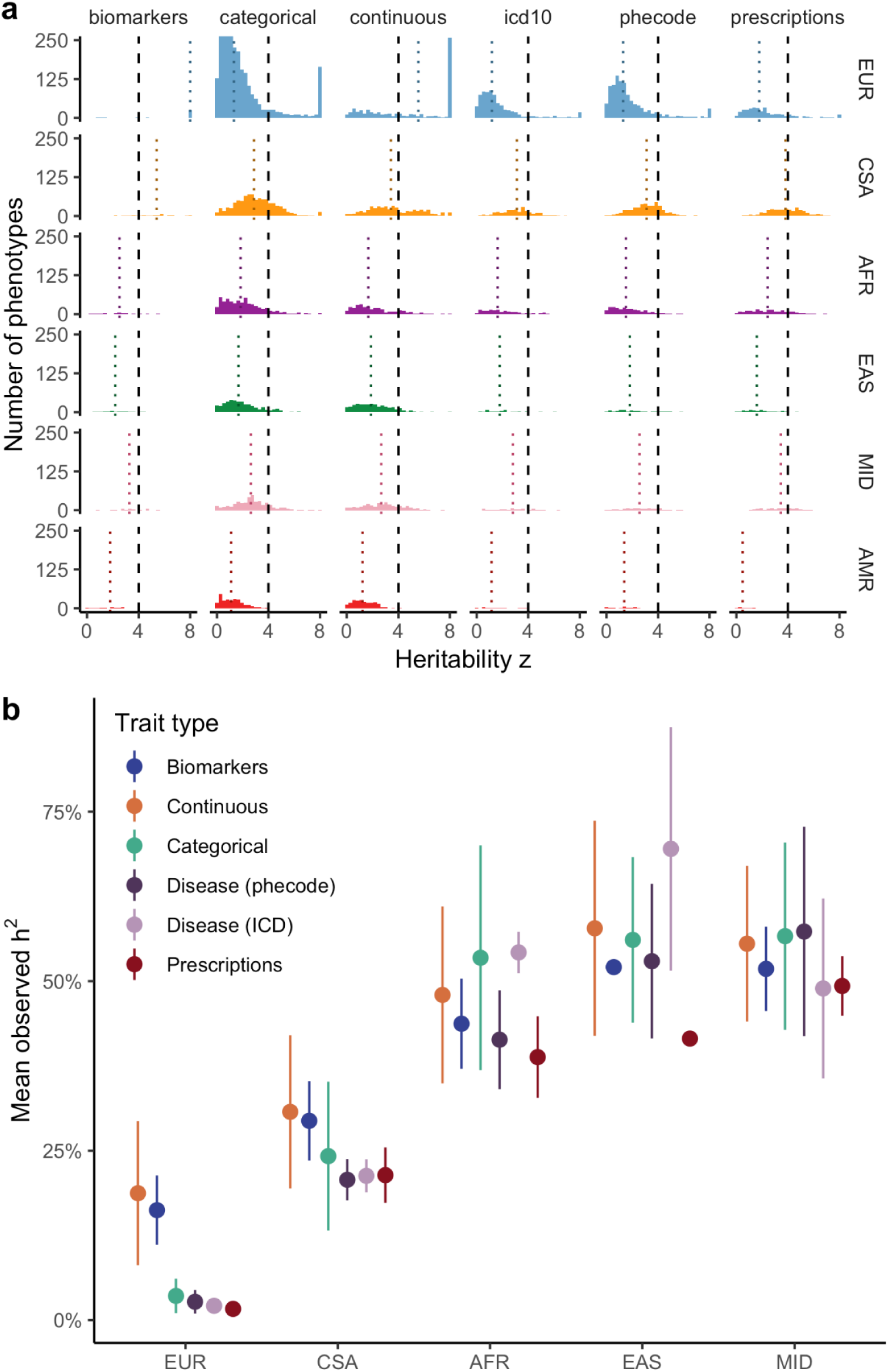
Heritability summaries across trait types and genetic ancestry groups. **(a)** The confidence metrics (heritability *z* score) across traits (columns) and ancestry groups (rows) are shown for the final heritability metrics used (S-LDSC for EUR, otherwise RHE-mc). Dashed line indicates inclusion criteria (*z* ≥ 4). (**b**) The mean observed heritability (h^2^) is plotted by ancestry group and trait type. For ancestry groups with smaller sample sizes, heritabilities are likely inflated due to a combination of residual stratification and winner’s curse, as only significantly heritable phenotypes in each ancestry group are shown. Error bars are standard deviations of the distribution of the heritability point estimates.

**Extended Data Figure 4.**
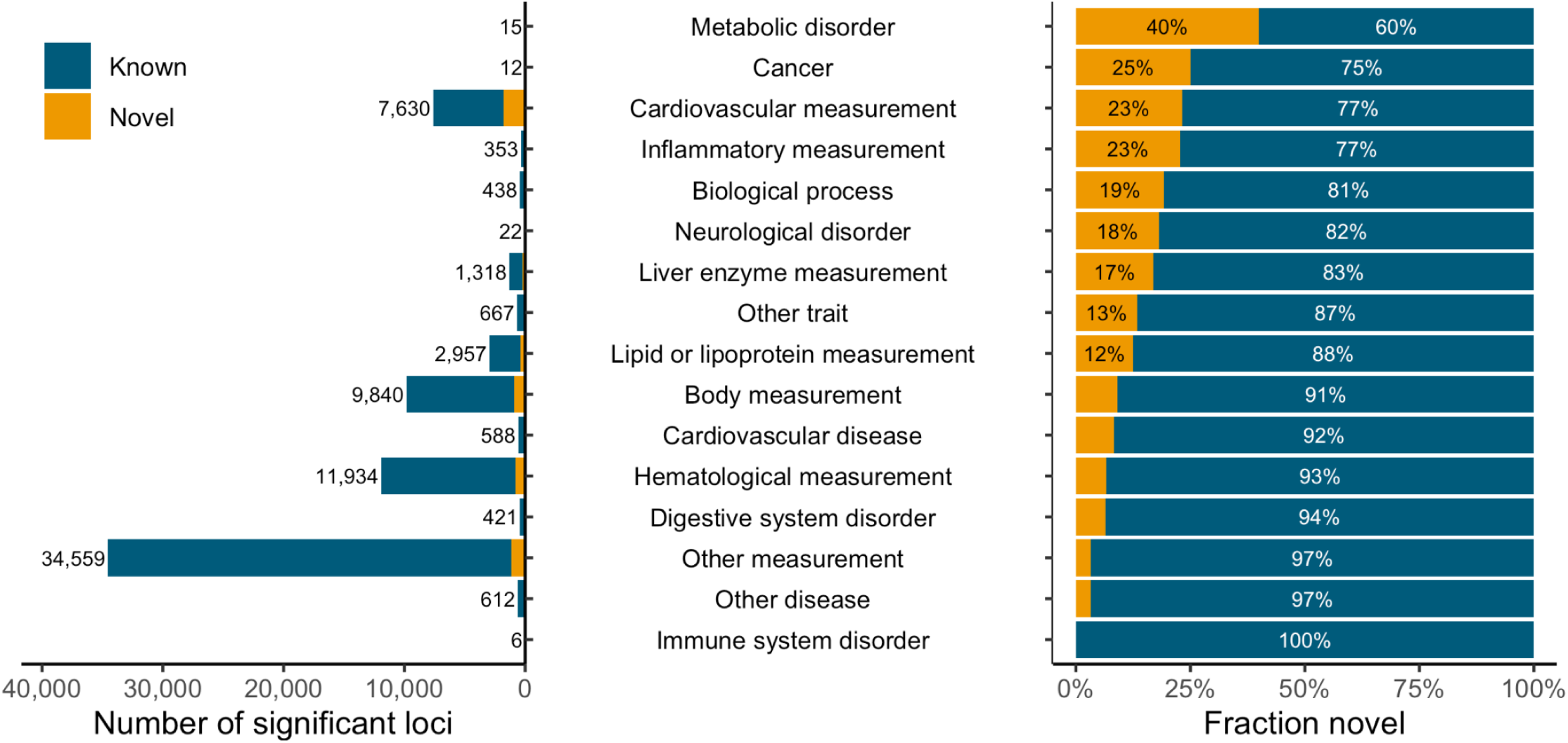
Improved identification of associations by EFO category. Number (left) and percentage (right) of known and novel variants identified in this study compared to the GWAS catalog across EFO categories.

**Extended Data Figure 5.**
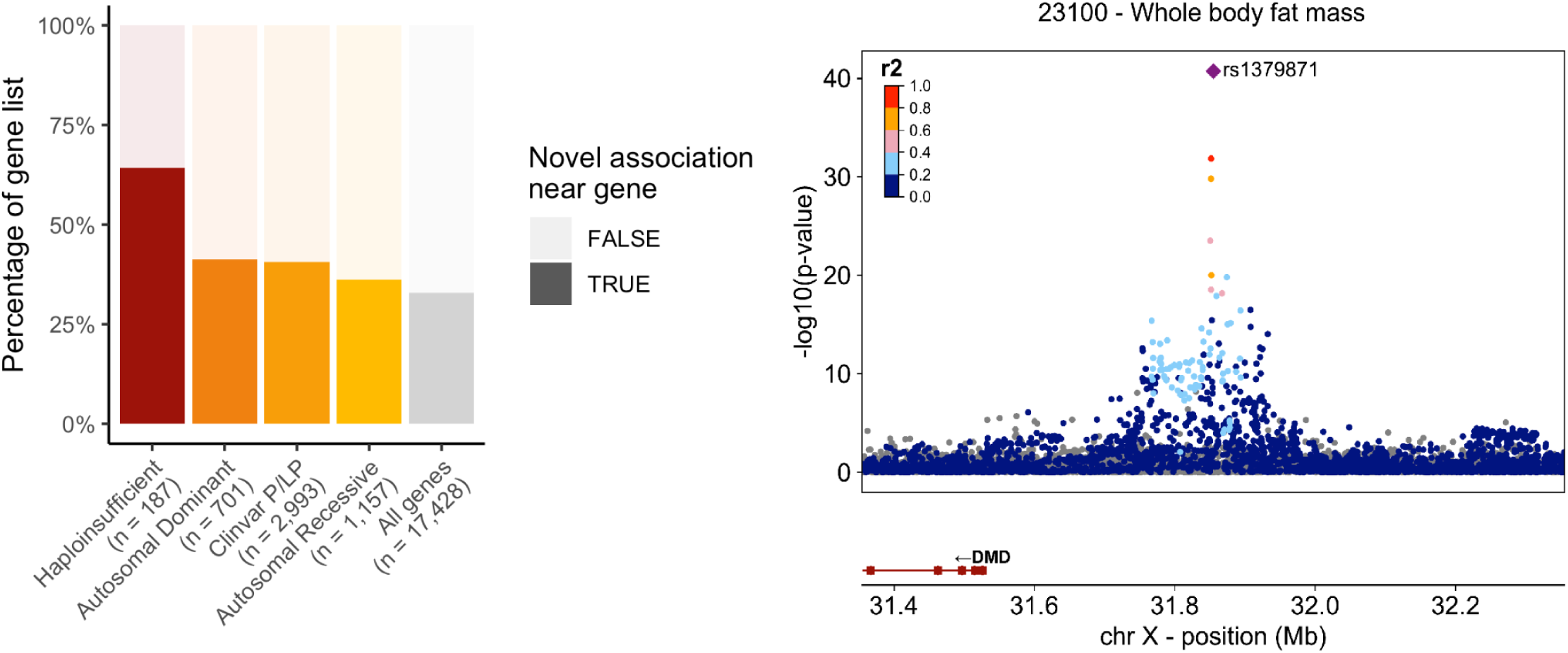
GWAS hits near haploinsufficient genes. **a**, The percentage of novel associations by gene category. 66% of haploinsufficient genes have a novel significant hit nearby, compared to 34% of all genes. **b**, Locuszoom plots of a 1Mb region around rs1379871 (purple diamond; *DMD*), for whole body fat mass (p = 1.84 x 10^-41^; N = 431,792). The -log10(p-value) is plotted along chromosomal position, with neighboring variants colored by sample-size weighted LD (with lead SNP) for ancestries included in meta-analysis (gray: LD not defined for at least one ancestry group). This variant has recently been identified in a larger study of BMI^71^.

**Extended Data Figure 6.**
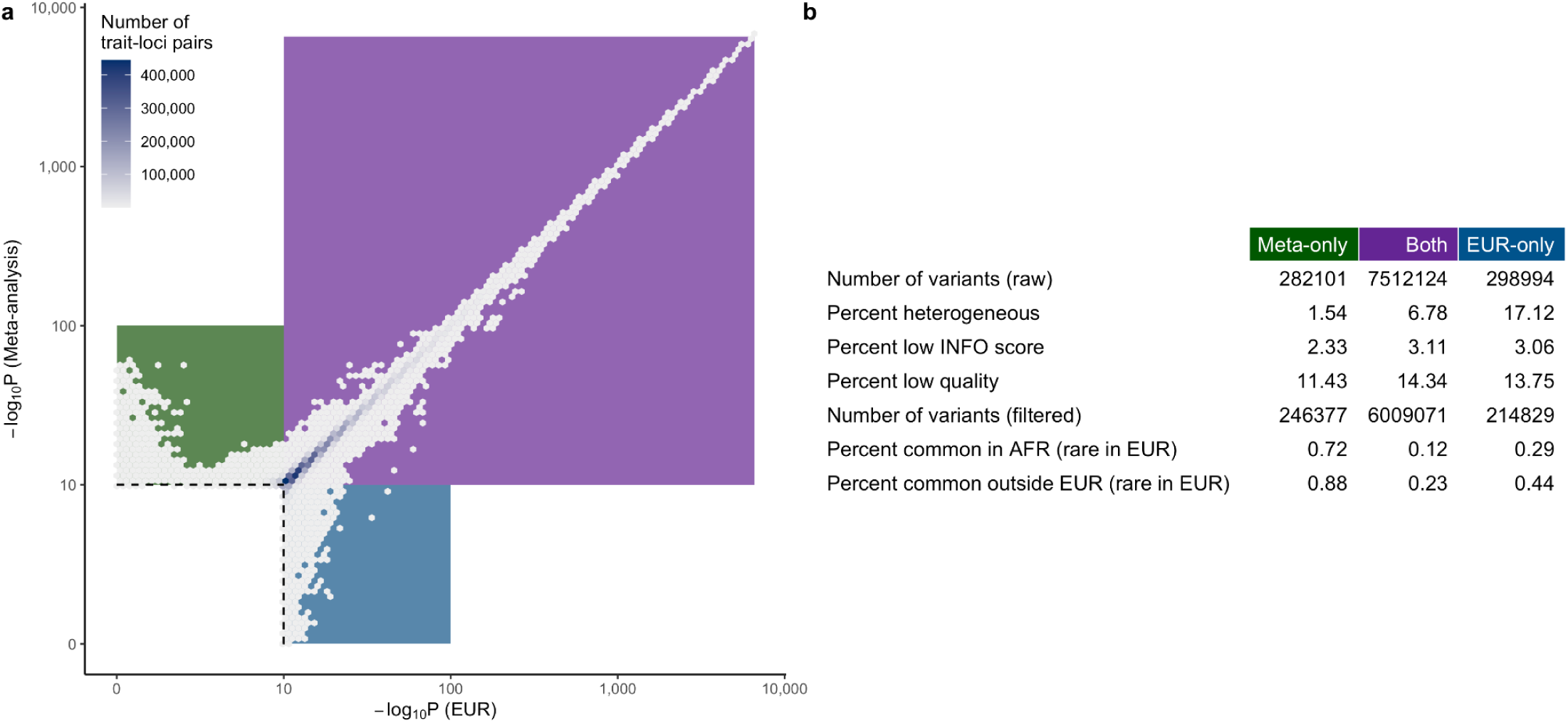
Comparison of meta-analysis and EUR summary statistics. **a,** As in Fig. 3c, the p-value in EUR is plotted compared to the p-value in the meta-analysis, as a density plot to indicate the relative number of points in each region of the plot. Three quadrants are highlighted for significant in meta-analysis only (green), both meta-analysis and EUR (purple), and EUR-only (blue). **b,** Summaries and meta-data of the variants in each of these three quadrants are shown. Heterogeneous is defined as Cochran’s Q p < 0.01, low INFO score is defined as INFO < 0.9, and low quality is defined as failing quality filters from gnomAD or allele frequency significantly differing between gnomAD and Pan-UKB in at least one ancestry group (see Supplementary Information, QC of summary statistics). Common is defined as frequency > 1%.

**Extended Data Figure 7.**
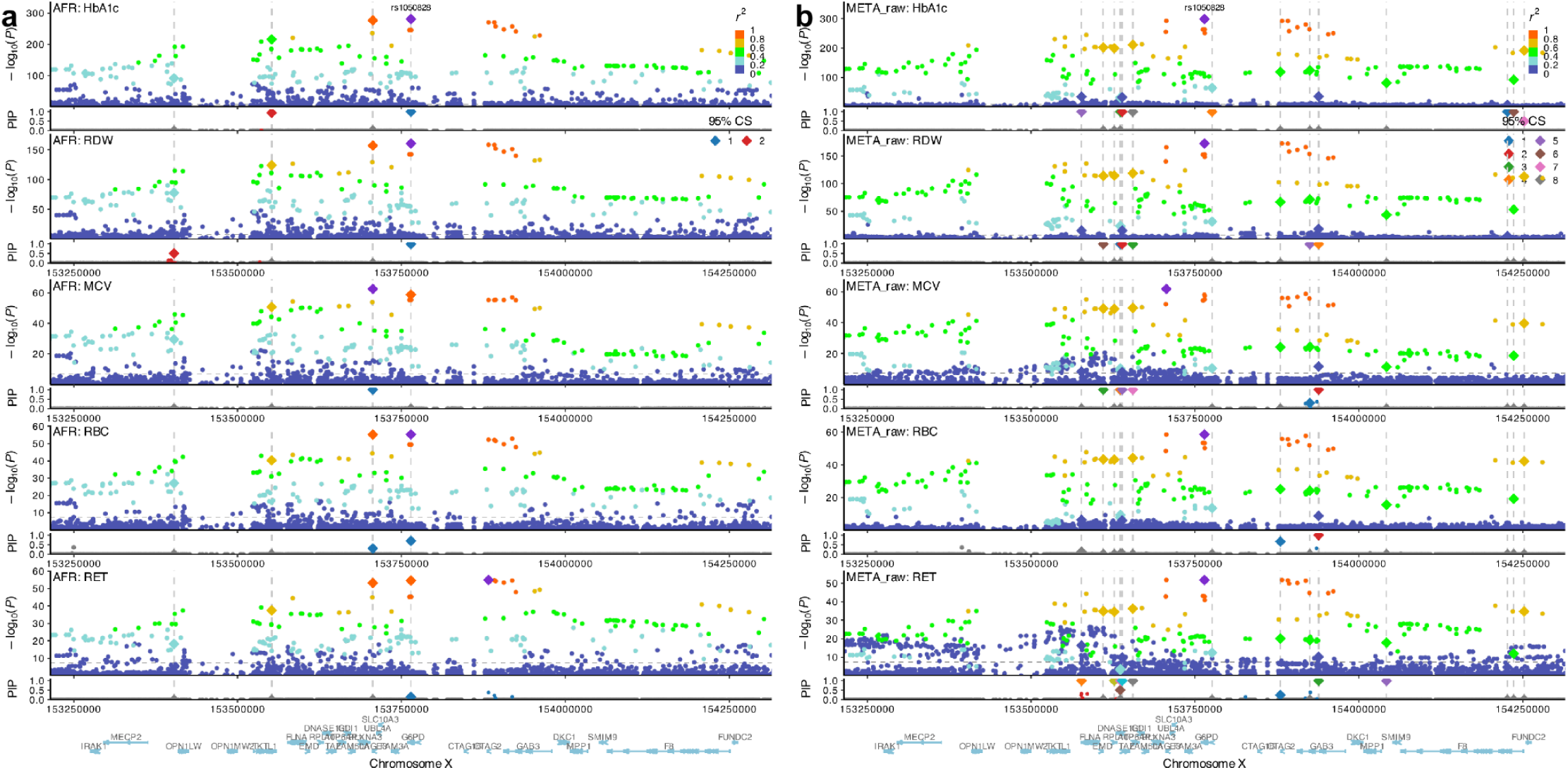
Fine-mapping of the G6PD locus. Fine-mapping results for rs1050828 (*G6PD*) in (**a**) AFR and (**b**) meta-analysis. **a**, AFR fine-mapping results highlight the missense variant (rs1050828) in a credible set, with a second independent signal for some phenotypes. **b**, Meta-analysis fine-mapped results show instability as the major signal at rs1050828 is discovered in a group with a relatively small sample size, which results in a small contribution to the LD panel and thus, poor performance in fine-mapping.

**Extended Data Figure 8.**
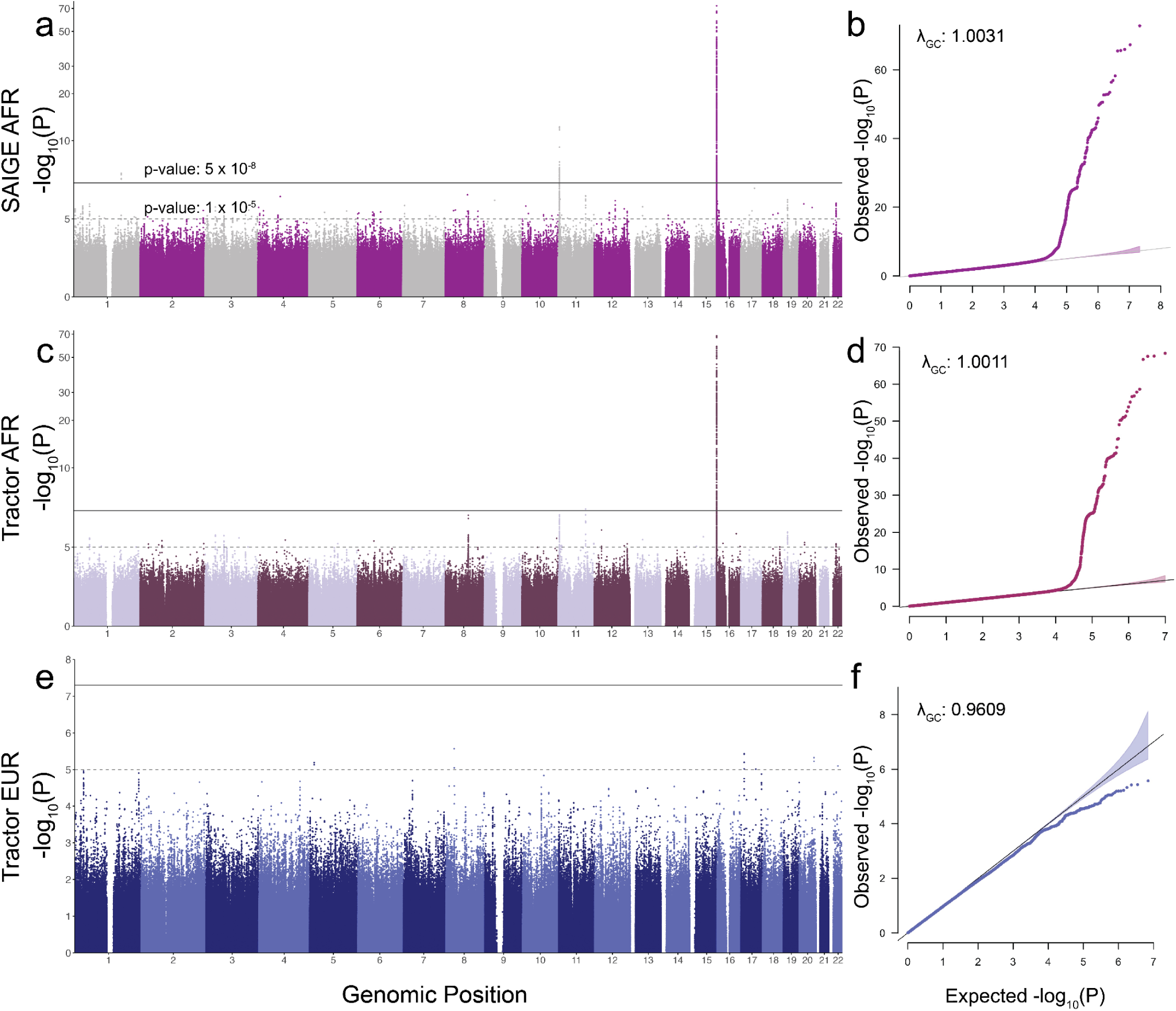
Manhattan and QQ plot comparison for SAIGE and *Tractor* GWAS for mean corpuscular hemoglobin concentration. a-b, Original GWAS performed using SAIGE for AFR. **c-f,** Among AFR individuals, Tractor GWAS results are shown for AFR haplotypes **(c, d)** and EUR haplotypes **(e, f)**.

## References

1. Abul-Husn, N. S. & Kenny, E. E. Personalized Medicine and the Power of Electronic Health Records. Cell 177, 58–69 (2019).

2. Zhou, W. et al. Global Biobank Meta-analysis Initiative: Powering genetic discovery across human disease. Cell Genom 2, 100192 (2022).

3. Buniello, A. et al. The NHGRI-EBI GWAS Catalog of published genome-wide association studies, targeted arrays and summary statistics 2019. Nucleic Acids Res. 47, D1005–D1012 (2019).

4. Sirugo, G., Williams, S. M. & Tishkoff, S. A. The Missing Diversity in Human Genetic Studies. Cell 177, 26–31 (2019).

5. Martin, A. R. et al. Clinical use of current polygenic risk scores may exacerbate health disparities. Nat. Genet. 51, 584–591 (2019).

6. Morales, J. et al. A standardized framework for representation of ancestry data in genomics studies, with application to the NHGRI-EBI GWAS Catalog. Genome Biol. 19, 21 (2018).

7. Witherspoon, D. J. et al. Genetic similarities within and between human populations. Genetics 176, 351–359 (2007).

8. Bamshad, M., Wooding, S., Salisbury, B. A. & Stephens, J. C. Deconstructing the relationship between genetics and race. Nat. Rev. Genet. 5, 598–609 (2004).

9. National Academies of Sciences, Engineering, and Medicine. Using Population Descriptors in Genetics and Genomics Research: A New Framework for an Evolving Field. (National Academies Press, 2023).

10. Meyer, M. N. et al. Wrestling with Social and Behavioral Genomics: Risks, Potential Benefits, and Ethical Responsibility. Hastings Cent. Rep. 53 Suppl 1, S2–S49 (2023).

11. Sinnott-Armstrong, N. et al. Genetics of 35 blood and urine biomarkers in the UK Biobank. Nat. Genet. 53, 185–194 (2021).

12. Lam, M. et al. Comparative genetic architectures of schizophrenia in East Asian and European populations. Nat. Genet. 1–9 (2019).

13. Chen, J. et al. The trans-ancestral genomic architecture of glycemic traits. Nat. Genet. 53, 840–860 (2021).

14. Hou, K. et al. Causal effects on complex traits are similar for common variants across segments of different continental ancestries within admixed individuals. Nat. Genet. 55, 549–558 (2023).

15. SIGMA Type 2 Diabetes Consortium et al. Association of a low-frequency variant in HNF1A with type 2 diabetes in a Latino population. JAMA 311, 2305–2314 (2014).

16. Cohen, J. et al. Low LDL cholesterol in individuals of African descent resulting from frequent nonsense mutations in PCSK9. Nat. Genet. 37, 161–165 (2005).

17. Liu, Z. et al. Genetic architecture of the inflammatory bowel diseases across East Asian and European ancestries. Nat. Genet. 55, 796–806 (2023).

18. Miller, L. H., Mason, S. J., Clyde, D. F. & McGinniss, M. H. The resistance factor to Plasmodium vivax in blacks. The Duffy-blood-group genotype, FyFy. N. Engl. J. Med. 295, 302–304 (1976).

19. Genovese, G. et al. Association of trypanolytic ApoL1 variants with kidney disease in African Americans. Science 329, 841–845 (2010).

20. Ross, M. J. New Insights into APOL1 and Kidney Disease in African Children and Brazilians Living With End-Stage Kidney Disease. Kidney Int Rep 4, 908–910 (2019).

21. Genovese, G., Friedman, D. J. & Pollak, M. R. APOL1 variants and kidney disease in people of recent African ancestry. Nat. Rev. Nephrol. 9, 240–244 (2013).

22. Mägi, R. et al. Trans-ethnic meta-regression of genome-wide association studies accounting for ancestry increases power for discovery and improves fine-mapping resolution. Hum. Mol. Genet. 26, 3639–3650 (2017).

23. Asimit, J. L., Hatzikotoulas, K., McCarthy, M., Morris, A. P. & Zeggini, E. Trans-ethnic study design approaches for fine-mapping. Eur. J. Hum. Genet. 24, 1330–1336 (2016).

24. Mahajan, A., et al. Trans-ancestry genetic study of type 2 diabetes highlights the power of diverse populations for discovery and translation. medRxiv (2020).

25. Huang, H. et al. Fine-mapping inflammatory bowel disease loci to single-variant resolution. Nature 547, 173–178 (2017).

26. Schaid, D. J., Chen, W. & Larson, N. B. From genome-wide associations to candidate causal variants by statistical fine-mapping. Nat. Rev. Genet. 19, 491–504 (2018).

27. Graff, M. et al. Discovery and fine-mapping of height loci via high-density imputation of GWASs in individuals of African ancestry. Am. J. Hum. Genet. 108, 564–582 (2021).

28. Luo, Y., et al. A high-resolution HLA reference panel capturing global population diversity enables multi-ethnic fine-mapping in HIV host response. medRxiv (2020).

29. Polygenic Risk Score Task Force of the International Common Disease Alliance. Responsible use of polygenic risk scores in the clinic: potential benefits, risks and gaps. Nat. Med. 27, 1876–1884 (2021).

30. Martin, A. R. et al. Human Demographic History Impacts Genetic Risk Prediction across Diverse Populations. Am. J. Hum. Genet. 100, 635–649 (2017).

31. Scutari, M., Mackay, I. & Balding, D. Using Genetic Distance to Infer the Accuracy of Genomic Prediction. PLoS Genet. 12, e1006288 (2016).

32. Wang, Y. et al. Theoretical and empirical quantification of the accuracy of polygenic scores in ancestry divergent populations. bioRxiv 2020.01.14.905927 (2020) doi:10.1101/2020.01.14.905927.

33. Ding, Y. et al. Polygenic scoring accuracy varies across the genetic ancestry continuum. Nature 618, 774–781 (2023).

34. Conti, D. V. et al. Trans-ancestry genome-wide association meta-analysis of prostate cancer identifies new susceptibility loci and informs genetic risk prediction. Nat. Genet. 53, 65–75 (2021).

35. Bigdeli, T. B. et al. Contributions of common genetic variants to risk of schizophrenia among individuals of African and Latino ancestry. Mol. Psychiatry (2019) doi:10.1038/s41380-019-0517-y.

36. Bastarache, L. Using Phecodes for Research with the Electronic Health Record: From PheWAS to PheRS. (2021) doi:10.1146/annurev-biodatasci-122320-112352.

37. Denny, J. C. et al. Systematic comparison of phenome-wide association study of electronic medical record data and genome-wide association study data. Nat. Biotechnol. 31, 1102–1110 (2013).

38. Bastarache, L. et al. Phenotype risk scores identify patients with unrecognized Mendelian disease patterns. Science 359, 1233–1239 (2018).

39. Zheng, J. et al. LD Hub: a centralized database and web interface to perform LD score regression that maximizes the potential of summary level GWAS data for SNP heritability and genetic correlation analysis. Bioinformatics 33, 272–279 (2017).

40. Ben-Eghan, C. et al. Don’t ignore genetic data from minority populations. Nature vol. 585 184–186 Preprint at 10.1038/d41586-020-02547-3 (2020).

41. Bycroft, C. et al. The UK Biobank resource with deep phenotyping and genomic data. Nature 562, 203–209 (2018).

42. 1000 Genomes Project Consortium et al. A global reference for human genetic variation. Nature 526, 68–74 (2015).

43. Li, J. Z. et al. Worldwide human relationships inferred from genome-wide patterns of variation. Science 319, 1100–1104 (2008).

44. Mathieson, I. & Scally, A. What is ancestry? PLoS Genet. 16, e1008624 (2020).

45. Lewis, A. C. F. et al. Getting genetic ancestry right for science and society. Science 376, 250–252 (2022).

46. National Academies of Sciences, Engineering, and Medicine et al. Using Population Descriptors in Genetics and Genomics Research: A New Framework for an Evolving Field. (National Academies Press, 2023).

47. Zhou, W. et al. Efficiently controlling for case-control imbalance and sample relatedness in large-scale genetic association studies. Nat. Genet. 50, 1335–1341 (2018).

48. Schizophrenia Psychiatric Genome-Wide Association Study (GWAS) Consortium. Genome-wide association study identifies five new schizophrenia loci. Nat. Genet. 43, 969–976 (2011).

49. COVID-19 Host Genetics Initiative. Mapping the human genetic architecture of COVID-19. Nature (2021) doi:10.1038/s41586-021-03767-x.

50. Purcell, S. et al. PLINK: a tool set for whole-genome association and population-based linkage analyses. Am. J. Hum. Genet. 81, 559–575 (2007).

51. Howrigan, D. Details and Considerations of the UK Biobank GWAS. Preprint at (2017).

52. Bulik-Sullivan, B. et al. An atlas of genetic correlations across human diseases and traits. Nat. Genet. 47, 1236–1241 (2015).

53. Finucane, H. K. et al. Partitioning heritability by functional annotation using genome-wide association summary statistics. Nat. Genet. 47, 1228–1235 (2015).

54. Pazokitoroudi, A. et al. Efficient variance components analysis across millions of genomes. Nat. Commun. 11, 4020 (2020).

55. Ghoussaini, M. et al. Open Targets Genetics: systematic identification of trait-associated genes using large-scale genetics and functional genomics. Nucleic Acids Res. 49, D1311–D1320 (2020).

56. Solovieff, N., Cotsapas, C., Lee, P. H., Purcell, S. M. & Smoller, J. W. Pleiotropy in complex traits: challenges and strategies. Nat. Rev. Genet. 14, 483–495 (2013).

57. Sun, L., Wang, Z., Lu, T., Manolio, T. A. & Paterson, A. D. eXclusionarY: 10 years later, where are the sex chromosomes in GWASs? Am. J. Hum. Genet. 110, 903–912 (2023).

58. Rasooly, D. et al. Genome-wide association analysis and Mendelian randomization proteomics identify drug targets for heart failure. Nat. Commun. 14, 3826 (2023).

59. Gage, P. J., Suh, H. & Camper, S. A. Dosage requirement of Pitx2 for development of multiple organs. Development 126, 4643–4651 (1999).

60. Tümer, Z. & Bach-Holm, D. Axenfeld-Rieger syndrome and spectrum of PITX2 and FOXC1 mutations. Eur. J. Hum. Genet. 17, 1527–1539 (2009).

61. Berry, F. B. et al. Functional interactions between FOXC1 and PITX2 underlie the sensitivity to FOXC1 gene dose in Axenfeld-Rieger syndrome and anterior segment dysgenesis. Hum. Mol. Genet. 15, 905–919 (2006).

62. Gibson, G. Population genetics and GWAS: A primer. PLoS Biol. 16, e2005485 (2018).

63. Martin, A. R., Daly, M. J., Robinson, E. B., Hyman, S. E. & Neale, B. M. Predicting Polygenic Risk of Psychiatric Disorders. Biol. Psychiatry 86, 97–109 (2019).

64. Liu, D. J. & Leal, S. M. Estimating genetic effects and quantifying missing heritability explained by identified rare-variant associations. Am. J. Hum. Genet. 91, 585–596 (2012).

65. Sarnowski, C. et al. Impact of Rare and Common Genetic Variants on Diabetes Diagnosis by Hemoglobin A1c in Multi-Ancestry Cohorts: The Trans-Omics for Precision Medicine Program. Am. J. Hum. Genet. 105, 706–718 (2019).

66. Atkinson, E. G. et al. Tractor uses local ancestry to enable the inclusion of admixed individuals in GWAS and to boost power. Nat. Genet. 53, 195–204 (2021).

67. Zhou, W. et al. Global Biobank Meta-analysis Initiative: powering genetic discovery across human diseases. bioRxiv (2021) doi:10.1101/2021.11.19.21266436.

68. All of Us Research Program Genomics Investigators. Genomic data in the All of Us Research Program. Nature (2024) doi:10.1038/s41586-023-06957-x.

69. Panagiotou, O. A., Willer, C. J., Hirschhorn, J. N. & Ioannidis, J. P. A. The power of meta-analysis in genome-wide association studies. Annu. Rev. Genomics Hum. Genet. 14, 441–465 (2013).

70. Lin, D. Y. & Zeng, D. Meta-analysis of genome-wide association studies: no efficiency gain in using individual participant data. Genet. Epidemiol. 34, 60–66 (2010).

71. Zhang, X. et al. WHOLE GENOME SEQUENCING ANALYSIS OF BODY MASS INDEX IDENTIFIES NOVEL AFRICAN ANCESTRY-SPECIFIC RISK ALLELE. medRxiv (2023) doi:10.1101/2023.08.21.23293271.

