## Supplementary Information for "Pan-UK Biobank GWAS improves discovery, analysis of genetic architecture, and resolution into ancestry-enriched effects"

### UKB Pan Ancestry Supplement


UKB Pan Ancestry Supplement 1

Ancestry analysis and relatedness inference 8

Initial ancestry assignment 8

Supplementary Figure 1 | Genetic ancestry assignment using principal components analysis (PCA). A-C) First 6 PCA biplots for UKB individuals in black with reference populations from the 1000 Genomes Project and HGDP on top colored by continental ancestry. D-F) Density of UKB participant global PCs. G-I) Genetic ancestry assignments for UKB participants based on reference panel meta-data labels. No reference data are included in these plots. A, D, and G) PCA biplots for PCs 1-2; B, E, and H) PCs 3-4; C, F, and I) PCs 5-6. 3-letter continental ancestry codes are as in Supplementary Table 1. 10

Supplementary Table 1 | Genetic reference panel data from the 1000 Genomes Project (1KG) and Human Genome Diversity Project (HGDP). Continental ancestries are EUR=European, CSA=Central/South Asian, AFR=African, AMR=Admixed American, EAS=East Asian, MID=Middle Eastern, and OCE=Oceanian. Population abbreviations with three-letter codes are the same as in the 1000 Genomes Project. 11

Supplementary Table 2 | Comparison of ancestry assignments based on random forests with 6 principal components and a range of probability thresholds. 12

Supplementary Table 3 | Comparison between ancestry assignments using a random forest trained on 6 principal components (PCs) versus 20 PCs. Assignments based on 6 PCs are shown in rows, while assignments based on 20 PCs are shown in columns. More individuals are assigned to “Other” using the random forest based on 20 PCs. 12

Supplementary Table 4 | Counts of individuals by continental ancestry as inferred from the 1000 Genomes and Human Genome Diversity Panel genetic data. Oceanians were removed from further analyses given the very small sample sizes. Those individuals whose ancestry groups could not be assigned (i.e., “Other”) were also removed. The number of outliers removed indicates the count remaining after pruning ancestry outliers according to PC coordinates. 3-letter continental ancestry codes are as in Supplementary Table 1. 12

Visualizing subcontinental ancestries 13

Supplementary Table 5 | Additional genetic reference panel data from the African Genome Variation Project (AGVP) used for visualization of subcontinental PCA within African ancestry assignments (AFR). 14

Supplementary Figure 2 | Subcontinental ancestry PCs in the AFR reference panel and UKB participants assigned to AFR. UKB participants assigned to AFR are shown in grey, while reference populations are on top and colored as in the map. A) Map of reference populations, B) PCs 1-2, C) PCs 3-4, D) PCs 5-6. Left panels show PC values, right panels show UKB PC densities. 15

Supplementary Figure 3 | Subcontinental ancestry PCs in the AMR reference panel and UKB participants assigned to AMR. UKB participants assigned to AMR are shown in grey, while reference populations are on top and colored as in the map. A) Map of reference populations, B) PCs 1-2, C) PCs 3-4, D) PCs 5-6. Left panels show PC values, right panels show UKB PC densities. 16

Supplementary Figure 4 | Subcontinental ancestry PCs in the CSA reference panel and UKB participants assigned to CSA. UKB participants assigned to CSA are shown in grey, while reference populations are on top and colored as in the map. A) Map of reference populations, B) PCs 1-2, C) PCs 3-4, D) PCs 5-6. Left panels show PC values, right panels show UKB PC densities. 17

Supplementary Figure 5 | Subcontinental ancestry PCs in the EAS reference panel and UKB participants assigned to EAS. UKB participants assigned to EAS are shown in grey, while reference populations are on top and colored as in the map. A) Map of reference populations, B) PCs 1-2, C) PCs 3-4, D) PCs 5-6. Left panels show PC values, right panels show UKB PC densities. 18

Supplementary Figure 6 | Subcontinental ancestry PCs in the EUR reference panel and UKB participants assigned to EUR. UKB participants assigned to EUR are shown in grey, while reference populations are on top and colored as in the map. A) Map of reference populations, B) PCs 1-2, C) PCs 3-4, D) PCs 5-6. Left panels show PC values, right panels show UKB PC densities. 19

Supplementary Figure 7 | Subcontinental ancestry PCs in the MID reference panel and UKB participants assigned to MID. UKB participants assigned to MID are shown in grey, while reference populations are on top and colored as in the map. A) Map of reference populations, B) PCs 1-2, C) PCs 3-4, D) PCs 5-6. Left panels show PC values, right panels show UKB PC densities. 20

Pruning ancestry outliers 21

Supplementary Figure 8 | PCA in UKB participants assigned to AFR and corresponding centroid distance across 3 PCs. Centroid distance distributions and PC biplots for the first 6 PCs are shown before (top) and after (bottom) pruning outliers. Vertical line in the top left centroid distance histogram shows the threshold chosen to remove outliers. 22

Supplementary Figure 9 | PCA in UKB participants assigned to AMR and corresponding centroid distance across 3 PCs. Centroid distance distributions and PC biplots for the first 6 PCs are shown before (top) and after (bottom) pruning outliers. Vertical line in the top left centroid distance histogram shows the threshold chosen to remove outliers. 23

Supplementary Figure 10 | PCA in UKB participants assigned to CSA and corresponding centroid distance across 3 PCs. Centroid distance distributions and PC biplots for the first 6 PCs are shown before (top) and after (bottom) pruning outliers. Vertical line in the top left centroid distance histogram shows the threshold chosen to remove outliers. 24

Supplementary Figure 11 | PCA in UKB participants assigned to EAS and corresponding centroid distance across 3 PCs. Centroid distance distributions and PC biplots for the first 6 PCs are shown before (top) and after (bottom) pruning outliers. Vertical line in the top left centroid distance histogram shows the threshold chosen to remove outliers. 25

Supplementary Figure 12 | PCA in UKB participants assigned to EUR and corresponding centroid distance across 5 PCs. Centroid distance distributions and PC biplots for the first 6 PCs are shown before (top) and after (bottom) pruning outliers. Vertical line in the top left centroid distance histogram shows the threshold chosen to remove outliers. 26

Supplementary Figure 13 | PCA in UKB participants assigned to MID and corresponding centroid distance across 5 PCs. Centroid distance distributions and PC biplots for the first 6 PCs are shown before (top) and after (bottom) pruning outliers. Vertical line in the top left centroid distance histogram shows the threshold chosen to remove outliers. 27

Relationship between ancestry and self-reported metrics 27

Supplementary Table 6 | Comparison between inferred genetic ancestry (columns) and self-reported ethnicity (rows). UKB code 21000 provided self-reported ethnic background. Codings and meanings are defined by the UKB. 28

Supplementary Figure 14 | Principal components roughly correlate with self-reported ethnicity. Principal components are as shown in Supplementary Fig. 1. 29

Supplementary Table 7 | Assigned population labels correlate with continental birthplaces.

Columns show assigned population labels. Rows indicate continental birthplaces. 3-letter continental ancestry codes are as in Supplementary Table 1. Cells shaded in green indicate the maximum fraction per row and column, which was used to calculate marginal percentages. Most shaded cells indicate the maximum for both rows and columns, but blue cells indicate which values were used to calculate only row marginals and yellow cells indicate which value was used to calculate only a column marginal when multiple cells in the row or column were shaded, respectively. 29

Pre-GWAS quality control 30

Sample QC 30

Variant QC 30

Supplementary Table 8 | Number of variants per population. 30

Phenotype QC 30

Continuous and categorical traits 30

Supplementary Figure 15 | Phenotype curation using a custom version of PHESANT. The flowchart summarizes filtering and transformation steps to parse the unprocessed phenotype data. Figure adapted from Millard et al15. 31

ICD-10 codes 32

Phecodes 32

Prescriptions 32

Other phenotypes 33

Association analysis 34

Computational framework 34

Covariates 34

Comparison of meta-analysis to mega-analysis 35

Tractor GWAS analysis 35

QC of summary statistics 36

Low frequency variants in cases 36

Supplementary Figure 16 | Lambda by case allele count. The median lambda across all binary traits by minimum allele count in cases is shown by population. 36

Variants with discrepant frequency compared to gnomAD 36

Supplementary Figure 17 | UKB and gnomAD frequencies. The frequencies in the AFR population in UKB and gnomAD are highly correlated (a), but many variants are discordant, especially at higher frequencies in UKB (b). These variants tend to fail quality filters in gnomAD. A similar pattern is observed for all populations overlapping between UKB and gnomAD (AFR, AMR, EAS, and EUR). 37

Supplementary Figure 18 | Ti/Tv ratio of discrepant variants. Variants that are discordant between UKB and gnomAD have lower Ti/Tv ratios. Points are colored by population (a: AFR, b: AMR, c: EAS, d: EUR) and sized proportional to the number of variants in the bin. Shaded region corresponds to variants that are "well-calibrated" (frequency within 2-fold of gnomAD) and thus retained for downstream analysis. 38

Variants missing from gnomAD 38

Supplementary Figure 19 | Variants removed by gnomAD filters. The number of variants that are well-calibrated (within 2X frequency) are compared to those missing from gnomAD, found in gnomAD but in a different population, or having a significantly different frequency from gnomAD. These metrics are broken down by UKB frequency within EUR (a) and by population (b). 39

LD matrices and scores 40
Supplementary Figure 20 | Pairwise comparisons of LD scores in UKB vs. gnomAD within each genetic ancestry group. Hapmap 3 SNPs are shown for (a) AFR, (b) AMR, (c) EAS, (d) EUR (compared to gnomAD NFE). Dashed line represents y=x. 40
Heritability analysis 41
Supplementary Figure 21 | (a-b) Correlation between UKB round 2 and Pan-UKB EUR LDSC (a) and S-LDSC (b). (c) The number of phenotypes by genetic ancestry group, shaded by significant heritability z scores (S-LDSC h2 z ≥ 4). 41
Supplementary Figure 22 | Characterization of RHEmc run-to-run variability. The first five phenotypes in the manifest are shown. Bars indicate empirical standard deviations (standard deviation of heritability estimates from 50 identical runs of heritability computation) normalized by the standard error of the heritability estimator for each phenotype. Missing points indicate failed convergence. Colors correspond to number of random vectors, indicating that variability goes down as the number of random vectors increases. We chose 50 random vectors for downstream analysis. 43
Supplementary Table 9 | 66 pilot phenotypes chosen for heritability analysis using multiple methods. In the phenotype manifest, phenocode 20002’s description is “Non-cancer illness code, self-reported” and here, the coding description is shown instead. The “note” column refers to the phenotype coding from UK Biobank, except in the cases of “irnt” which denotes that the phenotype was inverse rank normal transformed (typically noted in the “modifier” column of the manifest and release files). 44
Supplementary Figure 23 | Correlation between RHE-mc heritability point estimates (liability scale) and point estimates made in a previous round of heritability analysis restricted to the White British subset of UKB (Round 2) for the same pilot phenotypes. Color represents trait type, dotted line is y=x, error bars are +/- 1se. 45
Supplementary Figure 24 | Cross-method comparison of selected continuous phenotypes. Error bars represent +/- 1se. Only ancestry-trait pairs passing QC were included in this figure. RHEmc 25 bin (and 25 bin, 50 random vectors [RV]) was not run for EUR due to computational limitations. 46
Supplementary Figure 25 | Overview of heritability z scores across trait types and populations. (a) The number of traits passing in each ancestry as a function of h2 z score cutoff (S-LDSC for EUR, RHEmc [25 bins] for all other ancestries). (b) The number of traits passing in 1, 2, 3, 4, 5, or all 6 ancestries (colors) as a function of the z score cutoff. The ancestry-trait pairs used in this plot are pre sumstats QC. S-LDSC -derived z scores reported for EUR, RHEmc (25 bins) reported for all other phenotype-ancestry pairs. 47
Supplementary Figure 26 | Example of a QC-fail GWAS of categorical phenotype 3446 in the AMR genetic ancestry group, “type of tobacco currently smoked”, for category “Manufactured cigarettes” shown as a Manhattan plot (left) and a QQ plot (right). 47
Supplementary Figure 27 | Empirical summary statistics quality control approach. (a) Flowchart of QC approach with each filter used (left) as well as the number of phenotype-ancestry pairs passing each filter. Note that filters are applied sequentially in the listed order. The “heritability within bounds for all ancestries” and “lambda GC > 0.9 for all ancestries” fail for all ancestries if a single ancestry fails the respective filter. “S-LDSC ratio < 0.3 or ratio z score < 4 in all of EUR, CSA, or AFR” fail for all ancestry if any of EUR, CSA, AFR fail, but fail for the individual ancestry-trait pair only if the filter fails for a different ancestry group. (b-c) The distribution of lambda GC (b) and S-LDSC ratio (c) values by genetic ancestry group. Phenotypes that fail the S-LDSC ratio (referred to as “Controlled S-LDSC ratio” in Figure 2c) are highlighted in red. 49
Supplementary Figure 28 | Number of ancestry-trait pairs per trait type passing the z score >= 4

filter as a function of (1) EUR S-LDSC z >= 4, and (2) the total number of ancestry groups passing this filter, shown cumulatively. A greater proportion of the bar colored dark indicates a greater proportion of ancestry-trait pairs passing z >= 4 in a given number of ancestries also passed z >= 4 in EUR. 50

Supplementary Figure 29 | Number of phenotypes passing final quality control steps by combination of genetic ancestry groups in which the phenotype passes. 51

Maximal independent set 52

Supplementary Figure 30 | Distribution of pairwise phenotype correlations across all individuals for filtered high-quality phenotypes, for all correlations (a), and zoomed to a correlation threshold of r2 = 0.1 (b), which was selected to prune for independent phenotypes. 52

Locus definition within and across populations 53

Meta-analysis 54

Polygenicity 55

Supplementary Figure 31 | Polygenicity estimates across trait types. A histogram of polygenicity estimates (the proportion of SNPs with nonzero effects) using SBayesS for 392 phenotypes in EUR. 55

Summary statistics analysis 56

Consistency in summary statistics 56

Supplementary Figure 32 | Consistency of effects across ancestry groups. (a) As in Figure 3c, P-values from meta-analysis versus EUR GWAS alone, colored by p-value of heterogeneity among genetic ancestry groups. (b) For associations that are significant in more than one ancestry group, the majority of betas are positively correlated. 56

Known versus novel association comparisons 57

Supplementary Table 10 | EFO annotation summary. The number of traits mapping to EFO terms and categories is shown by trait type. The final column indicates traits that map to multiple categories. 58

Gene list analysis 59

Supplementary Figure 33 | Percentage of gene lists with at least one significant association. As in Fig. 4b, but all discovered associations rather than restricted to novel associations. 59

Ancestry-enriched associations 60

Supplementary Figure 34 | Forest plot showing association beta for each phenotype for rs1050828 across all available population groups. Error bars correspond to 95% confidence intervals. Abbreviations are defined in Supplementary Table 11. 60

Supplementary Table 11 | Top 5 phenotypes associated with SNP rs1050828 at gene G6PD. * indicates associations passing GWAS significance threshold 5 x 10-8. This variant is low frequency in CSA and EAS and thus, GWAS was not run in these groups. 61

Fine-mapping 62

Comparison between Tractor and SAIGE results 62

Supplementary Datasets 64

Supplementary Dataset 1 | Assigned genetic ancestry labels correlate with the country of birth or known migration events. The number of individuals by genetic ancestry and country of birth (non-UK) are shown. 64

Supplementary Dataset 2 | Summary of all phenotypes in Pan-UKB. Phenotypes are keyed by five keys: trait type, phenocode, pheno_sex, coding, and modifier. Where available, description and coding_description are provided from the UK Biobank showcase. For each ancestry group, we include the number of cases, heritability estimates (observed, liability, standard errors, and z

scores), whether the phenotype passes QC, and lambda GC. We provide QC flags, whether the phenotype is in the maximal independent set, and filename information, including a download link for the phenotype-specific file and tabix index on Amazon S3 and md5 checksums for each. 64
Supplementary Dataset 3 | Summary of all heritability metrics. Phenotypes are keyed as in Supplementary Dataset 2. For each ancestry group, we provide heritability estimates (observed, liability, standard errors, and z scores) for LDSC and S-LDSC, and for ancestry groups other than EUR, also RHE-mc, as well as details of QC flags. 64
Supplementary Dataset 4 | Pairwise genetic correlations. Genetic correlations (rg) from S-LDSC are computed for pairs of 528 phenotypes (phenotype_code_1 and phenotype_code_2), using summary statistics from EUR. 64
Supplementary Dataset 5 | Pairwise phenotypic correlations. Covariates were regressed out from each of the 452 high-quality phenotypes, and pairwise correlations (entry) were computed for each pair of phenotypes (residuals), i (with phenotype identifier in i_data) and j (identifier in j_data). The correlation for all phenotypes is available at gs://ukb-diverse-pops-public/misc/pairwise/pairwise_correlations_regressed.txt.bgz 64
Supplementary Dataset 6 | Polygenicity estimates. Polygenicity estimates (mean and standard deviation) from SBayesS for 451 phenotypes, along with convergence criteria (R_GelmanRubin). 64
Supplementary Dataset 7 | Summary statistics for key loci across GWAS methods. SAIGE AFR and SAIGE EUR refer to the SAIGE analyses performed on the African (AFR) and European (EUR) genetically inferred ancestry groups of UKB. Tractor AFR and Tractor EUR indicate the Tractor GWAS conducted on the African or European haplotype tracts, respectively, within the AFR group. Variants are filtered as described above in Tractor GWAS analysis. 64
**FAQ for diverse ancestry GWAS** **66**
Background 67
Who conducted this study? 67
What are the group’s overarching goals? 68
Why was this study done? 68
What is ancestry? Is it the same as race or ethnicity? 69
In this study, you perform many GWAS for many phenotypes. What is a GWAS? What is a phenotype? 70
What does it mean for a variant to be associated with a phenotype? Are the genetic variants discovered by GWAS “causal”? 70
Do these results imply that genetics are responsible for the phenotypic differences between ancestry groups? 71
Since biology is mostly shared, why is diversity in genetics so important? 72
What did you learn as part of this study? 72
Study design 73
What was done? 73
What data were used? 74
Have you used data from countries other than the UK? 74
How were participants recruited? 74
How did you decide which phenotypes to study? 75
How did you decide what ancestry groups to include? How did you assign individuals to each ancestry group? 75

What about people with mixed ancestries? 76
Why do you analyze ancestry groups separately? 77
Why have certain individuals been excluded in previous research? 78
Why are you including them now? 78
Social and Ethical Implications 79
What can be done with the results of this research? What are the potential benefits of this research? 79
Do you study the genetics of behavior? 80
Do genes determine the choices we make? 80
Are there policy or clinical implications for this research? 81
How has genetics research been used in the past to harm different groups? 82
Could this research be used to harm certain groups (e.g., through discrimination or stigmatization)? 83
What has been done to reduce the potential harms of this research? 83
References 85

### Ancestry analysis and relatedness inference

Controlling for population structure is a critical step for robustness in GWAS, specifically to avoid identifying spurious false associations with traits and diseases[1]. When associating genetic variants with a trait of interest, population structure is typically included as a covariate using principal components (PCs) that quantify genetic ancestry. “Genetic ancestry” is a statistical construct based on the genetic variants that an individual inherited from their ancestors. An individual’s self-identified race or ethnicity may at times differ from the corresponding genetic ancestry assigned by statistical algorithms. Treating ancestry, ethnicity, and race as equivalent concepts is incorrect. In all our analyses, we exclusively refer to genetic ancestry. Please refer to the FAQ at the end of this document for an in-depth discussion of these concepts as they pertain to our work.

To minimize false positive rates in GWAS analyses that arise due to confounding with population structure (see Association Analysis below), we used a three-stage approach to analyze genetic ancestry. Specifically, we: 1) assigned ancestry labels for within-group analysis using reference panel meta-data, 2) visualized population structure within subcontinental ancestries alongside reference panel data, and 3) pruned ancestry outliers within assigned population labels. This approach was intended to ensure that across a breadth of phenotypes, GWAS were as inclusive as possible across ancestry groups while balancing the removal of ancestry outliers so that GWAS summary statistics results were not inflated by population stratification introduced by these outliers.

#### Initial ancestry assignment

For the initial step of assigning ancestry labels to UKB participants, we first combined and harmonized reference panel data from phase 3 of the 1000 Genomes Project[2] with Human Genome Diversity Project (HGDP) samples genotyped on the Illumina 650k array and lifted over to hg19[3]. Briefly, we combined these reference datasets into continental ancestries according to their corresponding meta-data as shown in **Supplementary Table 1**. We filtered to keep SNPs with MAF > 0.5% and missingness < 5% using PLINK [4], resulting in 639,590 variants. Within continental ancestries, we then removed reference panel individuals determined to be 2nd degree relatives or closer using KING 2.0[5], resulting in 3,295 individuals.

We then intersected these reference panel data with UKB genotyped sites on both UK Biobank Axiom and UK BiLEVE Axiom arrays[6], resulting in a set of 64,233 SNPs. We ran PCA on unrelated individuals from the 1000 Genomes Project and HGDP reference data as defined above. To partition individuals in the UKB into more ancestrally homogeneous groups roughly corresponding to continental ancestry, we used the PC loadings from the reference dataset to project UKB individuals into the same PC space. We trained a random forest classifier using the sklearn package implemented in a custom python script using harmonized continental ancestry meta-data labels from the reference HGDP and 1000 Genomes Project training data. Because we have a multi-stage ancestry assignment approach, our initial assignments were intended to be permissive, specifically with: a) how strict of a random forest probability cutoff to use, and b) the number of PCs to use in the random forest training model. We first used a random forest with 6 PCs to distinguish the 7 project labels harmonized in **Supplementary Table 1**. We compared ancestry assignments with $p > 0.5$, $p > 0.6$, $p > 0.7$, $p > 0.8$, and $p > 0.9$, which showed the broadly expected trends of increasing numbers of individuals assigned to “Other” (**Supplementary Table 2**). Given our subsequent filtering steps based on ancestry outliers, we chose to be permissive with this initial filter and ultimately used the random forest $p > 0.5$ model. We next compared ancestry assignments using random forests based on 6 versus 20 PCs (**Supplementary Table 3)**. Results were overall highly concordant with the primary difference being that more individuals assigned the EUR label with the 6 PC model were assigned “Other” with the 20 PCs model. We therefore used the model based on the first 6 PCs. If individuals were not assigned with $p > 0.5$ using the 6 PC random forest model, they were dropped from subsequent analysis. PCs and classifications are shown in **Supplementary Fig. 1**. Using these assignments, we next determined relatedness within each population. Specifically, we ran PC-Relate implemented in Hail with k=10 and min_individual_maf=0.05. To get the maximal set of unrelated individuals, we then ran hl.maximal_independent_set(). Counts of individuals through stages of quality control by population including stages that are described in Pruning ancestry outliers are shown in **Supplementary Table 4**.

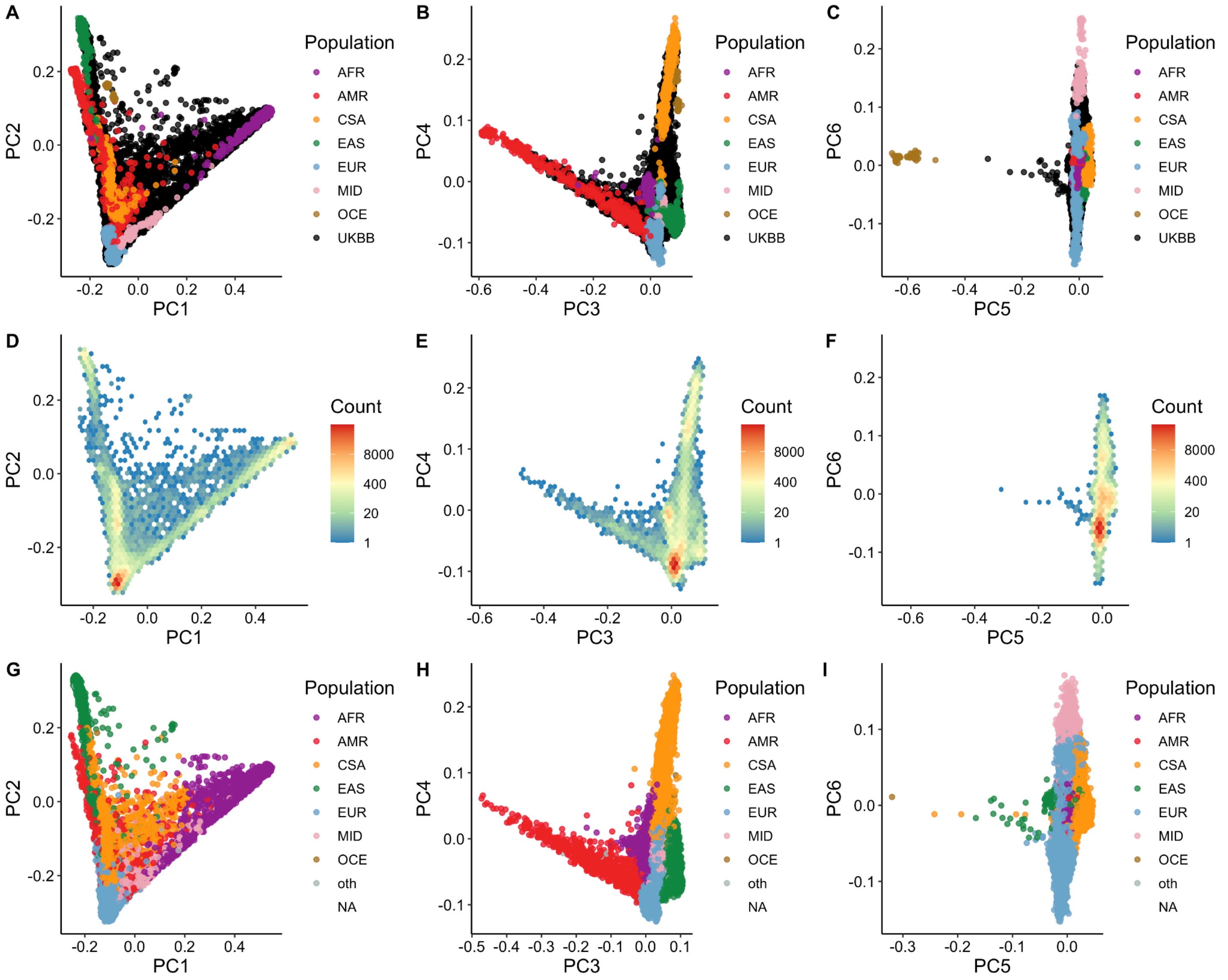


**Supplementary Figure 1 | Genetic ancestry assignment using principal components analysis (PCA).** A-C) First 6 PCA biplots for UKB individuals in black with reference populations from the 1000 Genomes Project and HGDP on top colored by continental ancestry. D-F) Density of UKB participant global PCs. G-I) Genetic ancestry assignments for UKB participants based on reference panel meta-data labels. No reference data are included in these plots. A, D, and G) PCA biplots for PCs 1-2; B, E, and H) PCs 3-4; C, F, and I) PCs 5-6. 3-letter continental ancestry codes are as in **Supplementary Table 1**.

**Supplementary Table 1 | Genetic reference panel data from the 1000 Genomes Project (1KG) and Human Genome Diversity Project (HGDP).** Continental ancestries are EUR=European, CSA=Central/South Asian, AFR=African, AMR=Admixed American, EAS=East Asian, MID=Middle Eastern, and OCE=Oceanian. Population abbreviations with three-letter codes are the same as in the 1000 Genomes Project.

| Project | Cont. ancestry | Population | N | Project | Cont. ancestry | Population | N | Project | Cont. ancestry | Population | N |
|---|---|---|---|---|---|---|---|---|---|---|---|
| 1KG | AFR | ACB | 96 | HGDP | CSA | Hazara | 24 | 1KG | EUR | IBS | 107 |
| 1KG | AFR | ASW | 61 | HGDP | CSA | Kalash | 25 | 1KG | EUR | TSI | 107 |
| 1KG | AFR | ESN | 99 | HGDP | CSA | Makrani | 25 | HGDP | EUR | Adygei | 17 |
| 1KG | AFR | GWD | 113 | HGDP | CSA | Pathan | 23 | HGDP | EUR | Basque | 24 |
| 1KG | AFR | LWK | 99 | HGDP | CSA | Sindhi | 25 | HGDP | EUR | French | 29 |
| 1KG | AFR | MSL | 85 | 1KG | EAS | CDX | 93 | HGDP | EUR | Italian | 13 |
| 1KG | AFR | YRI | 108 | 1KG | EAS | CHB | 103 | HGDP | EUR | Orcadian | 16 |
| HGDP | AFR | BantuKenya | 12 | 1KG | EAS | CHS | 105 | HGDP | EUR | Russian | 25 |
| HGDP | AFR | BantuSAfrica | 8 | 1KG | EAS | JPT | 104 | HGDP | EUR | Sardanian | 28 |
| HGDP | AFR | BiakaPygmy | 32 | 1KG | EAS | KHV | 99 | HGDP | EUR | Tuscan | 8 |
| HGDP | AFR | Mandenka | 24 | HGDP | EAS | Cambodian | 11 | HGDP | MID | Bedouin | 48 |
| HGDP | AFR | MbutiPygmy | 15 | HGDP | EAS | Dai | 10 | HGDP | MID | Druze | 47 |
| HGDP | AFR | San | 6 | HGDP | EAS | Daur | 9 | HGDP | MID | Mozabite | 30 |
| HGDP | AFR | Yoruba | 24 | HGDP | EAS | Han | 44 | HGDP | MID | Palestinian | 51 |
| 1KG | AMR | CLM | 94 | HGDP | EAS | Hezhen | 9 | HGDP | OCE | Melanesian | 19 |
| 1KG | AMR | MXL | 64 | HGDP | EAS | Japanese | 29 | HGDP | OCE | Papuan | 17 |
| 1KG | AMR | PEL | 85 | HGDP | EAS | Lahu | 10 | | | | |
| 1KG | AMR | PUR | 104 | HGDP | EAS | Miaozu | 10 | | | | |
| HGDP | AMR | Colombian | 13 | HGDP | EAS | Mongola | 10 | | | | |
| HGDP | AMR | Karitiana | 24 | HGDP | EAS | Naxi | 9 | | | | |
| HGDP | AMR | Maya | 25 | HGDP | EAS | Oroqen | 10 | | | | |
| HGDP | AMR | Pima | 25 | HGDP | EAS | She | 10 | | | | |
| HGDP | AMR | Surui | 21 | HGDP | EAS | Tu | 10 | | | | |
| 1KG | CSA | BEB | 86 | HGDP | EAS | Tujia | 10 | | | | |
| 1KG | CSA | GIH | 103 | HGDP | EAS | Uygur | 10 | | | | |
| 1KG | CSA | ITU | 102 | HGDP | EAS | Xibo | 9 | | | | |
| 1KG | CSA | PJL | 96 | HGDP | EAS | Yakut | 25 | | | | |
| 1KG | CSA | STU | 102 | HGDP | EAS | Yizu | 10 | | | | |
| HGDP | CSA | Balochi | 25 | 1KG | EUR | CEU | 99 | | | | |
| HGDP | CSA | Brahui | 25 | 1KG | EUR | FIN | 99 | | | | |
| HGDP | CSA | Burusho | 25 | 1KG | EUR | GBR | 91 | | | | |

**Supplementary Table 2 | Comparison of ancestry assignments based on random forests with 6 principal components and a range of probability thresholds.**

| | Count | | | | |
|---|---|---|---|---|---|
| **Super population** | RF p>0.9 | RF p>0.8 | RF p>0.7 | RF p>0.6 | RF p>0.5 |
| **EUR** | 443498 | 452647 | 455444 | 458004 | 459874 |
| **CSA** | 10759 | 10950 | 11013 | 11081 | 11124 |
| **AFR** | 8892 | 9072 | 9129 | 9191 | 9226 |
| **AMR** | 1096 | 1123 | 1133 | 1141 | 1152 |
| **EAS** | 2799 | 2849 | 2879 | 2905 | 2918 |
| **MID** | 1608 | 1639 | 1651 | 1658 | 1667 |
| **OCE** | 2 | 2 | 2 | 2 | 2 |
| **Other (< prob)** | 19723 | 10095 | 7126 | 4395 | 2414 |
| **TOTAL** | 488377 | 488377 | 488377 | 488377 | 488377 |

**Supplementary Table 3 | Comparison between ancestry assignments using a random forest trained on 6 principal components (PCs) versus 20 PCs.** Assignments based on 6 PCs are shown in rows, while assignments based on 20 PCs are shown in columns. More individuals are assigned to “Other” using the random forest based on 20 PCs.

| | **AFR** | **AMR** | **CSA** | **EAS** | **EUR** | **MID** | **OCE** | **Other** |
|---|---|---|---|---|---|---|---|---|
| **AFR** | 9045 | 101 | 0 | 0 | 24 | 0 | 0 | 56 |
| **AMR** | 5 | 916 | 0 | 0 | 221 | 0 | 0 | 10 |
| **CSA** | 25 | 24 | 10794 | 17 | 203 | 6 | 0 | 55 |
| **EAS** | 3 | 31 | 26 | 2745 | 89 | 1 | 0 | 23 |
| **EUR** | 2 | 29 | 233 | 1 | 457006 | 189 | 0 | 2414 |
| **MID** | 132 | 136 | 4 | 0 | 139 | 1250 | 0 | 6 |
| **OCE** | 0 | 0 | 0 | 0 | 0 | 0 | 2 | 0 |
| **Other** | 11 | 1 | 7 | 3 | 456 | 4 | 0 | 1932 |

**Supplementary Table 4 | Counts of individuals by continental ancestry as inferred from the 1000 Genomes and Human Genome Diversity Panel genetic data**. Oceanians were removed from further analyses given the very small sample sizes. Those individuals whose ancestry groups could not be assigned (i.e., “Other”) were also removed. The number of outliers removed indicates the count remaining after pruning ancestry outliers according to PC coordinates. 3-letter continental ancestry codes are as in **Supplementary Table 1**.

| **Continent:** | **AFR** | **AMR** | **CSA** | **EAS** | **EUR** | **MID** | **OCE** | **Other** |
|---|---|---|---|---|---|---|---|---|
| Count | 9,226 | 1,152 | 11,124 | 2,918 | 459,874 | 1,667 | 2 | 2,414 |
| Outliers removed (total) | 6,806 | 998 | 9,109 | 2,783 | 426,936 | 1,624 | N/A | N/A |
| Outliers removed (unrelated) | 6,259 | 991 | 8,286 | 2,701 | 362,558 | 1,568 | N/A | N/A |

#### Visualizing subcontinental ancestries

In the second step of our ancestry analyses outlined above, our goal was to visualize population structure at the subcontinental level. We ran PCA on the HGDP and 1000 Genomes Project reference panel genotypes within each meta-data label group, including AFR, AMR, CSA, EAS, EUR, and MID, then projected the UKB participants assigned to the corresponding ancestry group onto the same PC space (**Supplementary Fig. 2-8**). Because of the high amount of genetic diversity in African ancestry populations and our additional access to genetic data from the African Genome Variation Project genotyped on the Illumina Omni2.5 array[7], we merged these data with the 1000 Genomes Project and HGDP data for finer-scale analysis of individuals assigned to AFR specifically. We used this supplemented reference panel for PCA (**Supplementary Table 5**), then projected UKB participants assigned to AFR into the same PC space (**Supplementary Fig. 2**).

As shown in **Supplementary Fig. 2** and described more fully previously[8,9], most of the UKB participants assigned to AFR cluster most closely with West African reference panel populations and/or are along a cline on PC1 that corresponds with West African and European admixture. Based on ancestry proportions in the admixed ACB population in the 1000 Genomes Project, a sizable fraction of individuals likely have first generation admixture between parents from West African and European populations. A much smaller number of UKB individuals assigned to AFR have ancestry clustering with East and southern African reference populations, as described in more detail in Majara et al[8]. The relatively few UKB individuals assigned to AMR have ancestry spanning the full set of AMR individuals in the reference panel with the exception of the Pima, consistent with variable recent admixture proportions found in Hispanic/Latino populations (**Supplementary Fig. 3**). The UKB individuals assigned to CSA have ancestry spanning a range of individuals particularly from India, Pakistan, and to a lesser extent Bangladesh in the reference panel (**Supplementary Fig. 4**), with few to no individuals clustering with the Kalash, Makrani, Balochi, Pathan, or ITU reference populations. The UKB individuals assigned to EAS have variable ancestries with most clustering with Chinese and Japanese reference populations, but a sizable number that do not cluster closely with any populations in the reference panel (**Supplementary Fig. 5**). The UKB individuals assigned to EUR span the full breadth of diversity in the reference panel, with the majority clustering with the GBR and CEU populations in the reference panel (**Supplementary Fig. 6**). The MID reference panel is sparse, but most UKB individuals assigned to MID cluster most closely with the Palestinian or along a cline towards the Mozabite (**Supplementary Fig. 7**). All of these subcontinental ancestry trends are in line with expectations from self-reported ethnicity and birth record data described more fully below and shown in **Supplementary Table 6**, **Supplementary Dataset 1**, and **Supplementary Fig. 14**.

**Supplementary Table 5 | Additional genetic reference panel data from the African Genome Variation Project (AGVP) used for visualization of subcontinental PCA within African ancestry assignments (AFR).**

| Population | N | Population | N |
|---|---|---|---|
| Baganda | 97 | Jola | 79 |
| Banyarwanda | 95 | Kalenjin | 100 |
| Barundi | 91 | Kikuyu | 99 |
| Ethiopia | 107 | Mandinka | 87 |
| Fula | 74 | Sotho | 86 |
| Ga-Adangbe | 100 | Wolof | 78 |
| Igbo | 99 | Zulu | 100 |

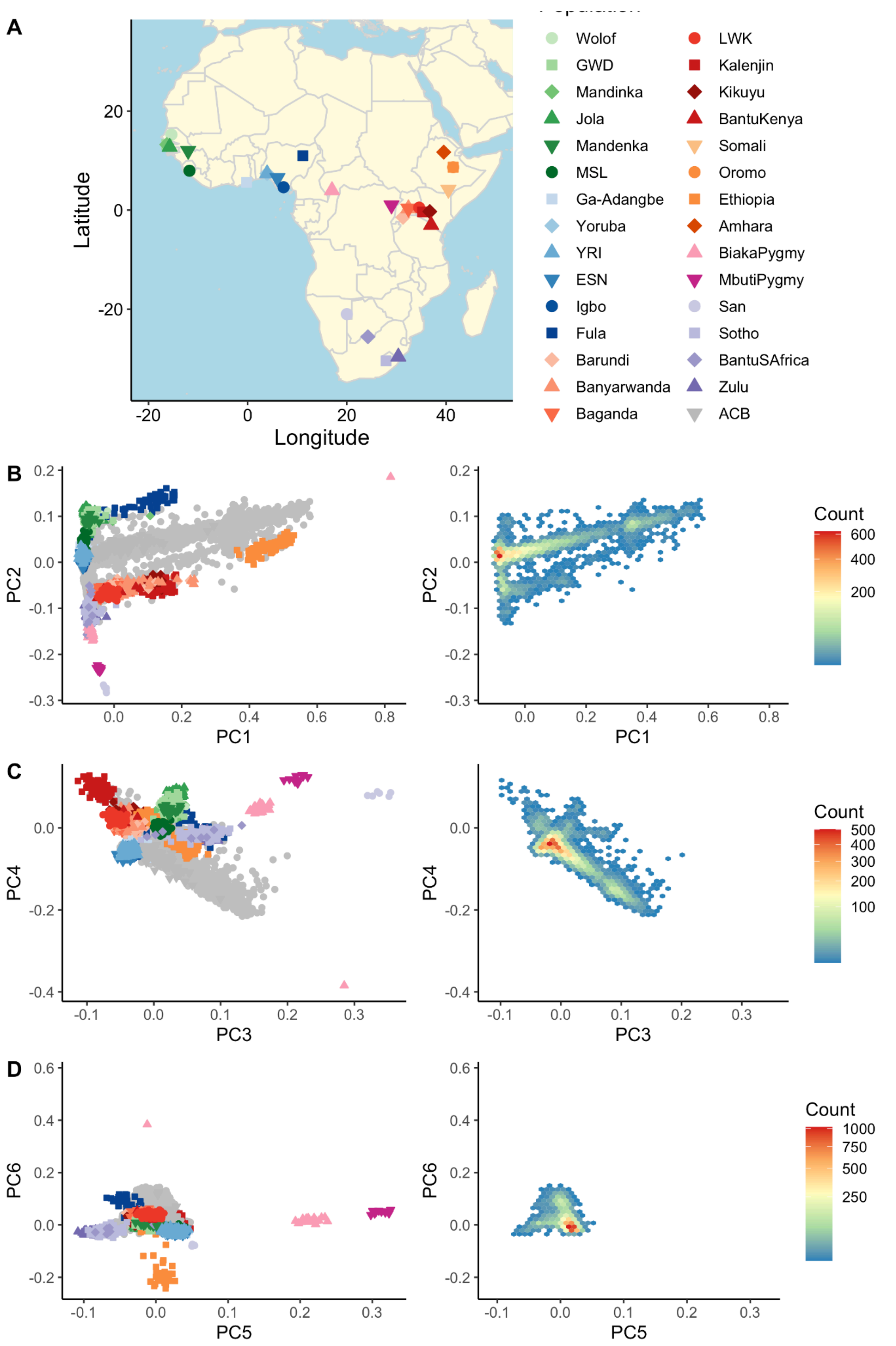


**Supplementary Figure 2 | Subcontinental ancestry PCs in the AFR reference panel and UKB participants assigned to AFR.** UKB participants assigned to AFR are shown in grey, while reference populations are on top and colored as in the map. A) Map of reference populations, B) PCs 1-2, C) PCs 3-4, D) PCs 5-6. Left panels show PC values, right panels show UKB PC densities.

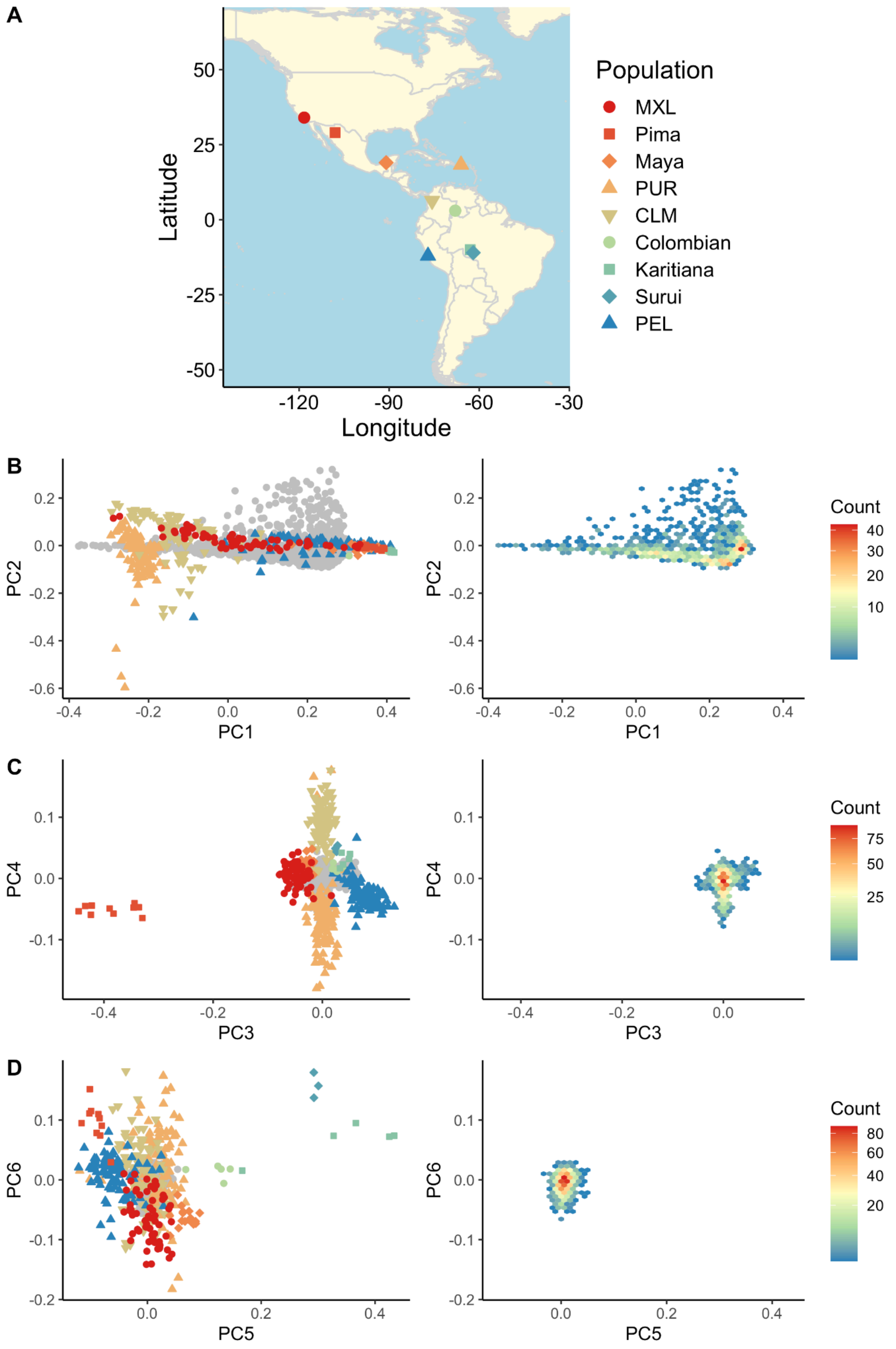


**Supplementary Figure 3 | Subcontinental ancestry PCs in the AMR reference panel and UKB participants assigned to AMR.** UKB participants assigned to AMR are shown in grey, while reference populations are on top and colored as in the map. A) Map of reference populations, B) PCs 1-2, C) PCs 3-4, D) PCs 5-6. Left panels show PC values, right panels show UKB PC densities.

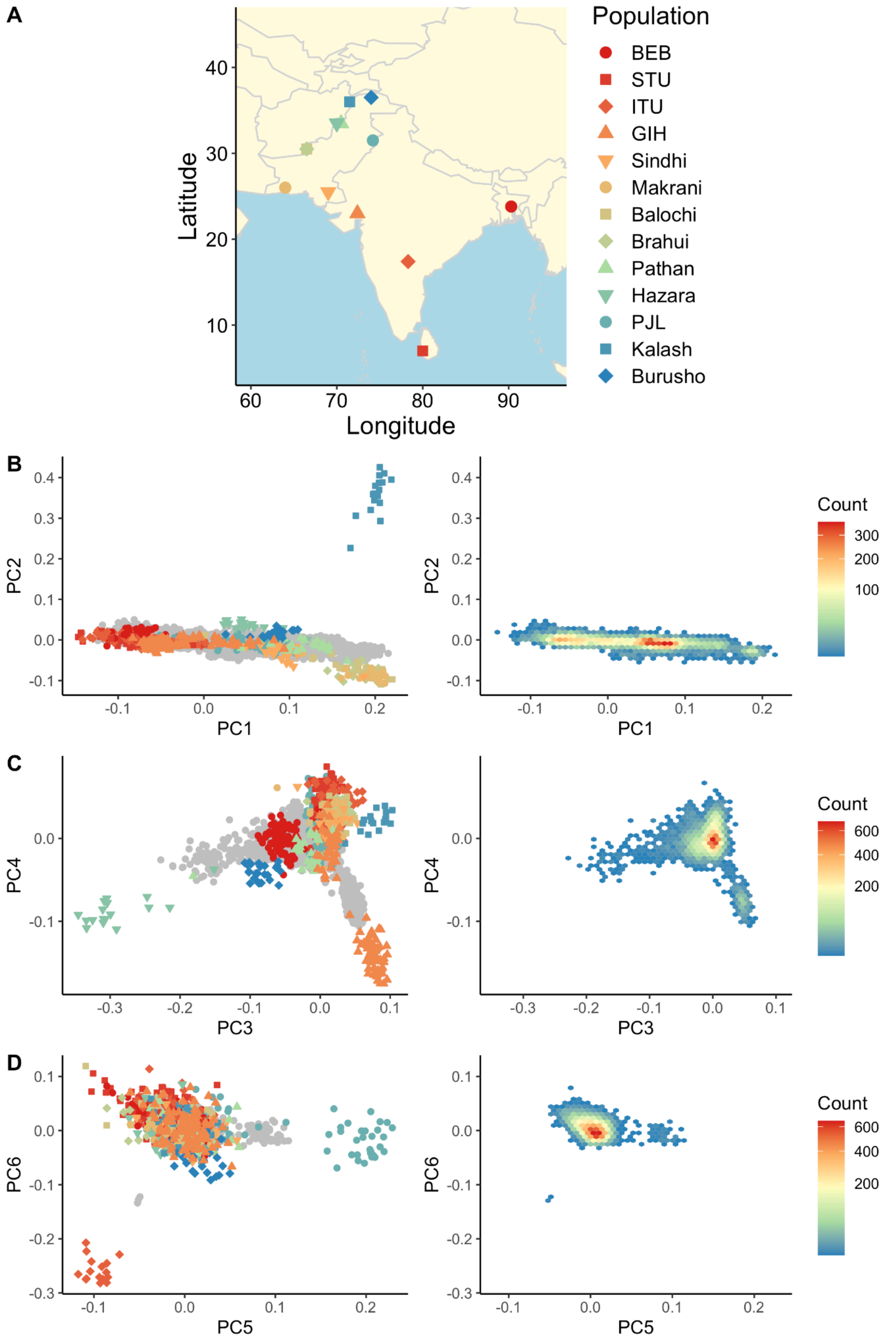


**Supplementary Figure 4 | Subcontinental ancestry PCs in the CSA reference panel and UKB participants assigned to CSA.** UKB participants assigned to CSA are shown in grey, while reference populations are on top and colored as in the map. A) Map of reference populations, B) PCs 1-2, C) PCs 3-4, D) PCs 5-6. Left panels show PC values, right panels show UKB PC densities.

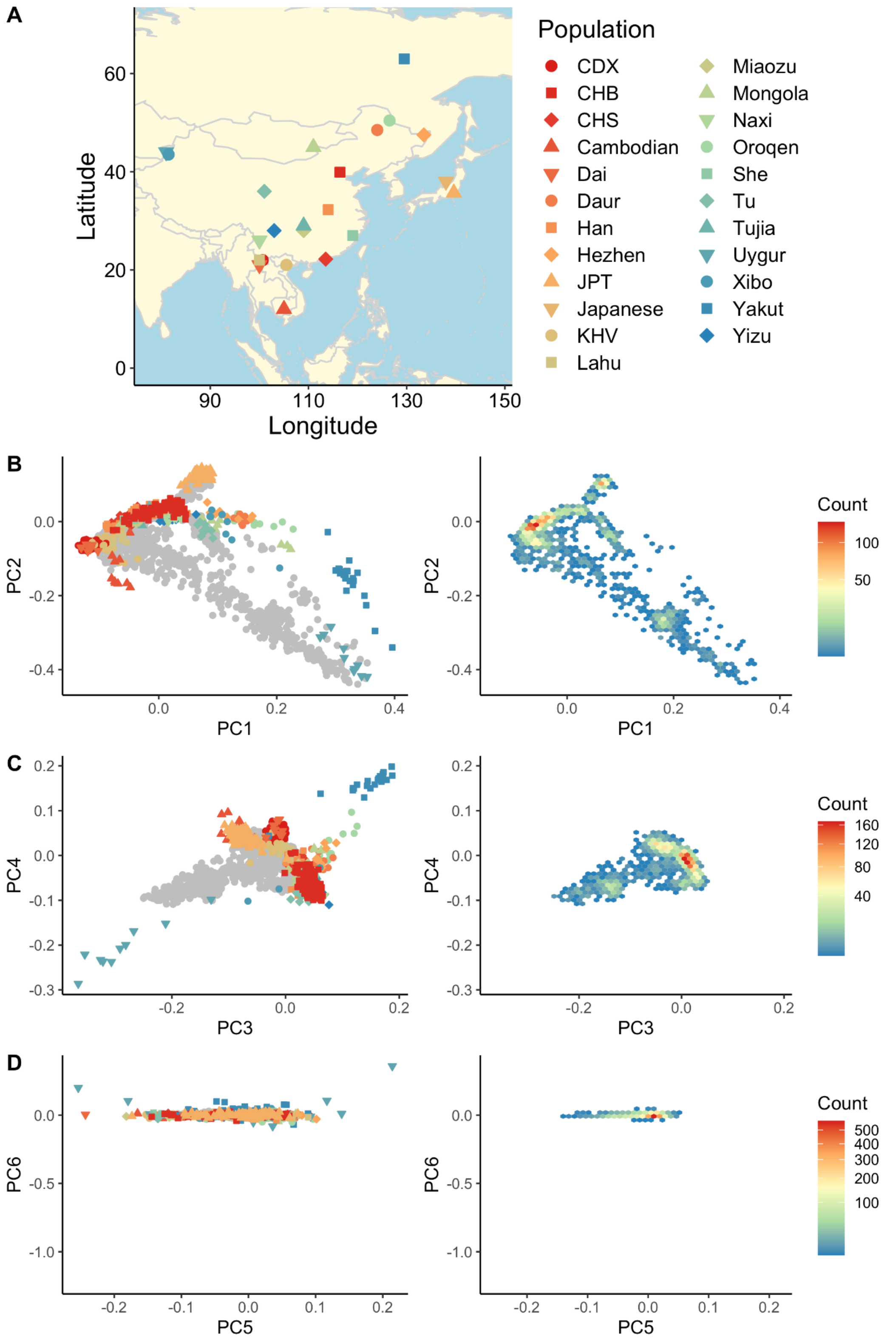


**Supplementary Figure 5 | Subcontinental ancestry PCs in the EAS reference panel and UKB participants assigned to EAS.** UKB participants assigned to EAS are shown in grey, while reference populations are on top and colored as in the map. A) Map of reference populations, B) PCs 1-2, C) PCs 3-4, D) PCs 5-6. Left panels show PC values, right panels show UKB PC densities.

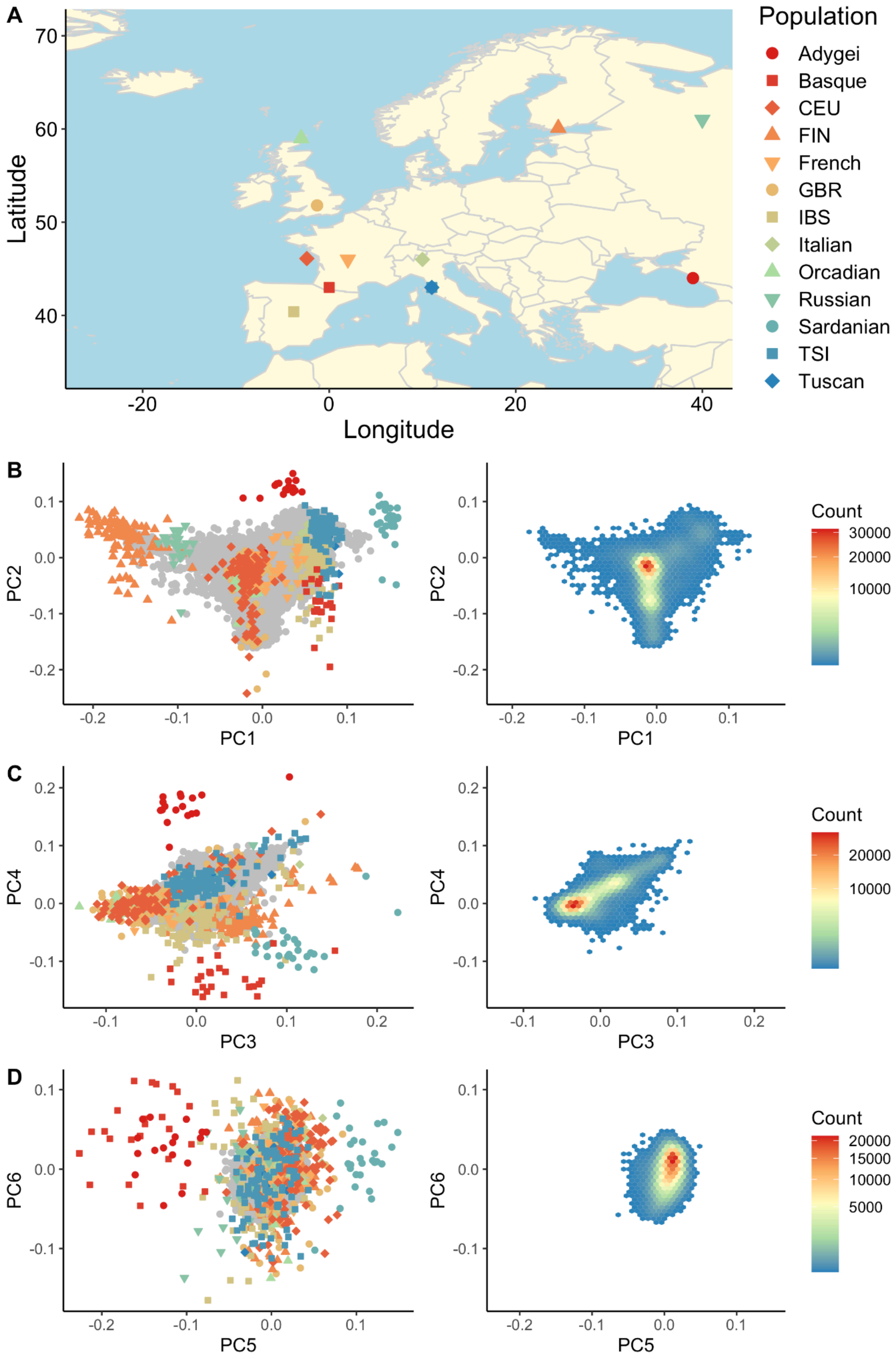


**Supplementary Figure 6 | Subcontinental ancestry PCs in the EUR reference panel and UKB participants assigned to EUR.** UKB participants assigned to EUR are shown in grey, while reference populations are on top and colored as in the map. A) Map of reference populations, B) PCs 1-2, C) PCs 3-4, D) PCs 5-6. Left panels show PC values, right panels show UKB PC densities.

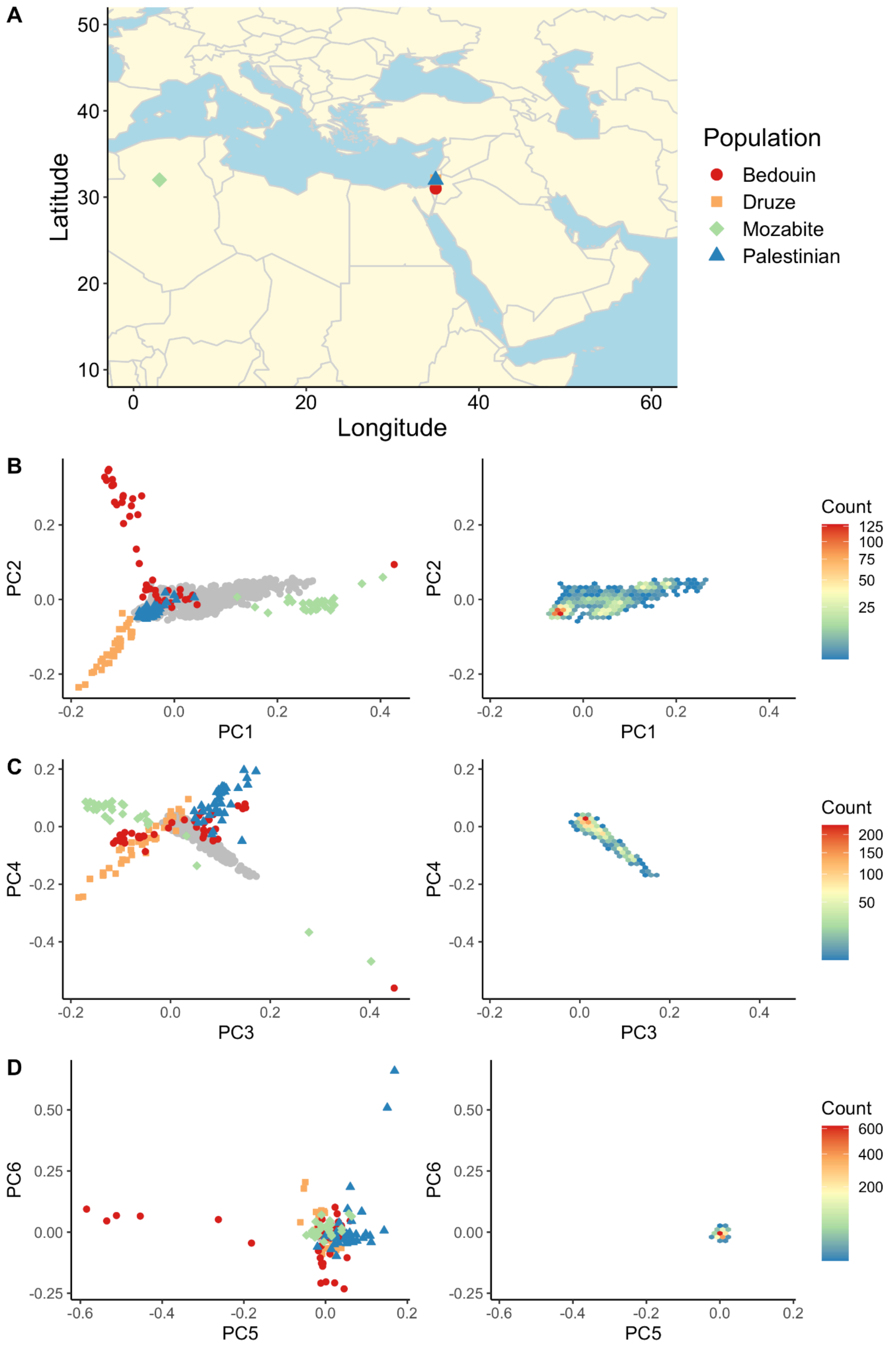


**Supplementary Figure 7 | Subcontinental ancestry PCs in the MID reference panel and UKB participants assigned to MID.** UKB participants assigned to MID are shown in grey, while reference populations are on top and colored as in the map. A) Map of reference populations, B) PCs 1-2, C) PCs 3-4, D) PCs 5-6. Left panels show PC values, right panels show UKB PC densities.

#### Pruning ancestry outliers

To further refine ancestry classifications, in our third stage of ancestry analyses outlined above, our goal was to prune ancestry outliers within assigned labels based on subcontinental structure. We started by rerunning PCA among UKB individuals within each assigned population label (i.e., excluding reference panel data) using individuals determined to be unrelated by PC-Relate (minimum kinship of 0.05). We then projected related individuals into the same PC space for use as covariates in generalized linear mixed models implemented in SAIGE and described in Association analysis. We calculated the total distance from population centroids across 10 PCs. Using the PC scores, we computed centroid distances across 3-5 centroids spanning these PCs depending on the degree of heterogeneity within each continental ancestry as follows:

$$d = \sum_{i=1}^{n} \frac{(X_i - \overline{X}_i)^2}{\sigma^2_{X,i}}$$

Where $d$ is the total centroid distance summed over $n$ total dimensions of an ellipse (i.e., PCs), $X_i$ is a vector of PCs, $\overline{X}_i$ is the mean PC score, and $\sigma^2_{X,i}$ is the variance of the PC scores for the $i^{th}$ PC. We identified ancestry outliers by plotting histograms of centroid distances and removing individuals who had outlying distances at the high end of the distribution (**Supplementary Fig. 8-13**).

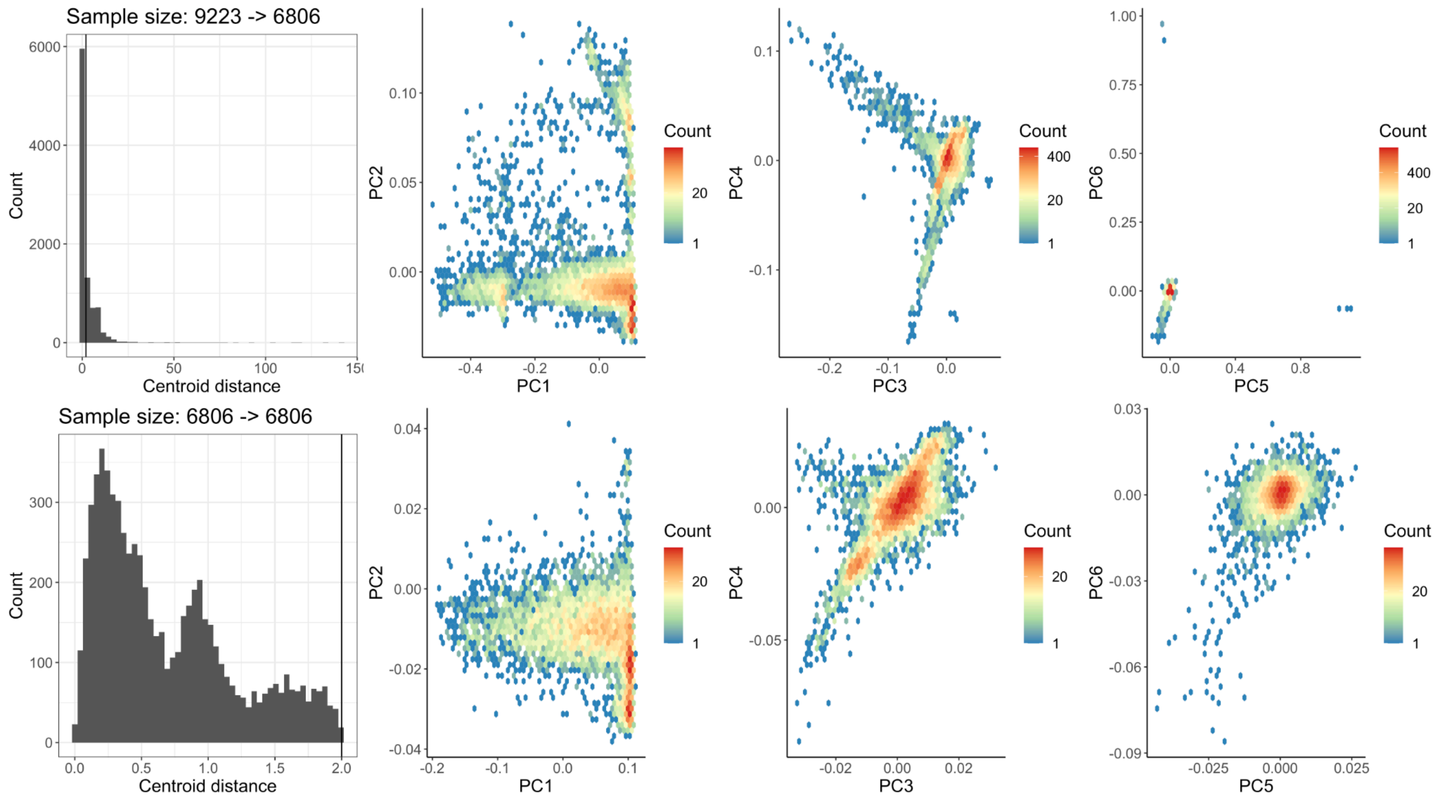


**Supplementary Figure 8 | PCA in UKB participants assigned to AFR and corresponding centroid distance across 3 PCs.** Centroid distance distributions and PC biplots for the first 6 PCs are shown before (top) and after (bottom) pruning outliers. Vertical line in the top left centroid distance histogram shows the threshold chosen to remove outliers.

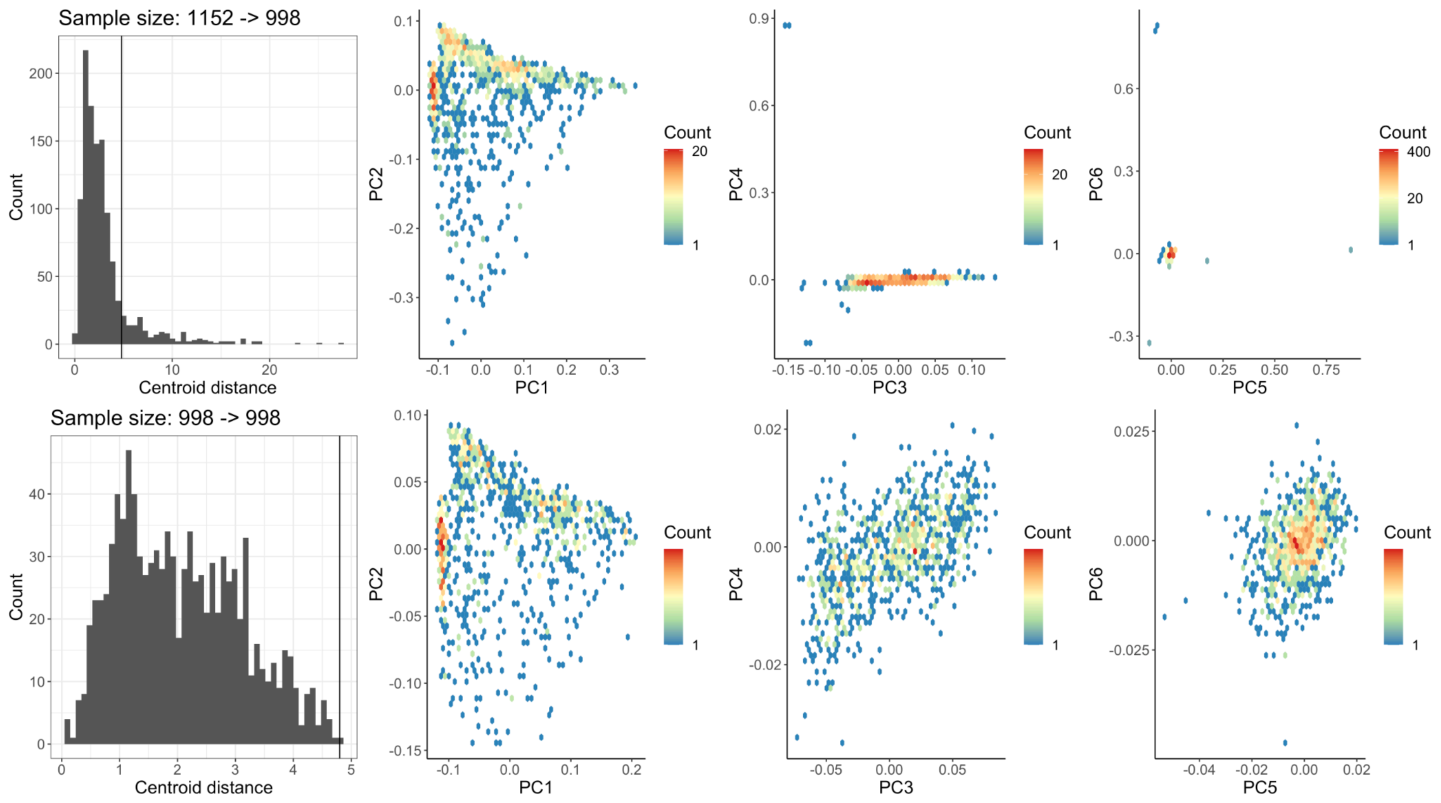


**Supplementary Figure 9 | PCA in UKB participants assigned to AMR and corresponding centroid distance across 3 PCs.** Centroid distance distributions and PC biplots for the first 6 PCs are shown before (top) and after (bottom) pruning outliers. Vertical line in the top left centroid distance histogram shows the threshold chosen to remove outliers.

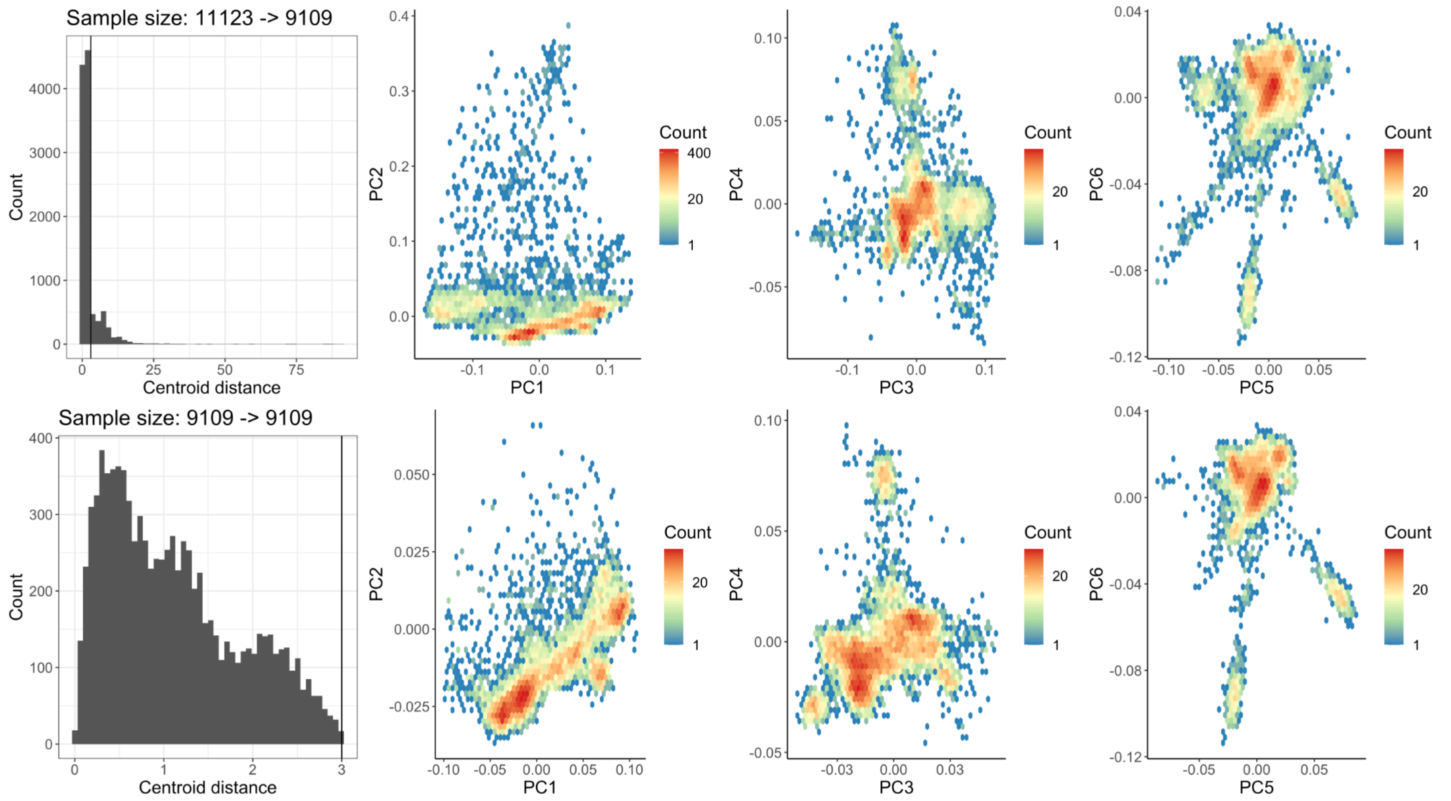


**Supplementary Figure 10 | PCA in UKB participants assigned to CSA and corresponding centroid distance across 3 PCs.** Centroid distance distributions and PC biplots for the first 6 PCs are shown before (top) and after (bottom) pruning outliers. Vertical line in the top left centroid distance histogram shows the threshold chosen to remove outliers.

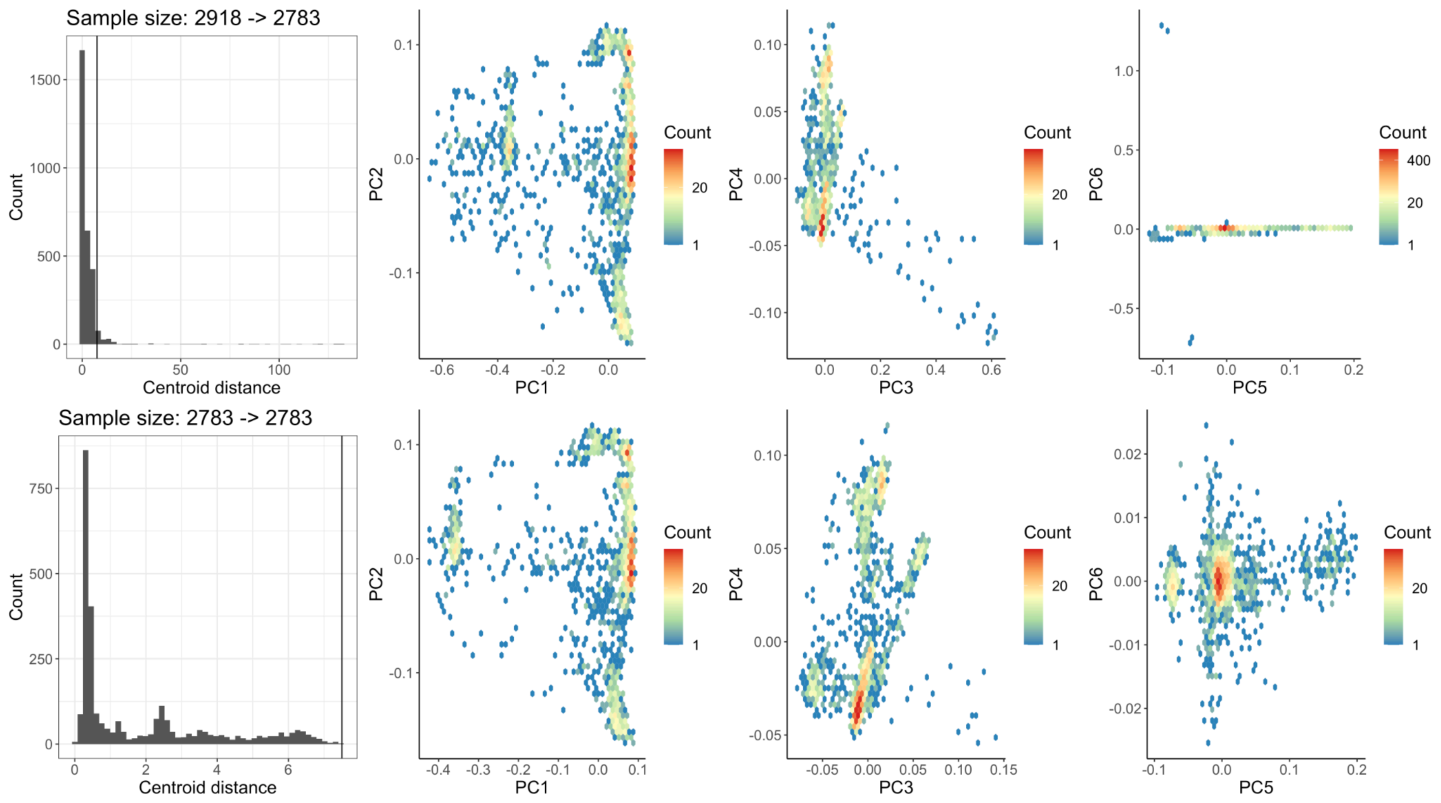


**Supplementary Figure 11 | PCA in UKB participants assigned to EAS and corresponding centroid distance across 3 PCs.** Centroid distance distributions and PC biplots for the first 6 PCs are shown before (top) and after (bottom) pruning outliers. Vertical line in the top left centroid distance histogram shows the threshold chosen to remove outliers.

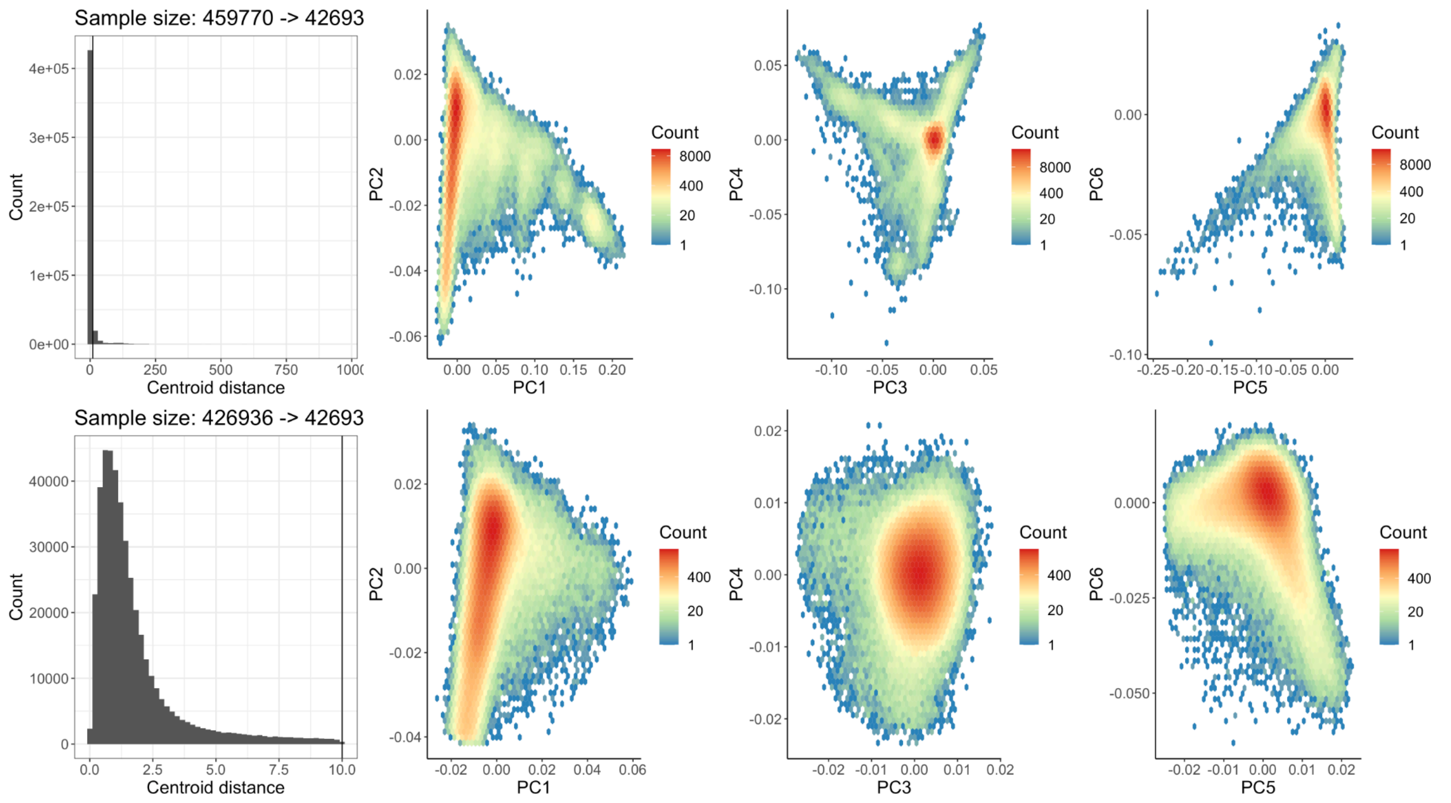


**Supplementary Figure 12 | PCA in UKB participants assigned to EUR and corresponding centroid distance across 5 PCs.** Centroid distance distributions and PC biplots for the first 6 PCs are shown before (top) and after (bottom) pruning outliers. Vertical line in the top left centroid distance histogram shows the threshold chosen to remove outliers.

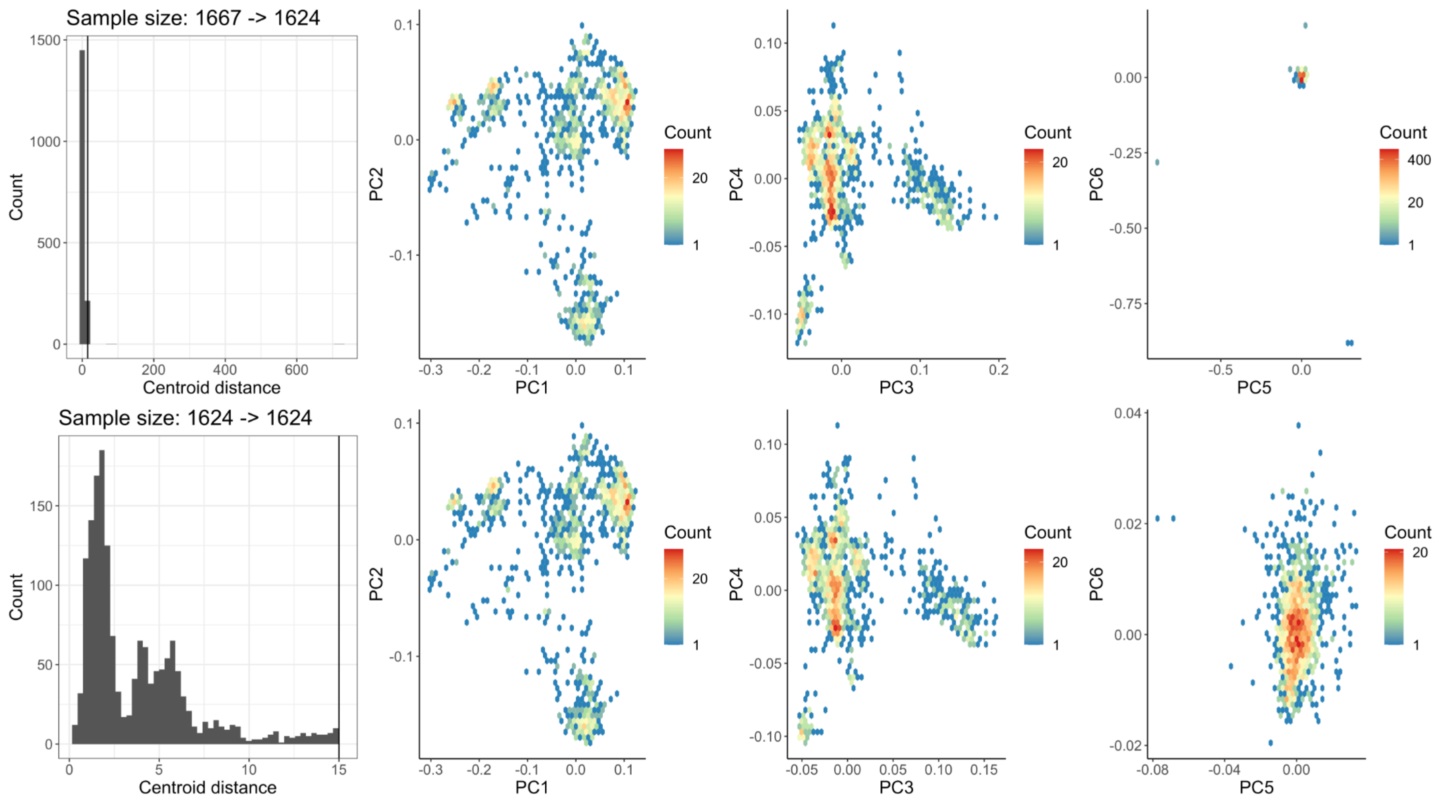


**Supplementary Figure 13 | PCA in UKB participants assigned to MID and corresponding centroid distance across 5 PCs.** Centroid distance distributions and PC biplots for the first 6 PCs are shown before (top) and after (bottom) pruning outliers. Vertical line in the top left centroid distance histogram shows the threshold chosen to remove outliers.

#### Relationship between ancestry and self-reported metrics

To explore the relationships between genetic ancestry labels inferred from population genetic reference panels, self-reported ethnicity (UKB code 21000 “Ethnic background”), and geographic birthplace data for those born outside the UK (UKB code 20115 “Country of Birth (non-UK origin)”), we report the overlap between our ancestry assignments and self-reported ethnicity data (**Supplementary Table 6**), continent (**Supplementary Table 7**) and country of birth (**Supplementary Dataset 1**). Most patterns are expected. For example, for ethnicity, EUR primarily report British, Irish, or Any other white background; CSA primarily report Indian, Pakistani, or Any other Asian background; AFR primarily report African or Caribbean; EAS primarily report Chinese; and AMR and MID primarily report Other ethnic group. Those who prefer not to self-report their ethnic group are depleted for EUR and enriched in CSA and AFR groups. While we have compared genetic ancestry and ethnicity data which show expected trends, these are notably distinct concepts[10,11], with the genetic ancestry labels we have used for GWAS dividing up the ancestry continuum.

Genetic ancestry labels for those born outside of the UK often align with continental birthplaces (**Supplementary Table 7**), especially for EUR or those born in Europe. However, non-EUR groups have smaller sample sizes and are more dispersed, reflective of diversity in birthplaces for example due to forced migration. For example, over 90% of UKB participants assigned to AFR that have birthplaces outside of the UK are from Africa, with most of the remaining participants from North and South America. Genetic ancestry groups often correspond with the country of birth (**Supplementary Dataset 1**) or in some cases with known mass migration events. For example, the high rates of CSA who immigrated from Uganda likely reflects the expulsion of minorities of Asian descent from Uganda in the 1970's, many of whom were UK citizens and emigrated to the UK[12].

**Supplementary Table 6 | Comparison between inferred genetic ancestry (columns) and self-reported ethnicity (rows)**. UKB code 21000 provided self-reported ethnic background. Codings and meanings are defined by the UKB.

| Coding | Meaning | AFR | AMR | CSA | EAS | EUR | MID | OCE | Other |
|---|---|---|---|---|---|---|---|---|---|
| -3 | Prefer not to answer | 167 | 17 | 136 | 36 | 1,194 | 32 | 0 | 1 |
| -1 | Do not know | 13 | 16 | 36 | 9 | 107 | 22 | 0 | 1 |
| 1 | White | 1 | 4 | 1 | 1 | 534 | 2 | 0 | 3 |
| 1001 | British | 25 | 251 | 201 | 62 | 428,254 | 152 | 0 | 2,157 |
| 1002 | Irish | 0 | 1 | 4 | 0 | 12,694 | 1 | 0 | 59 |
| 1003 | Any other white background | 1 | 284 | 69 | 13 | 15,057 | 323 | 0 | 75 |
| 2 | Mixed | 11 | 8 | 7 | 3 | 11 | 6 | 0 | 0 |
| 2001 | White and Black Caribbean | 449 | 43 | 18 | 2 | 22 | 59 | 0 | 4 |
| 2002 | White and Black African | 277 | 15 | 15 | 2 | 21 | 71 | 0 | 1 |
| 2003 | White and Asian | 1 | 4 | 498 | 105 | 182 | 7 | 0 | 5 |
| 2004 | Any other mixed background | 122 | 124 | 173 | 133 | 361 | 79 | 0 | 4 |
| 3 | Asian or Asian British | 0 | 0 | 35 | 3 | 4 | 0 | 0 | 0 |
| 3001 | Indian | 0 | 0 | 5,684 | 0 | 13 | 0 | 0 | 19 |
| 3002 | Pakistani | 0 | 0 | 1,736 | 0 | 3 | 0 | 0 | 9 |
| 3003 | Bangladeshi | 0 | 0 | 221 | 0 | 0 | 0 | 0 | 0 |
| 3004 | Any other Asian background | 1 | 0 | 1,213 | 359 | 106 | 56 | 1 | 11 |
| 4 | Black or Black British | 24 | 0 | 0 | 0 | 2 | 0 | 0 | 0 |
| 4001 | Caribbean | 4,175 | 12 | 87 | 3 | 0 | 4 | 0 | 18 |

| | | | | | | | | | |
|---|---|---|---|---|---|---|---|---|---|
| 4002 | African | 2,989 | 0 | 12 | 2 | 1 | 187 | 0 | 14 |
| 4003 | Any other Black background | 104 | 2 | 8 | 0 | 1 | 2 | 0 | 1 |
| 5 | Chinese | 0 | 0 | 2 | 1,487 | 4 | 1 | 0 | 10 |
| 6 | Other ethnic group | 812 | 357 | 842 | 657 | 1,023 | 644 | 1 | 20 |

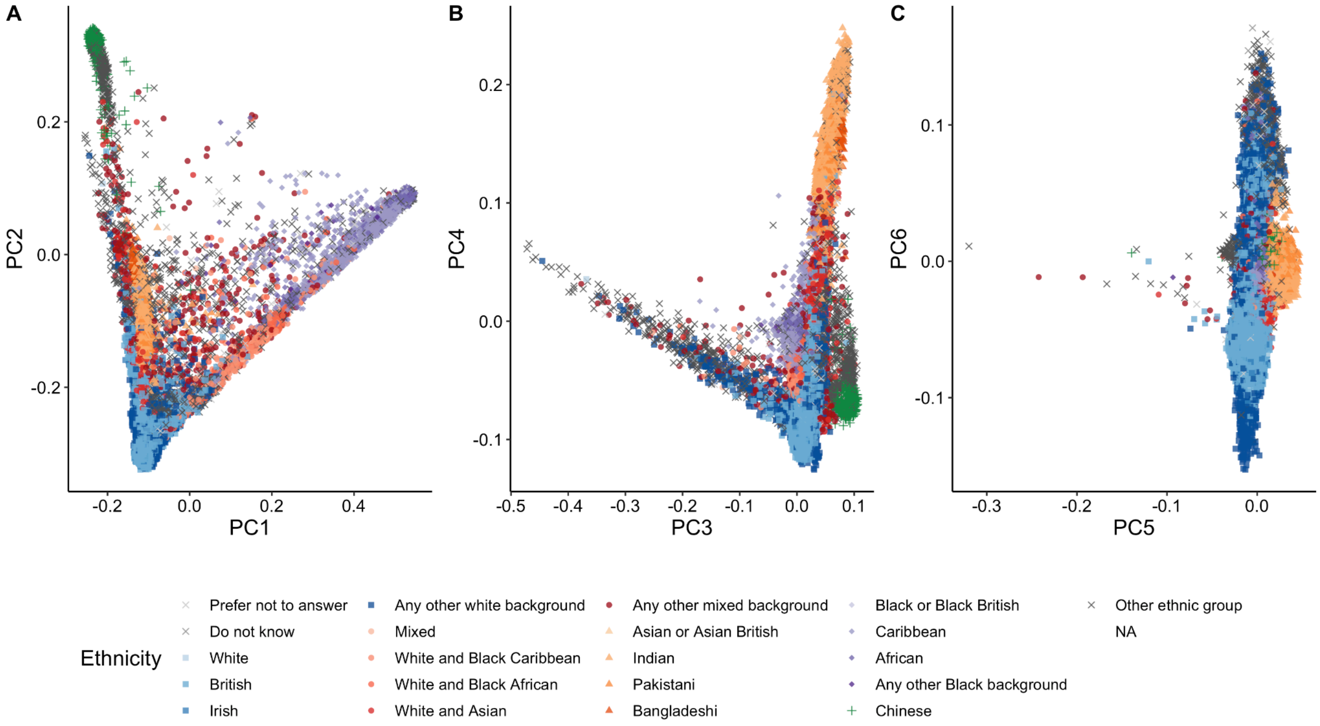


**Supplementary Figure 14 | Principal components roughly correlate with self-reported ethnicity.** Principal components are as shown in **Supplementary Fig. 1**.

**Supplementary Table 7 | Assigned population labels correlate with continental birthplaces.** Columns show assigned population labels. Rows indicate continental birthplaces. 3-letter continental ancestry codes are as in **Supplementary Table 1**. Cells shaded in green indicate the maximum fraction per row and column, which was used to calculate marginal percentages. Most shaded cells indicate the maximum for both rows and columns, but blue cells indicate which values were used to calculate only row marginals and yellow cells indicate which value was used to calculate only a column marginal when multiple cells in the row or column were shaded, respectively.

| | AFR | AMR | CSA | EAS | EUR | MID | OCE | oth | Percent aligned with majority |
|---|---|---|---|---|---|---|---|---|---|
| Africa | 5646 | 39 | 2626 | 100 | 3105 | 679 | 0 | 51 | 46.10% |
| Asia | 17 | 3 | 6414 | 2339 | 2246 | 341 | 1 | 60 | 56.16% |
| Europe | 42 | 25 | 37 | 24 | 9493 | 178 | 0 | 42 | 96.46% |
| North_America | 328 | 118 | 7 | 16 | 2131 | 7 | 0 | 14 | 81.30% |
| Oceania | 1 | 3 | 29 | 17 | 1595 | 5 | 1 | 12 | 95.91% |
| South_America | 186 | 623 | 102 | 17 | 378 | 16 | 0 | 7 | 46.88% |
| Percent aligned with majority | 90.77% | 76.82% | 69.60% | 93.08% | 50.10% | 55.38% | 50.00% | 27.42% | |

### Pre-GWAS quality control

#### Sample QC

We performed an initial sample QC using a sample exclusion criteria on UKB genotype data similar to previous approaches, including Bycrof et al[6] and a previous multi-phenotype GWAS from the Neale lab (referred to as “Round 2”). Specifically, we excluded individuals previously identified to have high autosomal genotype missingness (> 2%), outlier heterozygosity and missingness rates after adjusting for population structure, discordance between self-reported and genetically-inferred sex, putative sex chromosome aneuploidies, or excessive genetic relatedness (>200 estimated 3rd degree or closer relatives).

#### Variant QC

To define a set of variants for association analysis, we used imputed variants from UKB (97,059,328 variants from imputation version 3). We retained only variants with information scores > 0.8, resulting in 29,865,259 variants on the autosomes and X chromosome. For each population, we only considered variants with an allele count at least 20, as defined by the sum of the dosages; this resulted in a variable number of variants per population (**Supplementary Table 8**), spanning a union set of 28,987,534 variants. We performed further variant QC after associating genotypes to phenotypes (see QC of summary statistics, below).

**Supplementary Table 8 | Number of variants per population.**

| Population | Number of variants |
|---|---:|
| AFR | 21,964,524 |
| AMR | 11,624,137 |
| CSA | 15,969,799 |
| EAS | 10,238,883 |
| EUR | 23,861,814 |
| MID | 13,974,659 |
| Total | 28,987,534 |

#### Phenotype QC

##### Continuous and categorical traits

Phenotypes were processed using a custom version of PHESANT (https://github.com/astheeggeggs/PHESANT), a phenotype curation pipeline, as previously described[13,14] and summarized in **Supplementary Fig. 15**. We manually curated a collection of phenotypes for analysis, which were then processed through this pipeline which re-codes phenotypes and applies inherent orderings of ordinal categorical variables as specified in the data-coding file https://github.com/astheeggeggs/PHESANT/blob/master/variable-info/data-coding-ordinal-info-nov2019-update.txt. We ran the modified version of PHESANT using a 200Gb RAM virtual machine on the Google Cloud Platform. Phenotypes were curated jointly to ensure any recoding in the QC pipeline was applied consistently across ancestry groups. For continuous phenotypes with repeated measures, we average across measures applied to each sample. For both-sex and sex-specific analyses, inverse rank normalize transformation (IRNT) was applied across ancestry groups. Any sex-specific phenotypes present in the both-sex phenotype dataset were removed. A comparison of heritability estimates for some biomarker phenotypes from raw versus inverse-rank normal transformed values yielded higher heritability estimates for IRNT, so we opted to perform IRNT for all quantitative traits.

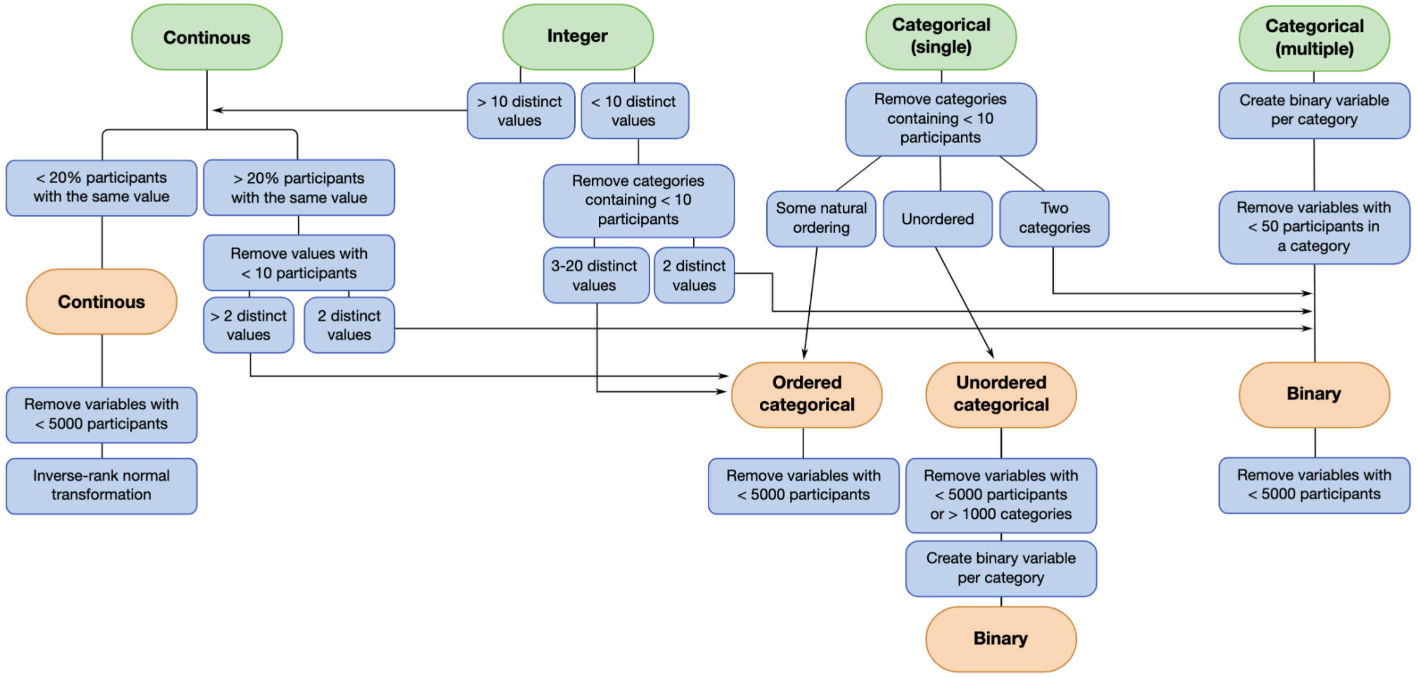


**Supplementary Figure 15 | Phenotype curation using a custom version of PHESANT.** The flowchart summarizes filtering and transformation steps to parse the unprocessed phenotype data. Figure adapted from Millard et al[15].

#### ICD-10 codes

ICD-10 codes were parsed using a custom Hail process. Briefly, UKB Fields 41202 (primary), 41204 (secondary), 41201 (external), and 40001 (cause of death) were expanded and combined such that if an individual had a given code in any of these four fields, the individual was marked as having that phenotype. Four digit codes (e.g. K509) were truncated to three digits (e.g. K50) and grouped together for analysis. ICD-9 codes were parsed in a similar fashion for use in phecode definitions, but were not used directly for genome-wide association analysis.

#### Phecodes

Phecodes[16] were processed using custom Hail and R scripts using the interim outputs from the createUKBphenome script (https://github.com/umich-cphds/createUKBphenome). Briefly, the aforementioned ICD-10 and ICD-9 codes were mapped to hierarchical disease endpoints as defined in the Phecode v1.2b1 (https://phewascatalog.org/phecodes). Per the Phecode definitions, cases were defined as those who have ICD-10 or ICD-9 codes (any of primary, secondary, external, or cause of death) listed in inclusion criteria, while controls were defined as those who were not included as cases for other Phecodes listed in exclusion criteria. If applicable, male-/female-specific analyses were conducted (*e.g.*, 174.1: Breast cancer [female]).

#### Prescriptions

We downloaded prescription data from UKB Field 42039 and processed through a custom pipeline to harmonize data. Specifically, of the ~82,000 unique prescriptions in the GP release, we selected only those 669 with > 10,000 instances to be used as phenotypes, as well as an additional 32 drugs used for Parkinson's (combination dopamine precursor/dopamine decarboxylase inhibitors). We built a manual curation pipeline wherein after splitting on the first white space and lower-casing the first token, converted each prescription string to 3 fields containing the generic name, dosage, and delivery system. We selected the first token as it often corresponds to the generic name and manually corrected mistakes. We created a simple ontology using a fourth field, giving for each prescription name, the corresponding pharmacological mechanism of action, drug category, and possible indication (for example, "Simvastatin TABS 40MG" would be represented as "simvastatin, 40mg, Tablet; HMG-CoA reductase inhibitor, statin"). We performed initial data exploration and extraction using Hail version 0.2.16-6da0d3571629 and Pandas version 0.24.2. The spreadsheet AirTable was used to facilitate the curation process, which was performed by an MD (G.S.).

We created the ontology with the aim of providing useful meta-data for grouping drugs/phenotypes, and it is intentionally redundant and not organized hierarchically. For example, ibuprofen will include both "NSAID" and "non-steroidal anti-inflammatory drug" and sertraline will include both "SSRI" and "selective serotonin reuptake inhibitor." Lactulose will include both "osmotic laxative" as well as "laxative."

To anticipate future needs, we investigated the potential to “bootstrap” from this small curated list to cover larger percentages of the full list of ~82,000 unique prescription strings in the GP release. First, we created an extensive list of regexes to automate the mapping from raw prescription string to the structured fields of possible generic, dosage, and delivery system. Next, to correct for mistakes in the possible generic (given that the first token is not always the correct generic name) as well as to fill out the crucial fourth field of pharmacological metadata, we performed a simple experiment. We randomly selected 20 prescriptions strings from a longer list of 3500 with >1000 instances, but which were not included in the original list of 669. By finding the best match (using the Levenstein distance or a related discrete metric) from our initial hand-curated list, we were able to correctly identify the generic name and drug category and indication 60% of the time, which could in principle simply be corrected, rather than manually curated from scratch, thereby significantly reducing the curation overhead. Our methodology suggests that by iteratively string matching and manually curating larger and larger regions of the long-tail of prescription strings, high-quality clinical phenotypes can be extracted without requiring full manual curation.

##### Other phenotypes

We built custom processes for some phenotypes, including those related to COVID-19 as well as custom phenotype combinations such as waist-hip ratio (phenotype computed from UKB code 48 / 49, then IRNT) and blood pressure traits, as well as two randomly generated phenotypes.

The final set of phenotypes can be found in **Supplementary Dataset 2**.

### Association analysis

All primary association analyses were performed using SAIGE[17], implemented in Hail Batch (https://hail.is), similar to the pipeline previously described[13]. Briefly, genetic data were extracted from Hail into BGEN files (one per megabase) and phenotypic data were extracted into TSV files (one per phenotype). For the creation of the GRM, we LD-pruned ($r^2 < 0.1$ for all genetic ancestry groups except EUR $r^2 < 0.05$) variants above 1% frequency, and in EUR, we removed a common inversion at 8:8055789-11980649 and the MHC region at 6:28477797-33448354 and downsampled the dataset by 45%. The null model (step 1) was built once for each phenotype with a GRM created using SAIGE. Score testing (step 2) was parallelized for each megabase using each BGEN file. Results were collected for each phenotype into Hail Table format, and across phenotypes into Hail MatrixTable.

For phenotype selection, we required at least 50 cases in each population except EUR where we required at least 100 cases given the larger sample size. In total, we assessed 1,105-7,185 phenotypes per population, each of which ranged from 980-420,531 individuals (**Fig. 1a**) and 10-23 million variants (**Supplementary Table 8**).

#### Computational framework

In order to rapidly generate the results, which required 3.8 million CPU-hours of computation time, we developed a scalable computational engine, Hail Batch. Batch is a Python module that allows the specification of tasks with dependencies in a directed acyclic graph. Importantly, the framework is scalable as it can be used within the internal Batch Service, a multi-tenant compute cluster in Google Cloud that is managed by the Hail team. At the time of use, this service enabled the simultaneous use of up to 100,000 CPUs, allowing this pipeline to be completed in approximately 6 days (wall-clock) at a cost of about $82K.

#### Covariates

We included the following covariates in each regression model: principal components (PCs) 1-10 (computed per-population, with PCs computed in unrelated individuals, then projecting related individuals, see Ancestry assignment and relatedness inference for more detail), age, sex, age * sex, $age^2$, and $age^2$ * sex.

#### Comparison of meta-analysis to mega-analysis

We compared two approaches to multi-ancestry association testing using mixed models for a pilot set of five phenotypes: a mega-analysis approach with all individuals in the same model, as well as a per-ancestry approach followed by meta-analysis. We compared inflation using two genomic control metrics, Lambda GC and Lambda 1000, and found consistent trends across all phenotypes, where the meta-analysis maintains the best control of stratification (lambda metrics closest to 1) with additional independent significant hits, indicating minimal evidence of inflation and better-controlled false-positive rates (**Extended Data Table 1**). Hence, we adopted this approach throughout the remainder of the project.

#### *Tractor* GWAS analysis

We performed an additional analysis to compare haplotype tract association methods (Tractor[9]) with the mixed models (SAIGE[17]) used throughout this manuscript. Tractor GWAS was conducted under a separate UKB application (95179). Thus, we mapped the Pan-UKB AFR sample list, consisting of 6,636 individuals, to the other application, resulting in 6,245 unrelated AFR individuals. For quality control prior to Tractor runs, we performed variant filtering using PLINK2[4], applying a 10% threshold for genotype and sample missingness, restricting to variants with MAF >= 0.5%, imputation score >= 0.8, and only biallelic SNPs. After this filtering, 9.95 million variants remained (compared to ~29 million in SAIGE). Unrelated AFR and EUR individuals were identified from the HGDP+1kGP joint-call dataset[18] for use in reference panels, which was lifted over to the GRCh37 reference genome using the GATK Picard LiftoverVcf tool[19]. Joint phasing of this dataset was performed using SHAPEIT5[20], followed by local ancestry inference using RFMix2[21]. ADMIXTURE[22] was also applied to assess global ancestry proportions. The global ancestry estimates from both RFMix2 and ADMIXTURE showed a strong correlation (r > 0.99), implying that local ancestry inference functioned well in this population. Using the local ancestry estimates from RFMix2, we ran Tractor GWAS for phenotype ID 30060 (Mean corpuscular Hb concentration), which was Inverse Rank-Normal Transformed (IRNT) using the RNOmni package in R[23]. The covariates used were identical to those used in the SAIGE analyses, including PC1-10, age, sex, age*sex, age2, and age2*sex.

### QC of summary statistics

#### Low frequency variants in cases

For binary traits, variants with very low allele counts in cases (e.g. a variant found only in one case) will produce unreliable summary statistics. To assess an appropriate cutoff, we computed the genomic control ($\lambda_{gc}$) for each phenotype across a range of minimum allele counts and frequencies (**Supplementary Fig. 16**). Notably, at case allele counts of less than 3, $\lambda_{gc}$ becomes unstable, and thus, we flagged as low confidence any association test where the allele count in cases or allele count in controls was less than 3.

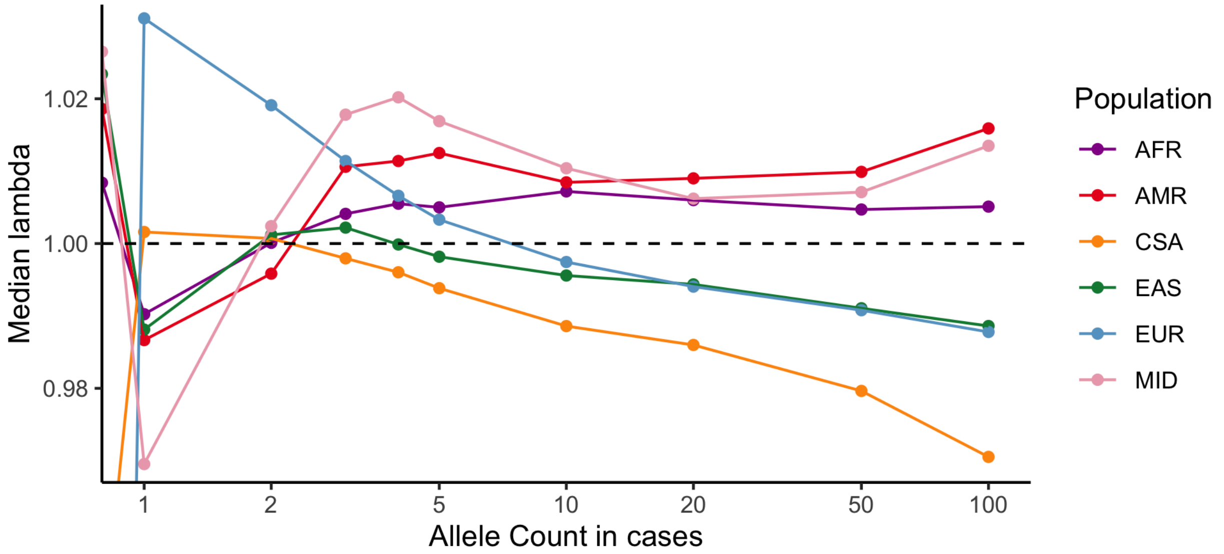


**Supplementary Figure 16 | Lambda by case allele count.** The median lambda across all binary traits by minimum allele count in cases is shown by population.

Further, for all traits (including quantitative), we also marked as low confidence those variants where the reference allele was found at fewer than 20 copies, indicating a rare variant in the reference genome.

#### Variants with discrepant frequency compared to gnomAD

In the course of assessing the robustness of the association statistics, we observed that a number of variants were found at significantly different frequencies from those found in gnomAD. For instance, 8:75857876 (rs11786917) is found at 30% frequency in the UKB EUR subset, while it is at 14.4% in the gnomAD NWE (North-Western European) subset. We thus investigated the discrepancy between UKB and gnomAD variation across populations, matching genetic ancestry labels where possible (AFR, AMR, EAS compared as-is, and EUR compared to NWE). We find the vast majority of variants have a concordant frequency (within a factor of two) between UKB and gnomAD (**Supplementary Fig. 17**a). However, a substantial number of variants are profoundly different in frequency, which is enriched for variants failing quality filters in gnomAD (**Supplementary Fig. 17**b). Among common (higher than 1% frequency in UKB) variants, variants that are discordant (greater than 2x difference in frequency) show a decreased transition/transversion (Ti/Tv) ratio (**Supplementary Fig. 18**), suggesting that they are enriched for errors.

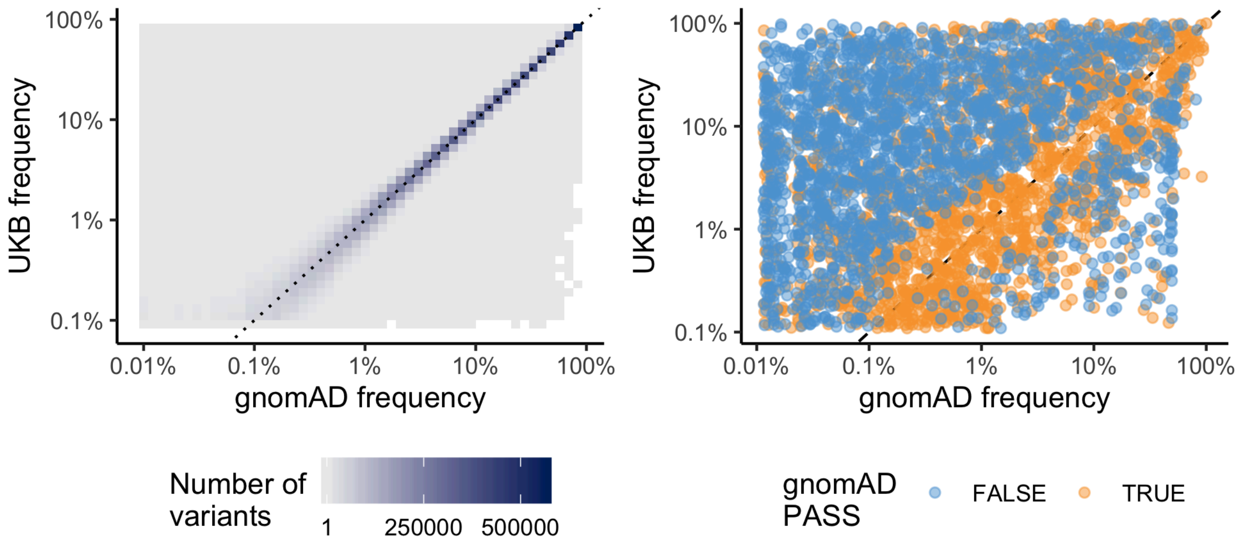


**Supplementary Figure 17 | UKB and gnomAD frequencies.** The frequencies in the AFR population in UKB and gnomAD are highly correlated (a), but many variants are discordant, especially at higher frequencies in UKB (b). These variants tend to fail quality filters in gnomAD. A similar pattern is observed for all populations overlapping between UKB and gnomAD (AFR, AMR, EAS, and EUR).

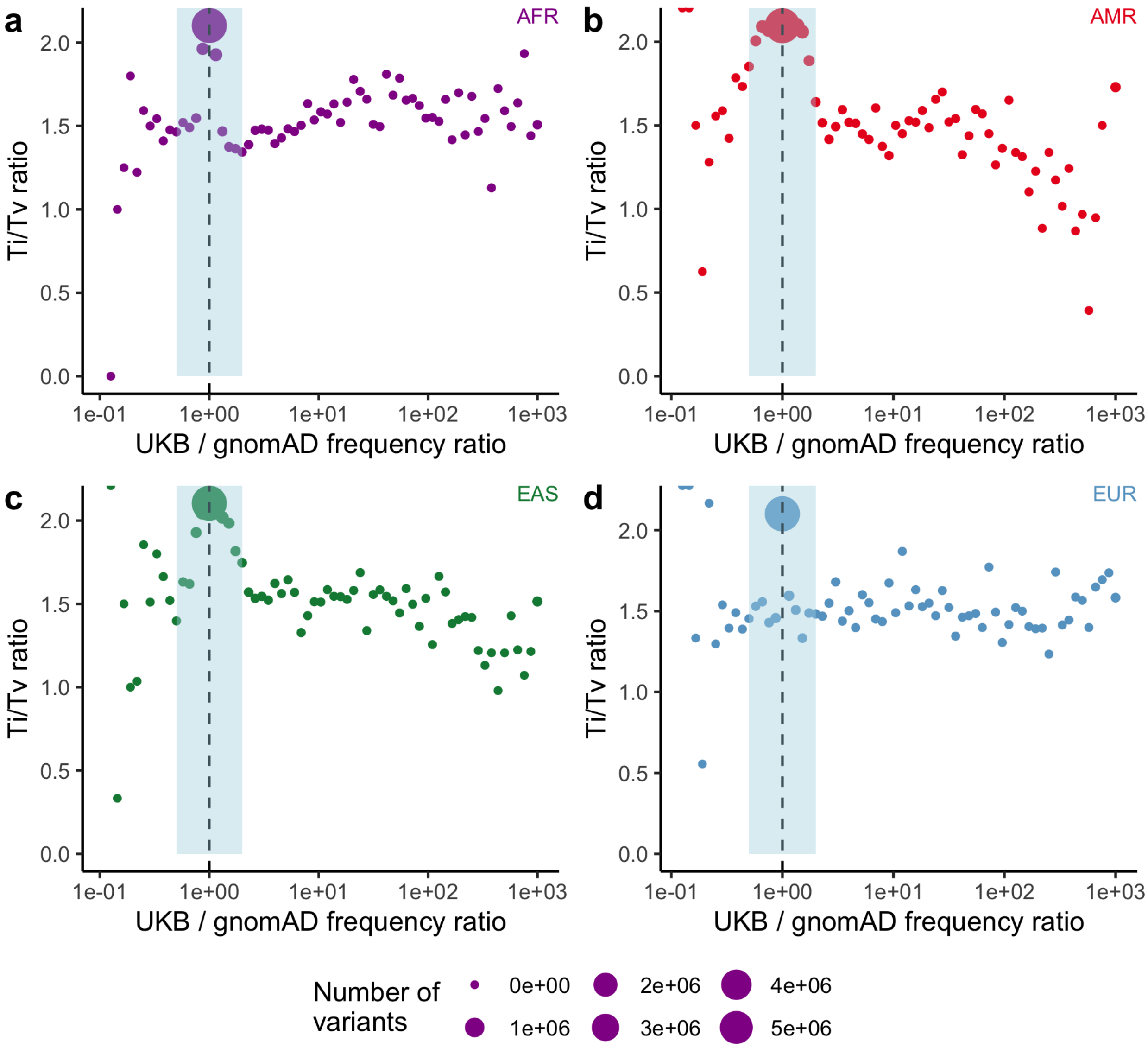


**Supplementary Figure 18 | Ti/Tv ratio of discrepant variants.** Variants that are discordant between UKB and gnomAD have lower Ti/Tv ratios. Points are colored by population (a: AFR, b: AMR, c: EAS, d: EUR) and sized proportional to the number of variants in the bin. Shaded region corresponds to variants that are "well-calibrated" (frequency within 2-fold of gnomAD) and thus retained for downstream analysis.

#### Variants missing from gnomAD

Additionally, there are 1,391,963 variants that are found in UKB that are not found in the gnomAD genomes. Similar to the variants that have discrepant frequencies, these variants have a reduced Ti/Tv ratio (1.69 compared to 2.1 for variants found in both UKB and gnomAD). We thus remove these variants along with 783,521 variants which have a discordant frequency in at least one population (**Supplementary Fig. 19**).

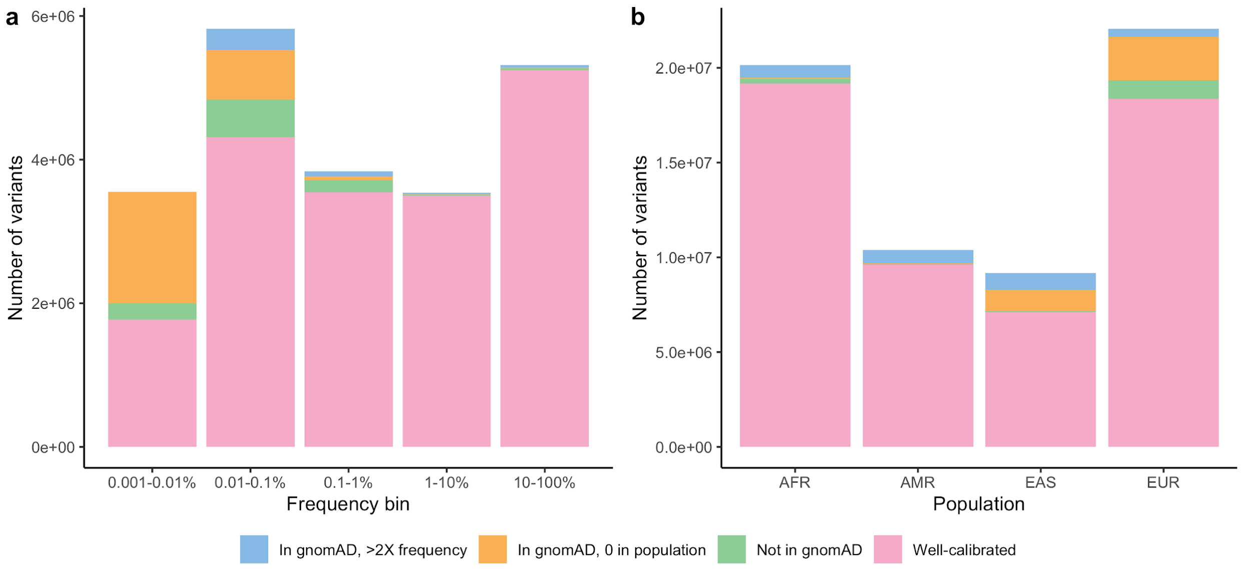


**Supplementary Figure 19 | Variants removed by gnomAD filters.** The number of variants that are well-calibrated (within 2X frequency) are compared to those missing from gnomAD, found in gnomAD but in a different population, or having a significantly different frequency from gnomAD. These metrics are broken down by UKB frequency within EUR (a) and by population (b).

### LD matrices and scores

LD matrices and scores were computed within each ancestry group. The genotype matrix (X) was standardized and variants were filtered to MAC > 20 (as for GWAS). For covariate correction, the residuals from the regression of $genotype \sim covariates$ were obtained via $X_{adj} = M_c X$ where $M_c = I - C(C^T C)^{-1} C^T$, the residual-maker matrix, and C is the matrix of covariates (as described above, Covariates). The LD matrix was produced via $\hat{r} = \frac{X^T_{adj} X_{adj}}{n}$ with a window size of 1 MB, with a bias adjustment by $\tilde{r}^2 = \frac{n-1}{n-2}\hat{r}^2 - \frac{1}{n-2}$. LD scores were subsequently computed summing the bias-adjusted values within a radius of 1 MB.

To better understand the similarities and differences between imputation-based and sequence-based LD scores and provide a more general comparison between LD scores from two separate cohorts, we next compared these LD scores with those obtained from gnomAD[24]. gnomAD within-ancestry LD matrices were previously computed after genotype standardization and variant filtering to MAF > 0.005 via $\hat{r} = \frac{X^T X}{n}$ with a 1 MB radius, with a bias adjustment and LD scores computed as above. We observed very strong concordance of LD scores between gnomAD and UKB (**Supplementary Fig. 20**). Computed correlation coefficients were above 0.9, with highest values observed for EUR and EAS, compared to AMR and AFR, potentially due to divergence between UK Biobank and gnomAD in these groups.

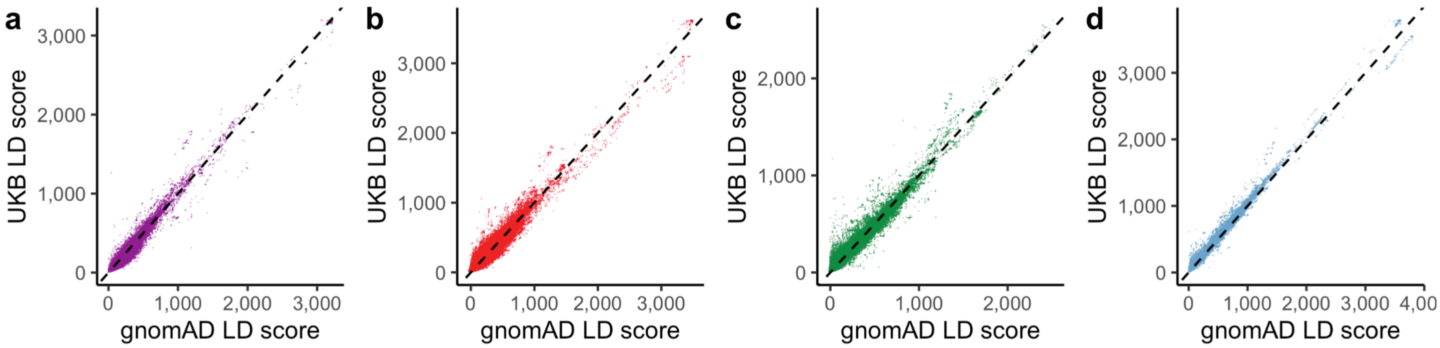


**Supplementary Figure 20 | Pairwise comparisons of LD scores in UKB vs. gnomAD within each genetic ancestry group**. Hapmap 3 SNPs are shown for (a) AFR, (b) AMR, (c) EAS, (d) EUR (compared to gnomAD NFE). Dashed line represents y=x.

For all meta-analysis results, we constructed reference panels of 5,000 individuals matched by genetic ancestry proportions to the given meta-analysis, and computed LD matrices and scores from these panels.

#### Heritability analysis

To better understand genetic architectures across a wide range of phenotypes and ancestries, we next estimated the SNP-heritability ($h^2_{SNP}$) for each GWAS. We thus first computed heritability using univariate LD score regression (LDSC) for GWAS summary statistics across 16,518 ancestry-trait pairs (7,228 unique phenotypes where at least one population had lambda GC between 0.5-2). Previous work has shown that the distribution of MAF and/or LD for causal variants differs from genome-wide variants, causing biased estimates of heritability[25,26]. In this manuscript, we compare our results to those of a previous multi-phenotype GWAS performed by the Neale lab (hereafter referred to as “Round 2”). Indeed, while LDSC heritability estimates from Europeans were highly correlated with Round 2 (**Supplementary Fig. 21**a), they appeared to be biased downwards as Round 2 estimates were computed using stratified LD score regression (S-LDSC) using the baselineLD v1.1 model. To improve our LDSC-based heritability estimates, we created 25 in-sample MAF and LD bins based on quintiles of each ancestry-specific feature, then used the resultant LD-scores to compute heritability while modeling MAF and LD[27,28]. We constructed LD bins using LD-scores computed as described above. Encouragingly, this S-LDSC approach largely resolved biases observed for ordinary LDSC (**Supplementary Fig. 21**b).

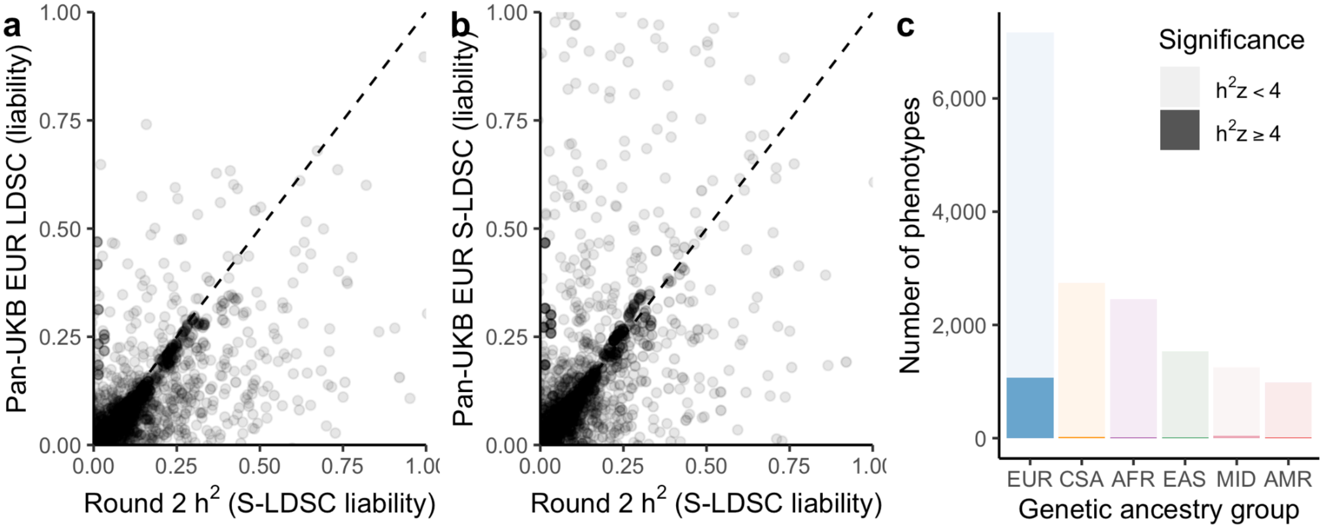


**Supplementary Figure 21** | (a-b) Correlation between UKB round 2 and Pan-UKB EUR LDSC (a) and S-LDSC (b). (c) The number of phenotypes by genetic ancestry group, shaded by significant heritability *z* scores (S-LDSC $h^2$ $z \geq 4$).

Heritability estimates in smaller ancestry groups (n < 10,000) were extremely noisy regardless of whether LDSC or S-LDSC methodology was used, with only 88 of 8,966 tested ancestry-trait pairs in non-EUR ancestries showing a heritability *z* score > 4, compared to 1,063 of 7,165 tested traits in EUR (**Supplementary Fig. 21**c). This is likely attributable to the low sample size of non-EUR genetic ancestry groups. Indeed, previous work has demonstrated that LDSC has lower power than genotype-based heritability estimation methods in low sample sizes[29].

To improve our heritability estimation power among non-EUR ancestry groups, we used multi-component Haseman-Elston regression implemented in RHE-mc[30]. This approach offers improved power by using a genotype-based analysis. We restricted to variants with MAF > 0.01 in Hardy-Weinberg equilibrium ($p > 10^{-7}$) in all populations, filtered out the MHC region (chr6:25Mb-35Mb), and removed related individuals (see Pruning ancestry outliers). This resulted in 4,923,127 SNPs across 376,430 individuals in all 6 genetic ancestry groups. To account for previously described relationships between heritability, LD-score, and MAF, we constructed MAF and LD bins: (1) 25 bins based on quintiles of LD score and MAF bins of size 0.1 as defined for S-LDSC, and (2) 8 bins based on quartiles of LD scores and two MAF bins (0-5%, 5-50%) as a sensitivity analysis. We included the same covariates as were used for GWAS, except for expanding to 20 PCs to match the original RHE-mc implementation[30]. Any phenotypes coded specifically for males or females were analyzed without inclusion of sex as a covariate. Because RHE-mc uses random vectors to improve computational efficiency at the expense of added run-to-run variability, we quantified the degree of run-to-run variability observed at various numbers of random vectors (**Supplementary Fig. 22**) and chose 50 random vectors to balance computational cost and variance.

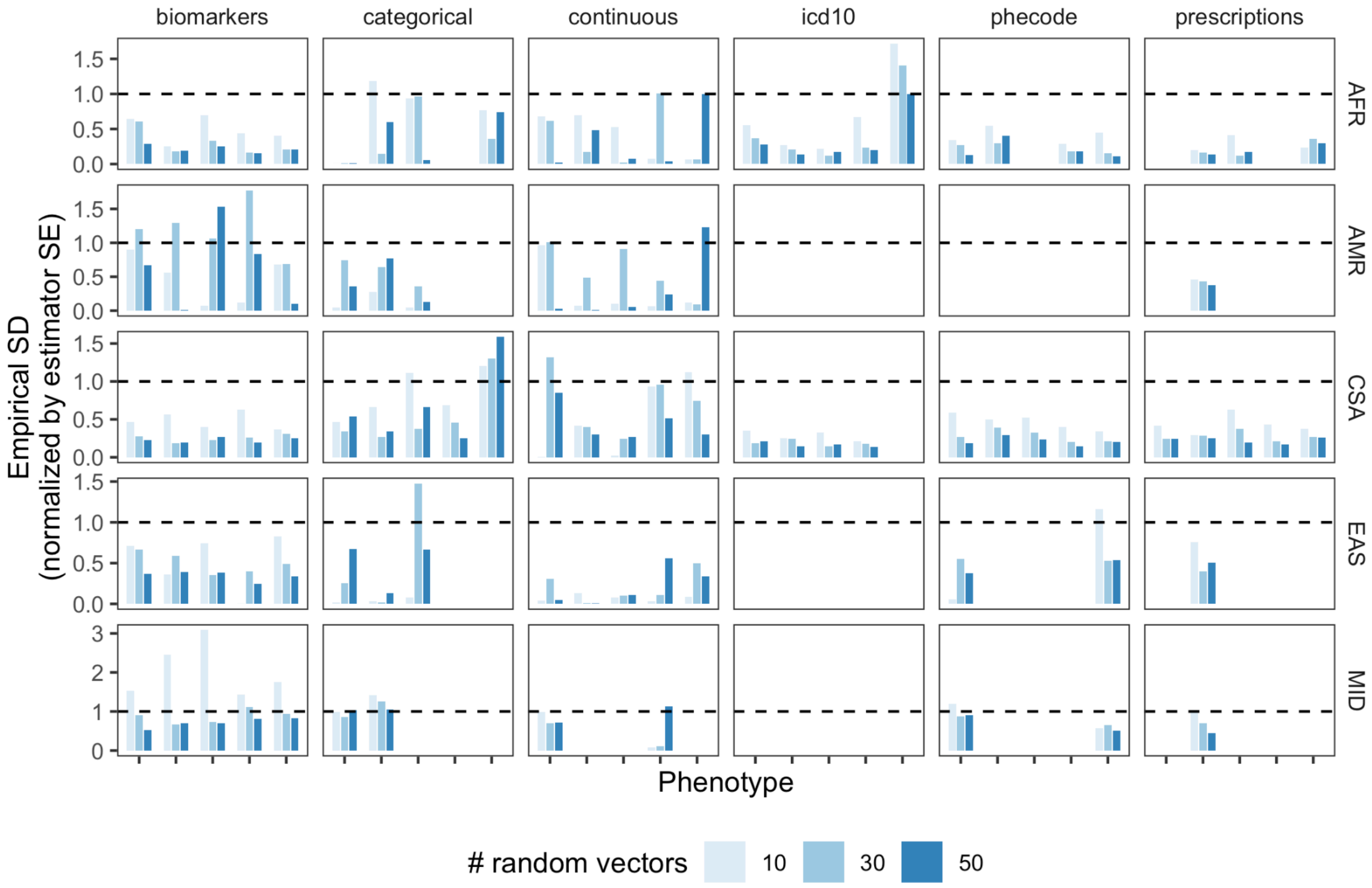


**Supplementary Figure 22 |** Characterization of RHEmc run-to-run variability. The first five phenotypes in the manifest are shown. Bars indicate empirical standard deviations (standard deviation of heritability estimates from 50 identical runs of heritability computation) normalized by the standard error of the heritability estimator for each phenotype. Missing points indicate failed convergence. Colors correspond to number of random vectors, indicating that variability goes down as the number of random vectors increases. We chose 50 random vectors for downstream analysis.

We took several approaches to benchmark this approach with respect to previously computed heritabilities and alternative methods. We computed heritability among the EUR group using RHE-mc for a selection of well-behaved, well-powered phenotypes including hypertension, type 2 diabetes, cholesterol, and others (**Supplementary Table 9**). These results showed high concordance with S-LDSC results computed using summary statistics from GWAS of the same phenotypes (**Fig. 2a**) as well as with heritability estimates from previous EUR-only genetic analyses in UKB (**Supplementary Fig. 23**). Given this close agreement and to avoid the untenable computational cost of using RHE-mc for EUR across UKB, we report only LDSC/S-LDSC heritabilities for all phenotypes in EUR. As we observed little difference between point estimates computed using 25 bins and 8 LD and MAF bins, we decided to proceed with estimates based on 25 bins for non-EUR ancestry groups to better model causal variant heterogeneity across the LD and MAF distribution, while using 8 bins for the pilot set of phenotypes tested in EUR to decrease computational complexity.

**Supplementary Table 9** | 66 pilot phenotypes chosen for heritability analysis using multiple methods. In the phenotype manifest, phenocode 20002's description is "Non-cancer illness code, self-reported" and here, the coding description is shown instead. The "note" column refers to the phenotype coding from UK Biobank, except in the cases of "irnt" which denotes that the phenotype was inverse rank normal transformed (typically noted in the "modifier" column of the manifest and release files).

| Trait type | Pheno code | Note | Description | Trait type | Pheno code | Note | Description |
|---|---|---|---|---|---|---|---|
| biomarkers | 30600 | irnt | Albumin | categorical | 20002 | 1113 | emphysema/chronic bronchitis |
| biomarkers | 30610 | irnt | Alkaline phosphatase | categorical | 20002 | 1202 | urinary frequency / incontinence |
| biomarkers | 30620 | irnt | Alanine aminotransferase | categorical | 20002 | 1464 | rheumatoid arthritis |
| biomarkers | 30630 | irnt | Apolipoprotein A | categorical | 20002 | 1465 | osteoarthritis |
| biomarkers | 30640 | irnt | Apolipoprotein B | categorical | 20002 | 1473 | high cholesterol |
| biomarkers | 30650 | irnt | Aspartate aminotransferase | categorical | 20002 | 1478 | cervical spondylosis |
| biomarkers | 30660 | irnt | Direct bilirubin | categorical | 20002 | 1657 | septicaemia / sepsis |
| biomarkers | 30670 | irnt | Urea | categorical | 3393 | 3393 | Hearing aid user |
| biomarkers | 30680 | irnt | Calcium | categorical | 6148 | 5 | Eye problems/disorders (Macular degeneration) |
| biomarkers | 30690 | irnt | Cholesterol | categorical | 6150 | 3 | Vascular/heart problems diagnosed by doctor (Stroke) |
| biomarkers | 30700 | irnt | Creatinine | continuous | 21001 | irnt | Body mass index (BMI) |
| biomarkers | 30710 | irnt | C-reactive protein | continuous | 3148 | irnt | Heel bone mineral density (BMD) |
| biomarkers | 30720 | irnt | Cystatin C | continuous | 4079 | irnt | Diastolic blood pressure, automated reading |
| biomarkers | 30730 | irnt | Gamma glutamyltransferase | continuous | 4080 | irnt | Systolic blood pressure, automated reading |
| biomarkers | 30740 | irnt | Glucose | continuous | 50 | irnt | Standing height |
| biomarkers | 30750 | irnt | Glycated haemoglobin (HbA1c) | continuous | 51 | irnt | Seated height |
| biomarkers | 30760 | irnt | HDL cholesterol | continuous | eGFR | irnt | Estimated glomerular filtration rate, serum creatinine |
| biomarkers | 30770 | irnt | IGF-1 | icd10 | H26 | | H26 Other cataract |
| biomarkers | 30780 | irnt | LDL direct | icd10 | I48 | | I48 Atrial fibrillation and flutter |
| biomarkers | 30790 | irnt | Lipoprotein A | icd10 | K21 | | K21 Gastro-oesophageal reflux disease |
| biomarkers | 30800 | irnt | Oestradiol | icd10 | K29 | | K29 Gastritis and duodenitis |
| biomarkers | 30810 | irnt | Phosphate | icd10 | K44 | | K44 Diaphragmatic hernia |
| biomarkers | 30820 | irnt | Rheumatoid factor | icd10 | K57 | | K57 Diverticular disease of intestine |
| biomarkers | 30830 | irnt | SHBG | icd10 | N18 | | N18 Chronic renal failure |
| biomarkers | 30840 | irnt | Total bilirubin | phecode | 153 | | Colorectal cancer |

| | | | | | | |
|---|---|---|---|---|---|---|
| biomarkers | 30850 | irnt | Testosterone | phecode | 172 | Skin cancer |
| biomarkers | 30860 | irnt | Total protein | phecode | 250.1 | Type 1 diabetes |
| biomarkers | 30870 | irnt | Triglycerides | phecode | 250.2 | Type 2 diabetes |
| biomarkers | 30880 | irnt | Urate | phecode | 290.1 | Dementias |
| biomarkers | 30890 | irnt | Vitamin D | phecode | 332 | Parkinson's disease |
| categorical | 20002 | 1065 | hypertension | phecode | 411 | Ischemic Heart Disease |
| categorical | 20002 | 1074 | angina | phecode | 411.2 | Myocardial infarction |
| categorical | 20002 | 1076 | heart failure/pulmonary odema | phecode | 530.12 | Ulcer of esophagus |

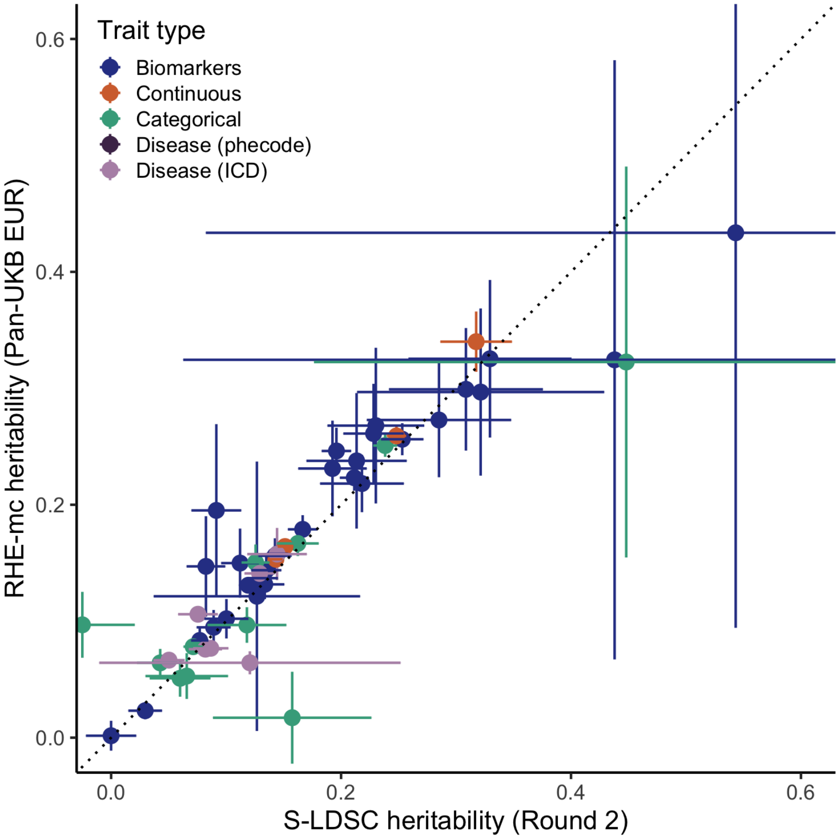


**Supplementary Figure 23** | Correlation between RHE-mc heritability point estimates (liability scale) and point estimates made in a previous round of heritability analysis restricted to the White British subset of UKB (Round 2) for the same pilot phenotypes. Color represents trait type, dotted line is y=x, error bars are +/- 1se.

For the smaller subsamples, HE regression produced substantially less noisy estimates than S-LDSC (**Figure 2b**), suggesting improved power. A cross-method comparison of heritability point estimates for selected continuous phenotypes revealed typically similar or higher point estimates observed using HE regression over LDSC and S-LDSC in non-EUR ancestries (**Supplementary Fig. 24**), as expected given the previously documented downward bias with LDSC in lower-powered settings[29]. We thus created a "final" heritability ($h^2_{SNP}$, hereafter referred to as $h^2$) estimate using S-LDSC for EUR, and RHE-mc for the remaining genetic ancestry groups. As expected given the relative sample sizes for each ancestry group, we observed the most traits with heritability *z* score ≥ 4 among EUR and CSA, with the fewest observed in AMR (**Supplementary Fig. 25**a). Of phenotypes meeting a cutoff of $h^2$ *z* ≥ 4 in at least one ancestry group, we observed that 1013 (62%) of phenotypes were significant in only one genetic ancestry group, 439 (27%) were significant in two genetic ancestry groups, and 184 (11%) were significant in three or more (**Supplementary Fig. 25**b).

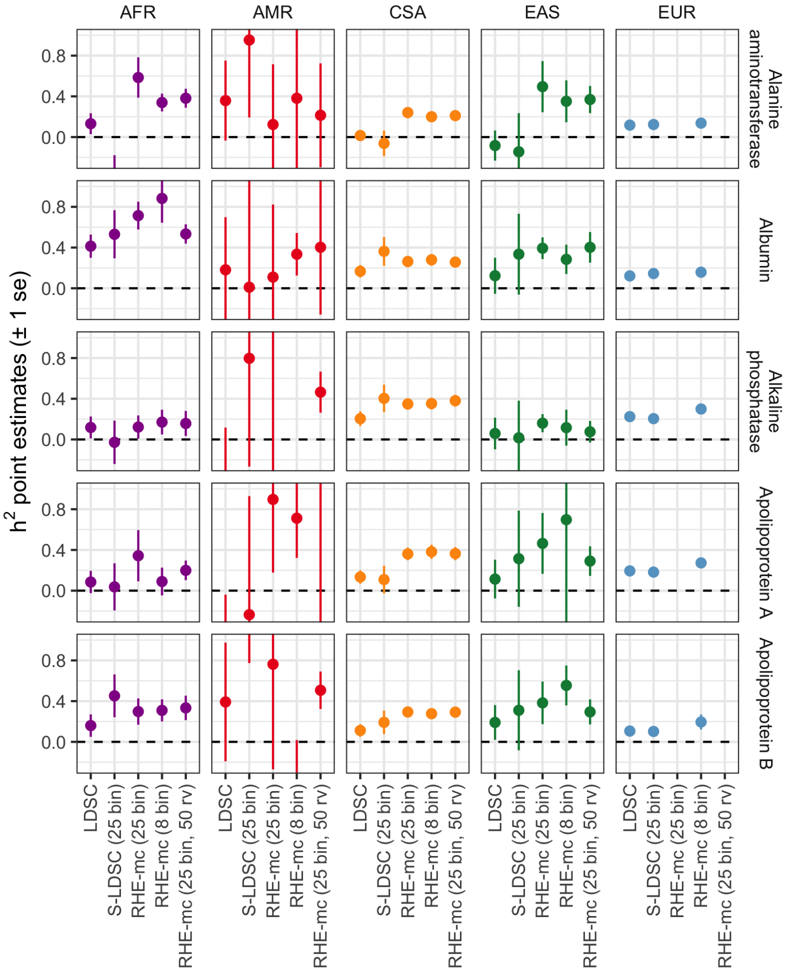


**Supplementary Figure 24** | Cross-method comparison of selected continuous phenotypes. Error bars represent +/- 1se. Only ancestry-trait pairs passing QC were included in this figure. RHEmc 25 bin (and 25 bin, 50 random vectors [RV]) was not run for EUR due to computational limitations.

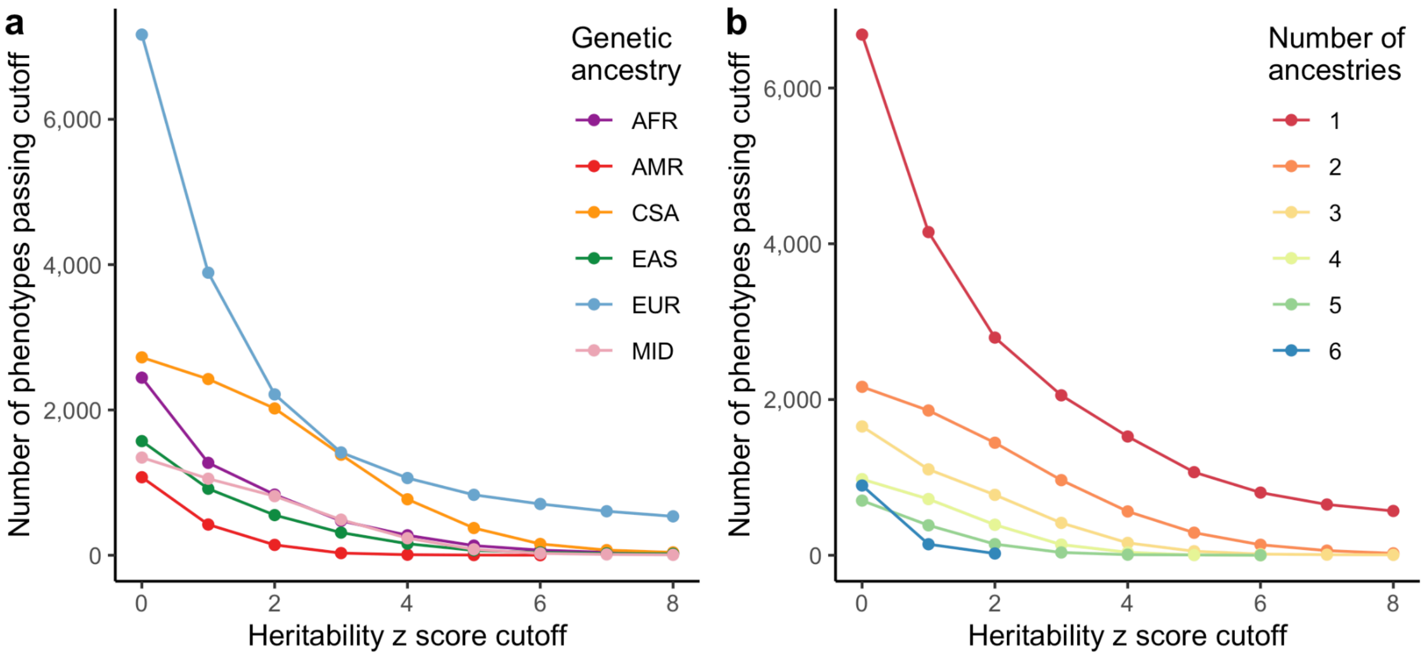


**Supplementary Figure 25** | Overview of heritability *z* scores across trait types and populations. (a) The number of traits passing in each ancestry as a function of $h^2$ *z* score cutoff (S-LDSC for EUR, RHEmc [25 bins] for all other ancestries). (b) The number of traits passing in 1, 2, 3, 4, 5, or all 6 ancestries (colors) as a function of the *z* score cutoff. The ancestry-trait pairs used in this plot are pre sumstats QC. S-LDSC -derived *z* scores reported for EUR, RHEmc (25 bins) reported for all other phenotype-ancestry pairs.

Analogous to the canonical example of an HLA allele more frequent in individuals with EAS ancestry naively showing association with chopstick usage[1], several traits in the UKB have high potential to produce spurious genetic associations due to confounding – e.g., food intake traits, geographic coordinates, country of birth. Indeed, we have observed that some of these traits display highly abnormal GWAS signals (**Supplementary Fig. 26**).

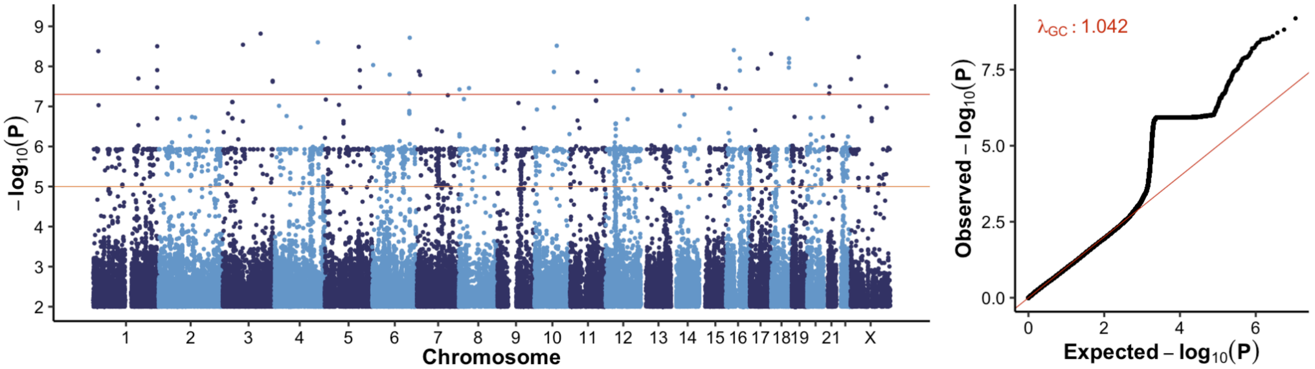


**Supplementary Figure 26** | Example of a QC-fail GWAS of categorical phenotype 3446 in the AMR genetic ancestry group, “type of tobacco currently smoked”, for category “Manufactured cigarettes” shown as a Manhattan plot (left) and a QQ plot (right).

Further complicating the interpretation of heritability differences across ancestry groups, we observed large systematic shifts in heritability point estimates across traits. Specifically, we observed higher heritability estimates overall in the CSA compared to other genetic ancestry groups, potentially reflecting a higher degree of heterogeneity (**Supplementary Fig. 4**) and residual stratification in the CSA GWAS results. Deeper exploratory analysis revealed that some phenotypes prone to cultural stratification (e.g., dietary preferences) tended to produce significantly out-of-bounds heritability estimates, abnormal $\lambda_{GC}$ estimates, and/or elevated S-LDSC ratio statistics.

To systematically identify traits with potentially problematic GWAS results, we devised a sequential filtering strategy leveraging heritability statistics (**Supplementary Fig. 27**a):

1. We identified phenotypes with sufficient power for downstream heritability-based analyses, by restricting to ancestry-trait pairs showing heritability significantly greater than 0 ($h^2$ $z \geq 4$).
2. As only 8 phenotypes passed QC in the AMR subsample (n = 975), we removed AMR ancestry-trait pairs from downstream analyses.
3. Interestingly, traits with significant, out of bounds observed-scale heritability point estimates ($h^2 \leq 0$, or $h^2 \geq 1$ with $h^2$ $z \geq 4$) appeared to be highly enriched for those especially prone to potential confounding based on their phenotype definitions (e.g. country of birth, ethnicity, occupation). We thus eliminated any traits with ≥ 1 genetic ancestry group showing significant out-of-bounds heritability estimates.
4. To avoid inclusion of phenotypes with substantial test statistic deflation, we also remove any traits with ≥ 1 genetic ancestry group with $\lambda_{gc} \leq 0.9$ (**Supplementary Fig. 27**b).
5. We then leveraged the ratio of the LDSC intercept-1 to the mean □$^{2}$ statistic-1 as a measure of the component of trait polygenicity (captured by mean chi-squared) explainable by population stratification (captured by the LDSC intercept[27]), and further computed a $z$ score for this ratio by dividing by the standard error. Based on the observed distribution of the LDSC ratio (**Supplementary Fig. 27**c), we eliminated traits with a high ratio (> 0.3) and a high ratio $z$ score (≥4). Although LDSC-based heritability estimation was not well powered for non-EUR ancestry groups, outliers from LDSC-based ratio

statistics for these cohorts nonetheless empirically identified several traits that appeared stratified (e.g., location-based and food intake traits).

6. To focus on traits that are well-powered in the UKB across diverse ancestries, we created a flag to indicate phenotypes that passed QC in the EUR ancestry group as well as at least one other ancestry group.

a

| Phenotype QC filters | Total | EUR | CSA | AFR | EAS | MID | AMR |
|---|---|---|---|---|---|---|---|
| GWAS run | 16,518 | 7,165 | 2,771 | 2,493 | 1,612 | 1,372 | 1,105 |
| Defined heritability estimate | 16,269 | 7,115 | 2,724 | 2,444 | 1,571 | 1,341 | 1,074 |
| Heritability z >= 4 | 2,485 | 1,037 | 771 | 275 | 159 | 235 | 8 |
| Remove AMR due to limited power | 2,477 | 1,037 | 771 | 275 | 159 | 235 | 0 |
| Heritability within bounds for all ancestries | 2,210 | 993 | 745 | 193 | 84 | 195 | 0 |
| Lambda GC > 0.9 for all ancestries | 2,186 | 988 | 742 | 187 | 81 | 188 | 0 |
| S-LDSC ratio < 0.3 or ratio z < 4 in all of EUR, CSA, or AFR | 2,049 | 926 | 699 | 173 | 77 | 174 | 0 |
| Passes all filters in EUR and at least 1 other ancestry group | 1,091 | 452 | 349 | 92 | 62 | 136 | 0 |

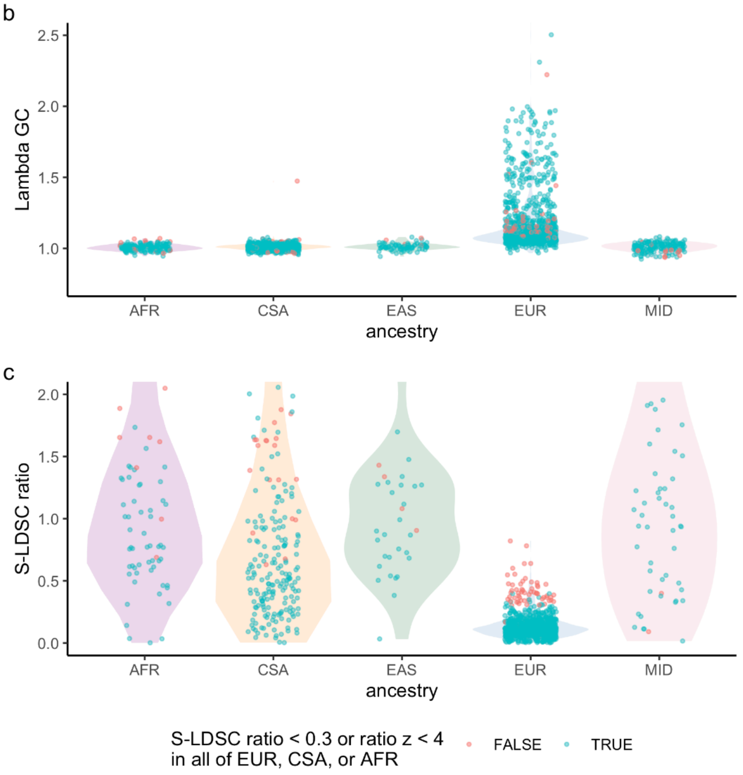


**Supplementary Figure 27** | Empirical summary statistics quality control approach. (a) Flowchart of QC approach with each filter used (left) as well as the number of phenotype-ancestry pairs passing each filter. Note that filters are applied sequentially in the listed order. The "heritability within bounds for all ancestries" and "lambda GC > 0.9 for all ancestries" fail for all ancestries if a single ancestry fails the respective filter. "S-LDSC ratio < 0.3 or ratio *z* score < 4 in all of EUR, CSA, or AFR" fail for all ancestry if any of EUR, CSA, AFR fail, but fail for the individual ancestry-trait pair only if the filter fails for a different ancestry group. (b-c) The distribution of lambda GC (b) and S-LDSC ratio (c) values by genetic ancestry group. Phenotypes that fail the S-LDSC ratio (referred to as "Controlled S-LDSC ratio" in Figure 2c) are highlighted in red.

Overall, we pruned 15,643 ancestry-trait pairs with available GWAS to 1,091 ancestry-phenotype pairs (452 unique phenotypes) that passed all filters and were significantly heritable in at least one non-EUR ancestry group (**Supplementary Fig. 27**). Supporting our choice of methods and the Z >= 4 cutoff, we observed that traits that showed significant heritability across multiple AFR/AMR/CSA/EAS/MID ancestry groups were also more likely to show Z >= 4 in the EUR group (**Supplementary Fig. 28**). We provide all final phenotype metrics in **Supplementary Dataset 2** and all component heritability estimates in **Supplementary Dataset 3**.

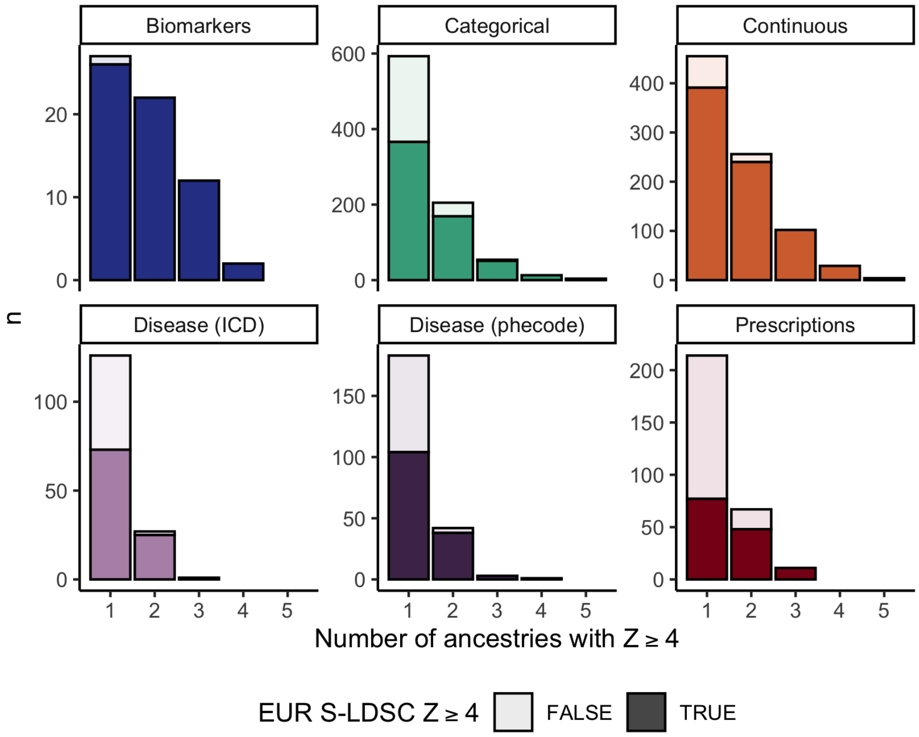


**Supplementary Figure 28** | Number of ancestry-trait pairs per trait type passing the *z* score >= 4 filter as a function of (1) EUR S-LDSC *z* >= 4, and (2) the total number of ancestry groups passing this filter, shown cumulatively. A greater proportion of the bar colored dark indicates a greater proportion of ancestry-trait pairs passing *z* >= 4 in a given number of ancestries also passed *z* >= 4 in EUR.

Of the phenotype-ancestry GWAS pairs that passed, 207 were shared between the two largest ancestry groups (EUR and CSA), with 110 phenotypes shared between three groups and 37 shared between four or more (**Supplementary Fig. 29**).

Finally, for convenience, we computed pairwise genetic correlations using LD score regression for 528 phenotypes for the EUR genetic ancestry group, which we make available as **Supplementary Dataset 4**.

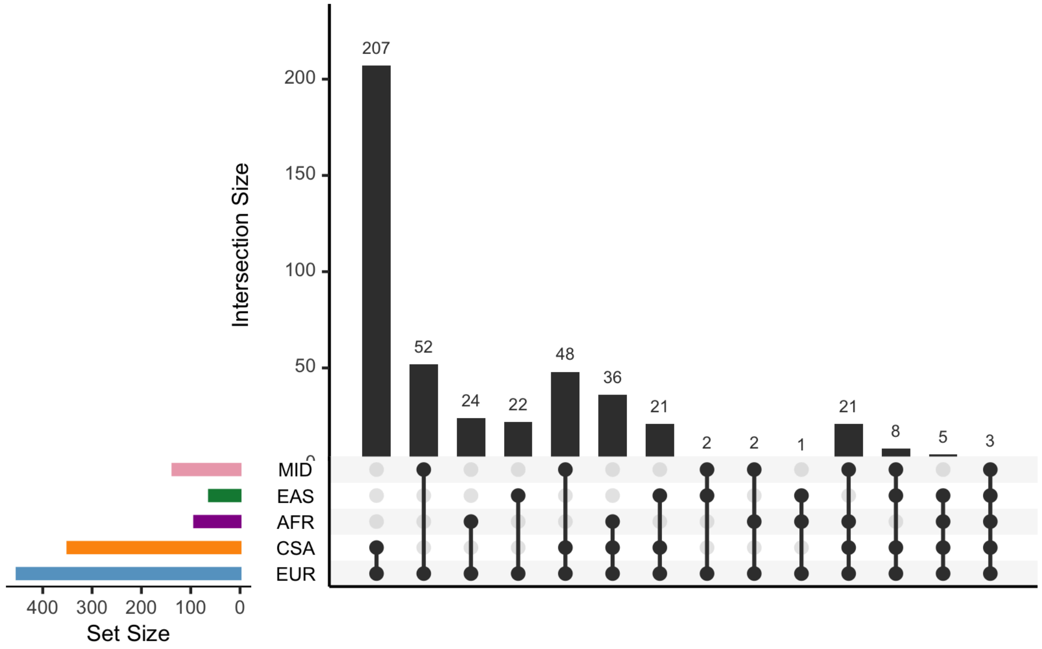


**Supplementary Figure 29** | Number of phenotypes passing final quality control steps by combination of genetic ancestry groups in which the phenotype passes.

### Maximal independent set

To ensure that closely related phenotypes were not double-counted (e.g. left and right arm fat percentage), we generated a maximal independent set of phenotypes for aggregate phenotypic comparisons to reduce bias among trait representation for broad analyses. To do so, we first generated a pairwise phenotypic correlation matrix in Hail, after regressing out covariates (see Covariates, above) from each phenotype. Based on the distribution of pairwise correlations for 452 phenotypes that pass QC as described above (**Supplementary Fig. 30**a), we filtered to pairs of traits with correlation $r^2 \geq 0.1$ (**Supplementary Fig. 30**b). We then generated the maximal independent set of phenotypes with hl.maximal_independent_set() two ways using a tiebreaker of higher case count (higher sample size for continuous phenotypes), retaining 151 phenotypes. We provide the full set of pairwise correlations at gs://ukb-diverse-pops-public/misc/pairwise/pairwise_correlations_regressed.txt.bgz and the filtered set in **Supplementary Dataset 5**.

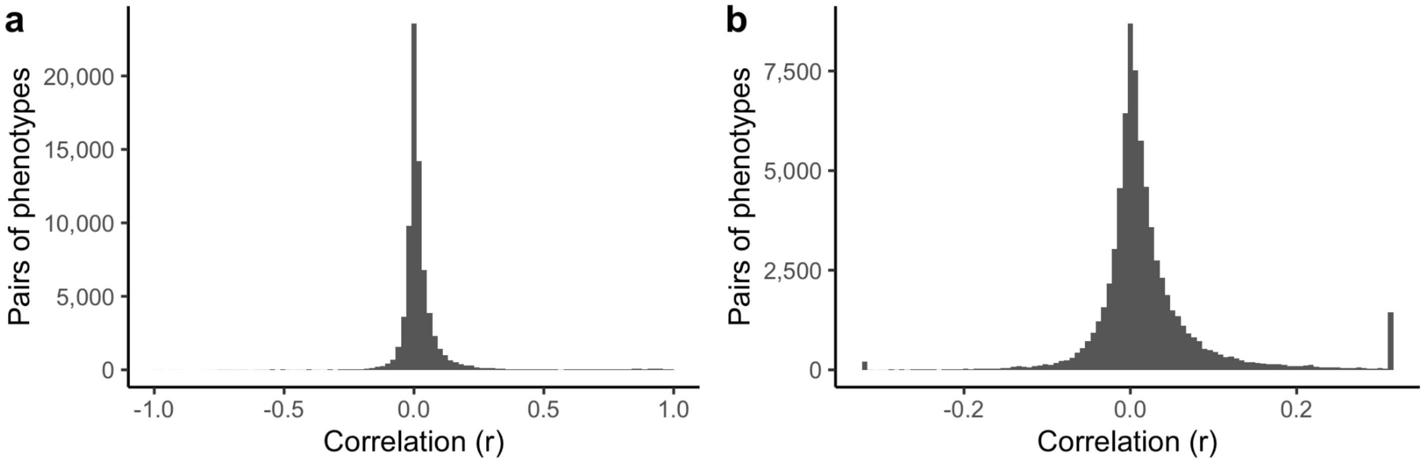


**Supplementary Figure 30** | Distribution of pairwise phenotype correlations across all individuals for filtered high-quality phenotypes, for all correlations (a), and zoomed to a correlation threshold of $r^2 = 0.1$ (b), which was selected to prune for independent phenotypes.

#### Locus definition within and across populations

Single ancestry and multi-ancestry LD reference panels for Plink clumping were created by randomly sampling 5,000 individuals from all samples available for a given set of ancestry groups. This probabilistic approach does not guarantee that the proportion of ancestry groups represented within an LD reference panel exactly matches the proportion of ancestry groups among all possible individuals available to be sampled for that reference panel. However, in expectation, these proportions should match. In the case of ancestry groups having fewer than 5,000 total individuals (AMR, EAS, and MID ancestry groups), the corresponding single-ancestry LD reference panels for those ancestry groups were exactly the same as the full sample for each ancestry group, i.e. no subsampling was performed.

Clumping was run using Plink in Hail Batch. P-value thresholds for index variants and variants within a clump were both 1. The lenient thresholds were chosen to allow for p-value thresholding at a later step. Clumping radius was chosen to be 500 kb and the clump $r^2$ was 0.1. The output summary files of Plink clumping were converted to Hail Tables. The Tables were joined into separate MatrixTables, depending on the LD reference panel used for the clumping, using Hail's multi_way_zip_join method (wrapped by mwzj_hts_by_tree in https://github.com/atgu/ukbb_pan_ancestry/blob/master/plink_clump_hail.py). These MatrixTables were then merged together by column (hl.union_cols()).

### Meta-analysis

Fixed-effect inverse-variance weighted meta-analyses were conducted for all ancestry-phenotype pairs using a custom Hail script (https://github.com/atgu/ukbb_pan_ancestry/blob/master/run_meta_analysis.py). Briefly, meta-analyzed $\beta_{meta}$ and $SE_{meta}$ were computed as follows:

$$\beta_{meta} = \frac{\sum_i \left[\beta_i / SE_i^2\right]}{\sum_i \left[1 / SE_i^2\right]} \text{ and } SE_{meta} = \sqrt{\frac{1}{\sum_i \left[1 / SE_i^2\right]}},$$

where $\beta_i$ and $SE_i$ were obtained from each ancestry-specific GWAS[31].

In addition to the meta-analyses across all the available ancestries for each phenotype, we also conducted all combinations of leave-one-ancestry-out meta-analyses. We repeated this process for only 1,091 high-quality ancestry-trait pairs spanning 452 phenotypes (see Heritability analysis). Finally, we computed Cochran's Q, a measure of heterogeneity, and the associated p-value for each meta-analysis statistic.

While previous studies have adopted a random effect meta-analysis to account for effect size heterogeneity across ancestries, we adopted a fixed effect meta-analysis here given that 1) current random effect meta-analysis approaches do not scale well for thousands of traits, and 2) recent studies have shown high cross-ancestry genetic correlations of causal effects for common variants[32,33].

### Polygenicity

We used the SBayesS [34] method implemented in the GCTB software (available at https://cnsgenomics.com/software/gctb) to calculate polygenicity. As with summary statistic based heritability estimates, polygenicity estimates from relatively small sample sizes are unstable. Thus, our analysis was focused on GWAS summary statistics derived from EUR for 451 phenotypes (all high quality phenotypes, except height which failed to converge; **Supplementary Dataset 6**). We used the LD reference panel provided by the software, which was built upon 50,000 unrelated European samples from the UK Biobank. For the Markov Chain Monte Carlo process, we employed 4 chains to compute the Gelman-Rubin convergence diagnostic, also known as potential scale reduction factor, for estimation of polygenicity (the proportion of SNPs with nonzero effects), and the default parameters otherwise. Considering potential convergence challenges in Bayesian models, we set a threshold value of less than 1.2 for the Gelman-Rubin convergence diagnostic to signify satisfactory convergence of the estimated polygenicity, which resulted in 392 phenotypes (**Supplementary Fig. 31**).

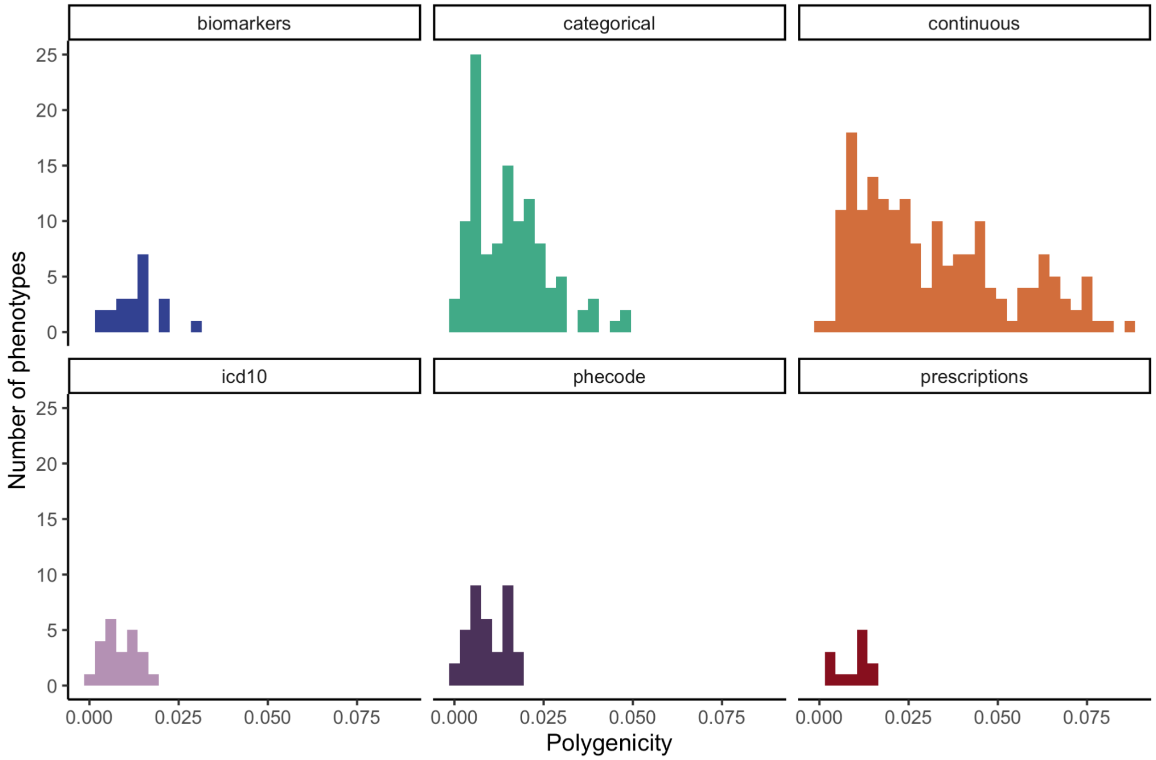


**Supplementary Figure 31** | Polygenicity estimates across trait types. A histogram of polygenicity estimates (the proportion of SNPs with nonzero effects) using SBayesS for 392 phenotypes in EUR.

### Summary statistics analysis

All analyses of summary statistics were performed using custom scripts written in Hail (version 0.2.59), available at https://github.com/atgu/ukbb_pan_ancestry. Briefly, custom aggregators were developed to compute a number of statistics, including top p-value by variant, the number of significant variants per trait per population, a comparison between EUR and meta-analysis summary statistics, and the correlation of effect sizes across populations, and a windowed analysis to assess the degree of overlap with previous associations.

#### Consistency in summary statistics

We investigated the extent to which data from multiple ancestries is consistent using two approaches. First, we compared the computed heterogeneity measures (Cochran's Q) for variants significant by meta-analysis to those significant in EUR-only GWAS. As expected, we find that variants significant in the meta-analysis only are less likely (1.5%) to be heterogeneous (Cochran's Q $p < 0.01$), compared to those variants discovered in both EUR and meta-analysis (6.8%), with those discovered only in the EUR subset most heterogeneous (17.1%; Extended Data Fig. 4). Accordingly, the most heterogeneous variants are skewed towards highly significant variants in EUR (**Supplementary Fig. 32**a). Further, for each trait, for each pair of ancestry groups with at least 20 associations at $p < 0.05$, we computed the correlation of beta vectors: for those vectors of betas that are significantly correlated, we find that the vast majority have a positive correlation, indicating the consistency of direction of effect sizes across ancestry groups (**Supplementary Fig. 32**b).

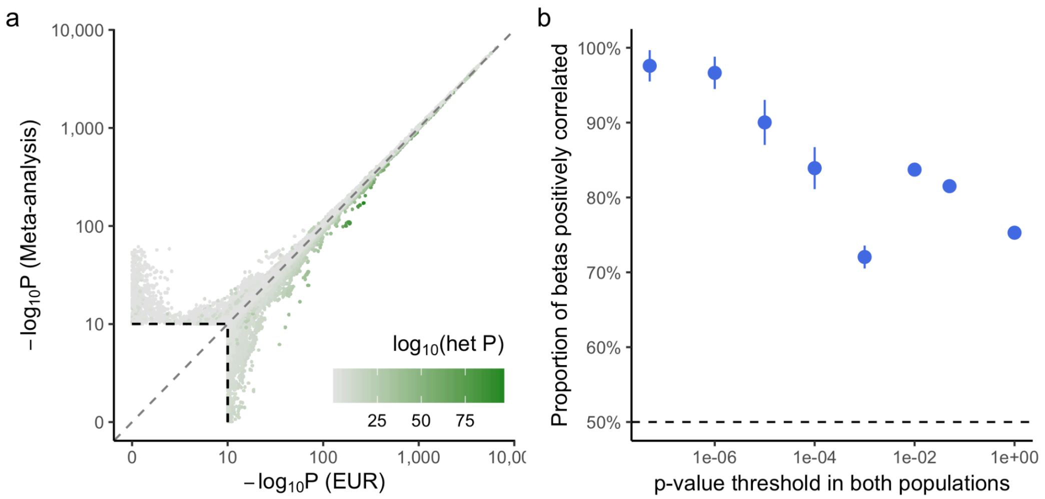


**Supplementary Figure 32 | Consistency of effects across ancestry groups.** (a) As in Figure 3c, P-values from meta-analysis versus EUR GWAS alone, colored by p-value of heterogeneity among genetic ancestry groups. (b) For associations that are significant in more than one ancestry group, the majority of betas are positively correlated.

#### Known versus novel association comparisons

We curated thousands of traits using a unified ontology, the Experimental Factor Ontology (EFO), which provides a systematic description of experimental variables with a hierarchical structure that spans a breadth of phenotypic domains including diseases, anthropometric traits, etc for projects such as the GWAS catalog[35,36]. To map as many traits in this project to EFO terms and categories as possible, we extended previous work from the OpenTargets OnToma package, which allows exact or fuzzy matching from a phenotype string. In total, we were able to map 3,047 (42%) of the traits with a GWAS to EFO terms. Of these traits, 2,566 (84%) were mapped to EFO categories defined by GWAS catalog. Our ability to map traits to EFO terms varied by trait type, with most disease, biomarker, and continuous traits mapping to EFO terms, whereas most and all of the categorical and prescription traits, respectively, did not map, including considerable questionnaire data (**Supplementary Table 10**). When defining novelty, we performed this analysis twice, first excluding two large similar previous efforts that also performed GWAS of all phenotypes in the UK Biobank, including the NEALE2[37] and UMich_SAIGE projects[17], to compare to GWAS prior to large-scale UK Biobank studies, and again with no exclusion.

Given differences in LD structure between genetic ancestry groups, we defined novelty of associations using distance-based windows, i.e. a variant is only considered to be novel if no known associations (for the same EFO term, or category, below) are present within 1 Mb, implemented in a custom Hail aggregator. Given the pervasive level of pleiotropy genome-wide as well as diagnostic and phenotypic imprecision, however, matching on exact EFO terms may overcount biological and pathway novelty. We therefore also evaluated novelty using EFO parent terms (denoted as trait categories here), which are more generic than EFO terms. For instance, body height (EFO_0004339) is an EFO term that maps to the body measurement EFO category (EFO_0004324) by GWAS catalog. We used the OpenTargets release 22.09 to retrieve known associations (gs://open-targets-genetics-releases/22.09/v2d), except for the NEALE2 which we instead used the original release[37] due to the lack of X chromosome associations and biomarker traits in the OpenTargets. To annotate EFO terms for UKB traits, we used a combination of previous mappings from the OpenTargets (gs://open-targets-genetics-releases/22.09/lut/study-index) and https://github.com/EBISPOT/EFO-UKB-mappings (a fixed non-corrupted tsv file is provided at https://github.com/atgu/ukbb_pan_ancestry/blob/master/opentargets/UK_Biobank_master_file.fixed.tsv), updated obsolete EFO terms to EFO release 3.49.0, and annotated biomarker EFO terms manually. Lastly, we added trait categories based on GWAS catalog mapping (https://www.ebi.ac.uk/gwas/api/search/downloads/trait_mappings). For the sake of consistency, we manually changed category annotations of 1) systolic blood pressure (EFO_0006335), diastolic blood pressure (EFO_0006336), and mean arterial pressure (EFO_0006340) from “Other measurement” to “Cardiovascular measurement” to be consistent with pulse pressure measurement (EFO_0005763), and 2) sitting height measurement (EFO_0011011) from “Other measurement” to "Body measurement" to be consistent with body height (EFO_0004339). The summary of EFO annotations is shown in **Supplementary Table 10**.

**Supplementary Table 10** | EFO annotation summary. The number of traits mapping to EFO terms and categories is shown by trait type. The final column indicates traits that map to multiple categories.

| Trait type | Num. EFO defined | Num. EFO missing | Num. EFO category defined | Num. EFO category (map to multiple) |
|---|---:|---:|---:|---:|
| biomarkers | 30 | 0 | 30 | 0 |
| categorical | 1,089 | 2,597 | 970 | 10 |
| continuous | 440 | 380 | 377 | 0 |
| icd10 | 662 | 259 | 565 | 6 |
| phecode | 826 | 500 | 624 | 3 |
| prescriptions | 0 | 445 | 0 | 0 |
| **Total** | **3,047** | **4,181** | **2,566** | **19** |

We identified 85,960 regionally-independent associations, of which 71,372 mapped to an EFO term/category. Of these, 36,708 significant associations (51%) have been reported previously for the exact same EFO term, while 34,664 (49%) did not. However, given the pervasive level of genome-wide pleiotropy as well as variability in diagnostic and phenotypic definitions, it is likely that matching on exact EFO terms may overestimate novelty. We therefore also computed an even more conservative measure of novelty using broad EFO categories and found that 66,123 (93%) were previously reported for the same EFO category and 5,249 (7%) are novel. Repeating this analysis without excluding previous UK Biobank efforts, we find 50,433 (70%)

associations previously reported for the same term, and 66,457 (93%) for the same category, resulting in 4,915 (7%) not mapping to the same category. Additionally, on the X chromosome, 2,226 of 2,448 (91%) associations were not found significant in Open Targets Genetics for the same EFO category; however, we note that the NEALE2[37] and UMich_SAIGE projects[17] did not have X chromosome results loaded into the OpenTargets release file, and thus, many of these associations are found in these resources as well.

#### Gene list analysis

We computed the top association for each variant using a custom Hail aggregator. Of 22,776,573 variants for which GWAS results were available (near 19,842 genes), 1,589,664 had at least one significant ($p < 5 \times 10^{-8}$) association, near 17,285 genes. Disease annotations for genes were obtained from https://github.com/macarthur-lab/gene_lists as previously described[24,38]. We filtered the 19,842 genes to 17,428 that are in the gene list "universe", of which 15,314 (87.8%) have a significant association. We repeated this analysis for each gene list, showing the fraction of genes in each gene list with at least one significant association, which is highest for haploinsufficient genes (180/187 = 96.3%; **Supplementary Fig. 33**), with a similar process for novel genes identified by EFO analysis (above; **Fig. 4b**).

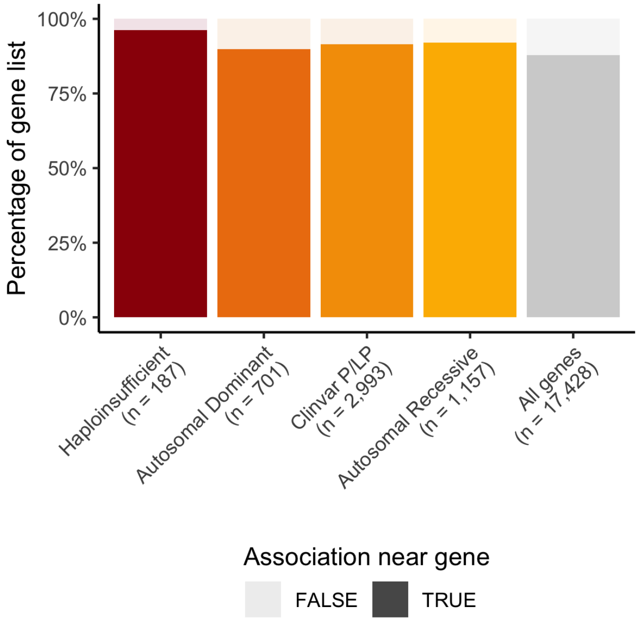


**Supplementary Figure 33** | Percentage of gene lists with at least one significant association. As in Fig. 4b, but all discovered associations rather than restricted to novel associations.

We generated LocusZoom plots for selected associations, including *PITX2* and *DMD*, using a modified version of https://github.com/krcurtis/locuszoom-plot.git. In these plots, linkage disequilibrium (LD) $r^2$ is computed as the sample-size weighted sum of LD from all the ancestry groups that are included in the meta-analysis: $\frac{\sum_{pop \in POP_{meta}} LD_{pop} \times N_{pop}}{\sum_{pop \in POP_{meta}} N_{pop}}$.

#### Ancestry-enriched associations

We manually investigated variants enriched for associations in the multi-ancestry meta-analysis compared to the EUR GWAS alone (top right of **Fig. 3c**). We identified a missense variant rs1050828 in *G6PD* associated with multiple phenotypes, which have low phenotypic and genetic correlations (**Fig. 4a**), indicating potential pleiotropy at this locus. We show the high-confidence effect sizes in a forest plot (**Fig 5c**), and all available summary statistics in **Supplementary Fig. 34** and **Supplementary Table 11**. Some of these phenotypes failed QC for some ancestry groups (e.g. HbA1c for AFR); however, we note that broad estimates of quality based on heritability may not be applicable to individual loci, such as the *G6PD* locus here.

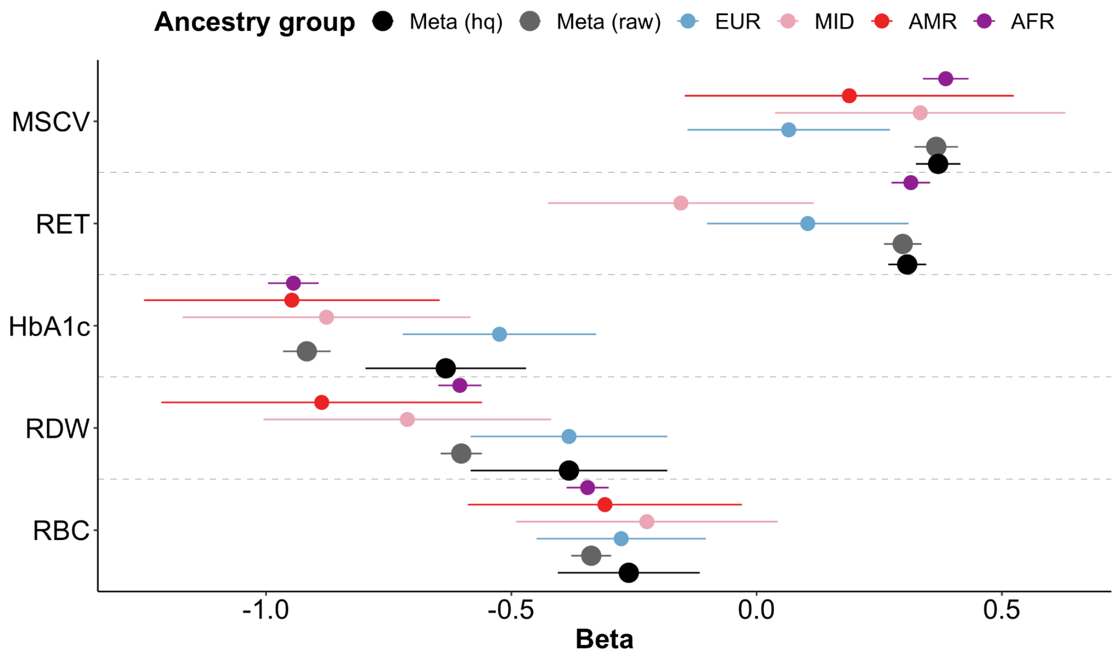


**Supplementary Figure 34** | Forest plot showing association beta for each phenotype for rs1050828 across all available population groups. Error bars correspond to 95% confidence intervals. Abbreviations are defined in **Supplementary Table 11**.

**Supplementary Table 11 | Top 5 phenotypes associated with SNP rs1050828 at gene G6PD.** * indicates associations passing GWAS significance threshold 5 x $10^{-8}$. This variant is low frequency in CSA and EAS and thus, GWAS was not run in these groups.

| Phenocode | Phenotype | Population | AF | Pvalue | QC flag failed |
|---|---|---|---|---|---|
| 30270 | Mean sphered cell volume (MSCV) | AFR | 1.74e-01 | 1.82e-59* | |
| | | $Meta_{raw}$ | 2.82e-03 | 1.04e-58* | |
| | | $Meta_{hq}$ | 2.75e-03 | 1.25e-57* | |
| | | MID | 1.19e-02 | 2.70e-02 | significant_z |
| | | AMR | 1.81e-02 | 2.70e-01 | significant_z<br>ancestry_resonable_n |
| | | EUR | 1.33e-04 | 5.34e-01 | |
| 30300 | High light scatter reticulocyte count (RET) | AFR | 1.74e-01 | 3.05e-55* | |
| | | $Meta_{hq}$ | 2.75e-03 | 1.30e-54* | |
| | | $Meta_{raw}$ | 2.78e-03 | 1.67e-52* | |
| | | MID | 1.19e-02 | 2.64e-01 | significant _z |
| | | EUR | 1.33e-04 | 3.20e-01 | |
| 30750 | Glycated haemoglobin (HbA1c) | $Meta_{raw}$ | 2.45e-03 | 1.09e-299* | |
| | | AFR | 1.73e-01 | 1.17e-281 | significant_z |
| | | $Meta_{hq}$ | 1.76e-04 | 3.13e-14* | |
| | | AMR | 1.90e-02 | 7.22e-10 | significant_z<br>ancestry_resonable_n |
| | | MID | 1.22e-02 | 4.76e-09* | |
| | | EUR | 1.31e-04 | 1.84e-07 | |
| 30070 | Red blood cell (erythrocyte) distribution width (RDW) | $Meta_{raw}$ | 2.84e-03 | 1.41e-173* | |
| | | AFR | 1.74e-01 | 3.18e-161 | significant_z |
| | | AMR | 1.82e-02 | 1.06e-07 | significant_z<br>ancestry_resonable_n |
| | | MID | 1.17e-02 | 1.95e-06 | significant_z |
| | | $Meta_{hq}$/EUR | 1.31e-04 | 1.84e-04 | |
| 30010 | Red blood cell (erythrocyte) count (RBC) | $Meta_{raw}$ | 2.84e-03 | 2.20e-59* | |
| | | AFR | 1.74e-01 | 6.54e-56 | significant_z<br>ancestry_resonable_n |
| | | $Meta_{hq}$ | 1.75e-04 | 4.19e-04 | |
| | | EUR | 1.31e-04 | 1.70e-03 | |
| | | AMR | 1.82e-02 | 3.00e-02 | significant_z<br>ancestry_resonable_n |
| | | MID | 1.17e-02 | 9.98e-02 | |

#### Fine-mapping

To identify putative causal variants, we conducted statistical fine-mapping using FINEMAP-inf and SuSiE-inf, which model infinitesimal effects[39]. We used ancestry-specific GWAS or cross-ancestry meta-analysis summary statistics, and a covariate-adjusted (see LD matrices and scores) in-sample dosage LD matrix, which we recomputed here using the exact samples included in each summary statistics. We defined fine-mapping regions based on a 3 Mb window around each lead variant and merged regions if they overlapped, as described previously[40]. For each method, we allowed up to ten causal variants per region and derived the posterior inclusion probability (PIP) of each variant using a uniform prior probability of causality. To achieve better calibration, we computed the minimum PIP across the methods and derived up to 10 independent 95% CSs from SuSiE-inf.

#### Comparison between Tractor and SAIGE results

We compared the Tractor analysis described above to our SAIGE association analysis. In this section, SAIGE AFR denotes the SAIGE analysis performed on the AFR ancestry groups. Tractor AFR and Tractor EUR indicate the Tractor GWAS conducted on the African or European haplotype tracts, respectively, within individuals from the AFR group. Tractor effectively controlled type 1 error within admixed AFR-like samples (**Extended Data Figure 8**); $\lambda_{GC}$ was better controlled in the Tractor AFR (1.0011) compared to SAIGE AFR (1.0031). As expected, the EUR tracts within admixed AFR-like samples were underpowered given that our study population included individuals with a mean AFR global proportion of 92.69%, which reduces the effective sample size of EUR ancestry to the relative proportion of the census size. Consequently, Tractor EUR (EUR tracts among AFR-like individuals) had a $\lambda_{GC}$ of 0.9609 and no genome-wide significant hits.

We observed a significant block of hits on chromosome 16 identified by both SAIGE AFR and Tractor AFR, with similar p-values (**Supplementary Dataset 7**; 'block 1'). Chromosome 11 harbored two separate loci passing genome-wide significance; one passing the threshold in SAIGE AFR and one in Tractor AFR (blocks 2 and 3, respectively). The Tractor-identified chr11 locus has divergent ancestry-specific allele frequencies at top loci, with an AFR-specific MAF of 1.6-1.8%, while the EUR-tracts were monomorphic. This MAF difference likely drove the improved ability of Tractor to identify this locus, as it is best-powered to identify ancestry enriched loci. For the four identified hits, we found largely similar effect sizes estimated from Tractor AFR and SAIGE AFR, while both AFR estimates varied considerably from EUR effect size estimates, which were not significant (**Supplementary Dataset 7**). Comparing the allele frequencies of variants from SAIGE AFR to those in Tractor AFR for shared variants, we observed a Pearson correlation of 0.8. Previous work shows that Tractor accurately estimates group-level effects, while traditional GWAS without local ancestry deconvolution produces effect sizes that reflect a weighted average of the group-level effects.

### Supplementary Datasets

**Supplementary Dataset 1 | Assigned genetic ancestry labels correlate with the country of birth or known migration events.** The number of individuals by genetic ancestry and country of birth (non-UK) are shown.

**Supplementary Dataset 2 | Summary of all phenotypes in Pan-UKB.** Phenotypes are keyed by five keys: trait type, phenocode, pheno_sex, coding, and modifier. Where available, description and coding_description are provided from the UK Biobank showcase. For each ancestry group, we include the number of cases, heritability estimates (observed, liability, standard errors, and z scores), whether the phenotype passes QC, and lambda GC. We provide QC flags, whether the phenotype is in the maximal independent set, and filename information, including a download link for the phenotype-specific file and tabix index on Amazon S3 and md5 checksums for each.

**Supplementary Dataset 3 | Summary of all heritability metrics.** Phenotypes are keyed as in Supplementary Dataset 2. For each ancestry group, we provide heritability estimates (observed, liability, standard errors, and z scores) for LDSC and S-LDSC, and for ancestry groups other than EUR, also RHE-mc, as well as details of QC flags.

**Supplementary Dataset 4 | Pairwise genetic correlations**. Genetic correlations (*rg*) from S-LDSC are computed for pairs of 528 phenotypes (*phenotype_code_1* and *phenotype_code_2*), using summary statistics from EUR.

**Supplementary Dataset 5 | Pairwise phenotypic correlations.** Covariates were regressed out from each of the 452 high-quality phenotypes, and pairwise correlations (*entry*) were computed for each pair of phenotypes (residuals), *i* (with phenotype identifier in *i_data*) and *j* (identifier in *j_data*). The correlation for all phenotypes is available at gs://ukb-diverse-pops-public/misc/pairwise/pairwise_correlations_regressed.txt.bgz

**Supplementary Dataset 6 | Polygenicity estimates.** Polygenicity estimates (mean and standard deviation) from SBayesS for 451 phenotypes, along with convergence criteria (R_GelmanRubin).

**Supplementary Dataset 7 | Summary statistics for key loci across GWAS methods.** SAIGE AFR and SAIGE EUR refer to the SAIGE analyses performed on the African (AFR) and European (EUR) genetically inferred ancestry groups of UKB. Tractor AFR and Tractor EUR indicate the Tractor GWAS conducted on the African or European haplotype tracts, respectively, within the AFR group. Variants are filtered as described above in Tractor GWAS analysis.

### FAQ for diverse ancestry GWAS

*Executive Summary:*
*The goal of this project is to provide a resource to researchers that promotes more inclusive research practices, accelerates scientific discoveries, and improves the health of all people equitably. In genetics research, it is statistically necessary to study groups of individuals together with similar ancestries. In practice, this has meant that most previous research has excluded individuals with non-European ancestries. Here, we describe an effort to build a resource using one of the most widely accessed sources of genetic data, the UK Biobank, in a manner that is more inclusive than most previous efforts -- namely studying groups of individuals with diverse ancestries. The results of this research have a number of important limitations which should be carefully considered when researchers use this resource in their work and when they and others interpret subsequent findings.*

This FAQ is intended to provide context around and describe some of the limitations of our analyses to a lay audience. It does not go into comprehensive detail around every phenotype, but instead highlights the overarching goals of the project, including how analyses were conducted, and describes some potential confounders that could affect our results. The practice of producing these public-facing FAQ documents was first adopted by the Social Science Genetics Association Consortium (https://www.thessgac.org/faqs) and has become a common practice among several genomics researchers. In this FAQ, we have followed a similar structure to that of others that have been previously released.

**Table of Contents**

FAQ for diverse ancestry GWAS
- Background
  - Who conducted this study?
  - What are the group's overarching goals?
  - Why was this study done?
  - What is ancestry? Is it the same as race or ethnicity?
  - In this study, you perform many GWAS for many phenotypes. What is a GWAS? What is a phenotype?
  - What does it mean for a variant to be associated with a phenotype? Are the genetic variants discovered by GWAS "causal"?
  - Do these results imply that genetics are responsible for the phenotypic differences between ancestry groups?
  - Since biology is mostly shared, why is diversity in genetics so important?
  - What did you learn as part of this study?
- Study design
  - What was done?

What data were used?
Have you used data from countries other than the UK?
How were participants recruited?
How did you decide which phenotypes to study?
How did you decide what ancestry groups to include? How did you assign individuals to each ancestry group?
What about people with mixed ancestries?
Why do you analyze ancestry groups separately?
Why have certain individuals been excluded in previous research?
Why are you including them now?
Social and Ethical Implications
What can be done with the results of this research? What are the potential benefits of this research?
Do you study the genetics of behavior?
Do genes determine the choices we make?
Are there policy or clinical implications for this research?
How has genetics research been used in the past to harm different groups?
Could this research be used to harm certain groups (e.g., through discrimination or stigmatization)?
What has been done to reduce the potential harms of this research?
References

### Background

#### Who conducted this study?

A team of researchers from the Analytic & Translational Genetics Unit (ATGU) at Massachusetts General Hospital and the Broad Institute of MIT and Harvard performed the analysis in this study.

The data used in these analyses are from the UK Biobank, a large-scale open database with hundreds of thousands of individuals' genotype data paired to electronic health records and survey measures. The UK Biobank recruited 500,000 people aged between 40-69 years in 2006-2010 from across the country to take part in this project. They have undergone measures, provided blood, urine and saliva samples for future analysis, detailed information about themselves, and agreed to have their health followed. The researchers at ATGU and the Broad Institute were not involved in the design of the UK Biobank resource or recruitment of participants, but have analyzed the breadth of this powerful resource.

Throughout this work, we regularly sought engagement and feedback from researchers and communities to help direct and contextualize this research and to discuss actions that will allow the substantial benefits of our analyses to outweigh the risks. Please see the section "What has been done to reduce the potential harms of this research?" to learn more about the specific individuals and groups who have been a part of this effort and for information on what we have done to reduce risks to disadvantaged groups.

#### What are the group's overarching goals?

Our fundamental goal was to build a resource that facilitates access to genetic association results (also known as summary statistics) for as many phenotypes in as many diverse populations as possible, particularly those that have traditionally been underrepresented in prior genetics work and excluded in most analyses of the widely-used UK Biobank resource.

We believe that easy access to high-quality data on diverse populations will accelerate research that can improve the health of the global population and can contribute to closing disparities that exist in the world. These association results can be used to better understand the biology underlying a broad range of traits. The additional specific benefits of including underrepresented populations in this study are described in detail in the section "What can be done with the results of this research? What are the potential benefits of this research?" and throughout this document. Substantial health and social disparities exist between the groups studied in this research. While these disparities are largely a direct result of environmental factors, we hope that our research will lead to further work that mitigates disparities, even though we do not directly study those disparities as part of this project.

#### Why was this study done?

This project is an effort to increase diversity in genetics research and to make use of data that is traditionally left out of analysis.

Historically in genetics research, for technical and social reasons described below, most prior work has been done only in populations with European ancestries. Data from participants with predominantly non-European ancestries were usually excluded from previous studies including many studies of the UK

Biobank. (See "Why have certain individuals been excluded in previous research?") Because of this, results from these genetic studies may not apply as well to other groups. This further implies that applying these findings to a clinical setting has the potential to increase, rather than decrease, health disparities.

Additionally, genetic studies require a large amount of expertise and computing power. By making these results publicly available, we remove this barrier to researchers and provide results for many phenotypes while using a consistent analysis pipeline. We hope this will accelerate the pace and reliability of genetic discoveries and encourage future studies to include data from more diverse participants.

#### What is ancestry? Is it the same as race or ethnicity?

The "ancestry" of a group of people is related to the set of ancestors from whom they inherited their genetic variants. It does not have natural boundaries and it is not the same as race or ethnicity.

The distinctions between a person's race and ancestry are important. "Ancestry" is a statistical construct based on the genetic variants that an individual inherited from their ancestors. "Race" and "ethnicity" are social constructs and group people based on perceived shared physical, geographical, cultural, language, religion, or other social characteristics, often in an inherently unequal manner. As a result, a person's race and ethnicity can depend on the time and place that an individual lives. Similarly, an individual's self-identified race or ethnicity may at times differ from the corresponding genetic ancestry assigned by statistical algorithms. Treating ancestry, ethnicity, and race as equivalent concepts is incorrect. In all our analyses, we exclusively refer to genetic ancestry.

When measuring ancestry across individuals, geneticists use statistical tools and very large data to identify groups of people who are more genetically similar. Based on the region where people in a group live and what is known about human migration, populations with similar ancestries are often given geographic labels. For example, the vast majority of the individuals in the UK Biobank appear to have similar ancestries to other individuals who have grandparents native to countries in Northern Europe. For this reason, geneticists often refer to such individuals as having "European" ancestry. That said, ancestry is a continuum that does not have obvious boundaries. It is possible to divide a group of individuals into any number of "ancestry groups." In this research, we use rough continental categories, as described in more detail in "How did you decide what populations to include? How did you assign individuals to each ancestry group?"

#### In this study, you perform many GWAS for many phenotypes. What is a GWAS? What is a phenotype?

A Genome-Wide Association Study (GWAS) is a scan of millions of genetic variants in the human genome, looking for variants that are associated with a particular "phenotype".

A phenotype is a disease, outcome, or trait that can be measured and studied in a quantitative manner. Examples of phenotypes include height, self-reported smoking behavior, or whether a person has been diagnosed with type 2 diabetes.

To test whether a genetic variant is associated with the phenotype, we compare individuals that have a copy or copies of the variant to those who have none. If the difference is large enough that it is very unlikely to have occurred by chance, we say that the variant is "associated" with the phenotype.

#### What does it mean for a variant to be associated with a phenotype? Are the genetic variants discovered by GWAS "causal"?

Associations can occur for many reasons. Many of these reasons are not causal effects of the genetic variant on the phenotype.

For example, consider a case when the phenotype is a disease. It could be that the variant triggers a biological mechanism that directly leads to disease. This variant would be associated with the phenotype and would also be considered causal. On the other hand, it could be that the variant is by chance more common in certain groups or communities, and that they have a higher prevalence of disease due to environmental factors like pollution or social programs. This variant would also be associated with the phenotype but it would not be considered causal. As a third example, many variants that have significant associations in a GWAS are not expected to themselves be causal, but instead might be associated because they are nearby on the chromosome to a variant that is causal. Finally, in some cases, genetic variants may only be associated with the phenotype in certain settings. Generally, GWAS can only tell us if a genetic variant is associated with a phenotype; **it cannot tell us why the variant is associated and does not demonstrate that the relationship is causal**.

#### Do these results imply that genetics are responsible for the phenotypic differences between ancestry groups?

They do not. It is important to keep in mind that human populations are more genetically alike than we are different.

We don't know whether there is a genetic basis for differences between groups because GWAS variants aren't necessarily causal. Because of this, interpreting differences in genetic associations between groups is incredibly complicated, as associations can appear for a variety of reasons that don't imply causality. Please see our explanation "What does it mean for a variant to be associated with a phenotype? Are the genetic variants discovered by GWAS "causal"?".

Additionally, for all of the phenotypes considered by this study, **there is not a single genetic variant that determines whether you will have a certain condition**. Instead, in most cases, there are many, many genetic variants that each have a small association with the phenotype, yielding on average very similar outcomes in different populations. Because these associations are so small, it takes substantial follow-up work and an enormous amount of data to determine whether a genetic variant has a population-specific effect -- often more data than are currently available to researchers.

This is further complicated by the fact that there are many other factors (unrelated to biological differences) that can lead to differences in associations between populations. Perhaps the most important of these reasons is that different populations often face different environmental factors (e.g., discrimination, rates of poverty, geography), and these factors may affect genetic associations. Together, this means that GWAS alone are insufficient to explain biological differences among populations.

#### Since biology is mostly shared, why is diversity in genetics so important?

Diversity in genetic studies is important to ensure that findings are generalizable and that everyone can benefit from these findings.

While previous genetic studies have provided deep insights into the molecular basis of many phenotypes, participants in those studies have mostly consisted of groups of people who trace their ancestry back to regions within Europe. While all people have more genetic similarities than differences, certain genetic variants or combinations of variants are more common in groups that have ancestries that trace back to close-by regions. As a result, most previous studies have been best-suited for understanding the role of genetic variants that are more common in people from those European regions. Expanding genetics research to include individuals with diverse ancestries will improve our understanding of these phenotypes for everyone. For example, more diversity will help researchers establish which genetic variants are actually causal and which are just simply associated. It will also help researchers discover new biological mechanisms since some genetic variants are only common enough to study in certain populations. Discovering these biological mechanisms will help us better understand the underlying biology of important phenotypes shared by everyone. Additionally, studying underrepresented groups may make precision medicine possible for these currently underserved populations themselves.

#### What did you learn as part of this study?

Our results from this study provide a starting point for understanding the genetic underpinnings of thousands of traits and diseases across global populations.

In this work, we identified thousands of links between genetics and human diseases in traits, many of which had not been seen before due to the Euro-centric nature of previous studies. For example, we find a genetic variant that influences triglyceride levels and is commonly found in individuals with African ancestry but very rare in individuals with European ancestry. Other variants in this gene have previously been implicated in heart failure. Furthermore, we developed computational resources and an approach for future studies performing similar analyses to maximize future disease gene discovery.

We released these results well in advance of scientific publication or submission timelines because of the disproportionate value of these results to the field. For example, as part of our study, we released GWAS results for a set of phenotypes related to COVID-19. While it is well-understood that differential rates of transmission of COVID-19 are primarily social and environmental, GWAS results may improve our understanding of the biological mechanisms that influence disease severity and accelerate the discovery of an urgently-needed treatment. By facilitating access to analyses conducted in underrepresented populations, we hope that the broader scientific community makes use of these data such that data from diverse ancestry participants in other cohorts are not discarded. As this study moves forward, we and others in the research community will continue to produce important scientific insights.

### Study design

#### What was done?

We used genetic and phenotypic data from ~500,000 participants in the UK Biobank to conduct genome-wide association studies (GWAS) among individuals with diverse ancestral backgrounds for several thousand phenotypes. This included more than 20,000 individuals with primarily non-European ancestries.

The UK Biobank is an open access database with hundreds of thousands of individuals' genetic data paired to electronic health records and survey measures. We conducted GWAS for all phenotypes deemed to have sufficient statistical power. These phenotypes include a total of more than 16,000 GWAS conducted across a very broad range of phenotypes. A few examples of phenotypes we studied include anthropometric measures and physical attributes (e.g. height, BMI, bone density), blood panel traits (e.g. white blood cell count, cholesterol, blood glucose), common diseases (e.g. diabetes, cardiovascular disease, psychiatric disorders), electronic health record data (e.g. diagnosis codes entered by clinicians), prescription data (e.g. prescribed to take statins), health surveys (e.g. dietary intake, activity levels, general health satisfaction), social surveys (e.g. educational attainment, occupation), and many other measures. To summarize, phenotypes included both data pulled from electronic medical records as well as participants' survey responses to questionnaires given online or at the clinic.

#### What data were used?

The data used in these analyses comes from the UK Biobank, a large-scale open database with hundreds of thousands of individuals' genotype data paired to electronic health records and survey data.

Researchers can gain access to the UK Biobank by writing a proposal for a research project, which then is reviewed for approval. The UK biobank is more thoroughly described on their website and in a scientific publication.

#### Have you used data from countries other than the UK?

No, all participants lived in the UK at the time that the data were collected.

Because all of our data come from the UK Biobank and we did not collect additional data as part of this project, all individuals must have lived in the UK during the UK Biobank recruitment phase. (See "How were participants recruited?") However, the data do include individuals who were born outside of the UK but were recruited to be part of the UK Biobank sample after having moved to the UK. Since there are important differences in genetic diversity between the UK, the US, and other countries, we hope that resources comparable to this one will be produced as large new datasets become available in the future.

#### How were participants recruited?

Our team did not recruit participants for this study. Instead, we analyzed existing data from the UK Biobank, a collection of ~500,000 individuals collected in the United Kingdom about ten years ago.

Here, we describe how participants in the UK Biobank cohort were recruited. Following the success of an initial pilot study in 2005-2006, the main stage of recruitment for the UK Biobank resource began in 2007, with the goal of recruiting 500,000 individuals between the ages of 40 and 69. The age restriction was due to the primary aim of the study: to improve the prevention, diagnosis and treatment of serious illnesses that typically onset later in life, including diabetes, cancer, arthritis, heart disease, stroke, and dementia. To that end, individuals from across the UK were contacted by post to participate in the study, with names, addresses, and dates of birth provided by the UK National Health Service (NHS). The 500,000 recruitment goal was reached in July of 2010, and recruitment ended shortly after. Focusing on voluntary recruitment of an older subset of the UK population sent by mail resulted in a sample of individuals that is more healthy and wealthy than the average Brit and that **had a greater fraction of European ancestries than the UK population overall**. This means that this cohort is not a perfect representation of the UK population, which further means that the results of our study may not reflect the UK population as a whole. This limitation is important to keep in mind when considering our results.

#### How did you decide which phenotypes to study?

We included all phenotypes that were available to us for which there was sufficient data to conduct a GWAS.

Due to the scale of this project, we relied on quality control procedures that worked well in general for all phenotypes rather than using specific quality controls for each one. Quality controls are the procedures used to minimize errors in the data we use in our analysis. For example, individuals who report extremely large or small values of a phenotype are often just incorrectly recorded and can bias analyses. Therefore, we adjusted the values for such individuals using a standard transformation. For complete details on our quality control procedures, see our wiki. Certain phenotypes may require different quality control procedures than those we used in our analyses. As a result, some researchers may prefer to include different sets of individuals or define phenotypes slightly differently in their future work.

#### How did you decide what ancestry groups to include? How did you assign individuals to each ancestry group?

The ancestry groups we used in these analyses are based on those described in two large existing globally and genetically diverse datasets. To assign each person to each ancestry group, we applied statistical methods to each individual's genetic data.

Specifically, we compared the genome of each participant in the UK Biobank to the data in two large reference datasets containing genetically diverse individuals from across the globe, the 1000 Genomes Project and Human Genome Diversity Project (HGDP). We statistically assessed how similar each participant's ancestry was to individuals from the populations included in these reference panels. These previous studies included labels which break down populations broadly into continental groupings that share ancestors and history over the course of tens to thousands of generations. The ancestry labels include African, American (which in these studies is the ancestry shared by many Hispanic/Latino groups), Central/South Asian, East Asian, European, and Middle Eastern. We assigned each individual into the ancestry groups that he/she was most similar to, adopting the same labels as used previously for consistency. We dropped those individuals who did not have a confident ancestry assignment. Notably, this approach does not rely on any other information, including self-reported race, ethnicity, or ancestry. We conducted our studies in all of the populations that had large enough numbers of individuals to learn about the genetic underpinnings of some traits, with individuals from each population analyzed together.

These ancestry labels have many limitations. First, as described in response to the question “What is ancestry?”, there is substantial diversity within each labeled population. Second, the 1000 Genome Project and HGDP data used to define continental populations have gaps in representation. They include more individuals from some regions than others, but this is not necessarily reflective of a region’s corresponding genetic diversity. For example, the individuals in the “African” population in the 1000 Genomes Project data have more West African ancestors than ancestors from other parts of Africa. For this reason, groups with ancestries from other parts of Africa may not be identified as accurately. Third, many individuals have ancestors from more than one of the groups defined by the 1000 Genome Project and HGDP. Such individuals are said to be “admixed,” which means that different parts of their genomes come from different continental populations. We discuss how such individuals affect our analysis more in the question below in “What about people with mixed ancestries?”. We hope that in the future, more diverse reference data sets will become available so that analyses such as ours will be less susceptible to these limitations.

#### What about people with mixed ancestries?

Many admixed individuals are included in this study.

Since ancestry is a continuum (see “What is ancestry?”), some ancestry labels consist nearly exclusively of individuals with varying amounts of admixture. For example, most individuals that are labeled as having “American” or “African” ancestries by our statistical algorithms share recent common ancestors with those labeled as having “European” ancestries. As long as there are enough people with a similar pattern of admixture -- as there are in the “African” and “American” ancestry groups -- we can study them together in a GWAS. However, some individuals have less common patterns of admixture, such that there are not enough similar individuals that we can group them together in a genetic study. Therefore, we currently drop such individuals from analysis. However, we believe that including these individuals should also be a priority, and plan to implement tools that allow us to include them in ongoing and future work to allow for increased inclusion of admixed participants in future studies.

#### Why do you analyze ancestry groups separately?

Different ancestry groups are analyzed separately for statistical reasons; this does not imply that there are biological differences between ancestry groups (see “Do these results imply that genetics are responsible for the phenotypic differences between ancestry groups?”).

To help understand why previous scientific efforts have restricted to only one ancestry group, a classic GWAS example is of chopstick use. If we conducted such a GWAS in a group with some people with East Asian ancestries and some people with non-East Asian ancestries, we would almost surely find many associations. These associations would not likely correspond to biological mechanisms that affect manual dexterity or a personal preference for wooden cutlery, but they would just identify genetic variants that are by chance more or less common in East Asian populations relative to the ancestries represented by others in the data. When GWAS is conducted in groups of individuals with similar ancestries, associations are less likely to be driven by these types of non-causal factors, which makes the results easier to interpret and makes follow-up work more productive.

#### Why have certain individuals been excluded in previous research?

Due to the statistical reasons described above, there are some scientific advantages to studying large groups of genetically similar individuals. Unfortunately, due to a number of historical, infrastructural, political, ideological, monetary and other societal factors, this has resulted in the disproportionate recruitment of individuals with European ancestries, effectively excluding other groups from participating in most genetic studies.

Previous work[41] described some of the more societal explanations for why data collection has heavily focused on European populations. For statistical reasons (see Why do you analyze ancestry groups separately?), standard practice in genetic studies has been to only analyze the largest homogenous subset of the data, which in practice means only including individuals who are of the largest ancestry group. Due to this Euro-centric bias in data collection, the largest group usually consists of those with European ancestry. This snowballing effect has driven the overrepresentation of European-ancestry individuals in published GWAS, and perpetuates the continued data collection and study of European-ancestry individuals because they have been more fully characterized.

A description of how ancestry is classified and where ancestry labels come from is in the section on "How did you decide what populations to include? How did you assign individuals to each ancestry group?".

#### Why are you including them now?

The UK Biobank provides a unique opportunity to study people with diverse ancestries. Although people with recent ancestors from outside the UK make up a small fraction of the data in the biobank, there are enough to run and learn new biology from GWAS. Many of these genotypic and phenotypic datasets are the largest available that include individuals with certain ancestries.

Because associations between single genetic variants and phenotypes tend to be very small, a large number of individuals need to be studied to find reliable associations. That is, we often need at least tens of thousands of individuals (and often more) to find and validate genetic associations. With this in mind, the groups of individuals that have been omitted from previous genetic studies were far too small to be able to produce reliable results. **Analyzing these data and releasing the results to the research community and the public will hopefully accelerate research that will benefit global populations.**

#### Social and Ethical Implications

##### What can be done with the results of this research? What are the potential benefits of this research?

Including more diverse populations in gene discovery efforts benefits all individuals, and may be especially beneficial for underserved populations.

Perhaps the most notable example of this is in the improvements to ‘fine-mapping.’ Fine-mapping is using statistics to attempt to identify which variant is responsible for an observed association in a GWAS. Because we inherit entire segments of chromosomes together, we jointly inherit all the variants contained on those segments. As a result, associations that are identified by GWAS may correspond to the effect of a variant on another part of the same chromosome. Due to differences between ancestry groups in the size and location of these segments, studying different groups allows us to narrow the search for the truly associated variant to a smaller number of genetic variants. Since there are fewer variants that need to be examined in laboratory studies, fine-mapping accelerates the process of linking genetic associations to potential biological pathways and the development of potential therapies. One example of leveraging multi-ethnic populations to better determine causal gene sets is in a publication on inflammatory bowel disease[42]. By using a combination of European and East Asian cohorts, researchers were able to refine their GWAS signals to single variants with high certainty. This allowed them to link inflammatory bowel disease to specific immune cells and gut mucosa. Including diverse populations thus allows improved determination of strong genetic determinants and pathways for disease that can be targeted therapeutically, both for the minority populations themselves, but also for people of all ancestries.

Another benefit from including diverse populations in GWAS scans is that we have the potential to identify variants that are unique to different populations. Some genetic variants are only found in groups with certain ancestries. If a variant has an effect on a phenotype but is not found in samples with European ancestries, it would be impossible to discover the association between the variant and phenotype in people with only European ancestry. In contrast, studying diverse populations will discover these kinds of variants. Identifying more variants will help map out the biological causes of disease, which will benefit people of all ancestries.

Last but not least, GWAS have become large enough that polygenic scores are now very widely used in research settings and are even being considered for applications in clinical settings. However, given very large Eurocentric study biases in genetic studies, polygenic scores are currently far more accurate in populations with European ancestries. This means we can predict several traits and diseases in European ancestry populations rather well--similarly in fact to how accurately LDL cholesterol predicts heart disease. In contrast, these study biases mean polygenic scores are currently several-fold less accurate for example in East Asian and African ancestry populations, sometimes not much more predictive than a coin flip. One of our driving motivations to do genetic studies in more diverse participants and make these results widely accessible is to ensure that genetics can deliver on a mission to improve healthcare for all.

#### Do you study the genetics of behavior?

This study includes results for all phenotypes which had sufficient sample size to run a well-powered GWAS, including many behavioral phenotypes that were collected by the UK Biobank.

Several researchers, including some involved in this project, have also conducted GWAS of behavioral phenotypes in populations with only European ancestries. Behavioral research on these topics is particularly sensitive. We recommend that interested readers should also read the FAQs for those papers, which go into much greater detail on the interpretation as well as social and ethical implications of studying the genetics of behavior. Those FAQs may be found here: https://www.thessgac.org/faqs.

#### Do genes determine the choices we make?

Genes do not determine our choices or who we become.

If they did, identical twins would make all of the same decisions, have the same interests, etc. Years of twin studies have shown that, while identical twins tend to be more similar than fraternal twins, they also tend to be different in a lot of ways. This suggests that environmental factors--such as culture, institutions, and policy--also play a large role in our phenotypes. And even for highly heritable phenotypes where many genetic variants are strongly associated with the phenotype, the associations that are identified by GWAS may not represent causal mechanisms. Furthermore, even when associations represent causal relationships, these causal pathways are complicated and interact with the environment. For example, imagine that there was a major pandemic that caused countries to shut down the public school systems. In such a scenario, we might imagine that the genetic influences that are related to academic achievement in the pandemic regime may be different from the influences that are related to academic achievement for those in formal public schooling.

That said, despite the limitations in interpreting genetic associations with behavior, this research is still valuable. For example, socioeconomic status (SES) is among the most important risk factors for many diseases and health outcomes. This means that any genetic variant that is associated with a disease may just reflect the association between the disease and SES. By analyzing GWAS of SES-related phenotypes alongside GWAS of disease phenotypes, researchers can focus on genetic variants that are associated with disease but not SES. These variants are more likely to represent strong biological risk factors of disease that can be tested rigorously in follow-up research.

#### Are there policy or clinical implications for this research?

No. Making policy or clinical decisions based on the results of this study would be incredibly premature.

GWAS look for associations between genetic variants and phenotypes in the world as it is today. Policy and clinical questions ask what the world would look like if we did things differently. GWAS cannot answer those questions. We are hopeful that the results of this study will facilitate future policy and clinical research for the global population, but those who use our results to make broad policy claims are overinterpreting the results.

#### How has genetics research been used in the past to harm different groups?

Genetics unfortunately has a legacy of racist research that has harmed minority populations.

Indeed, the term "eugenics" was coined in the late 1800s by one of most prominent early researchers in heredity, Francis Galton, and perhaps the most influential geneticist and statistician in history, Ronald Fisher, was an active proponent of the belief that socio-economic disparities in society were primarily driven by genetic factors in the early 1900s. These racist attitudes among several in the scientific community laid the groundwork for the "forced sterilizations, anti-miscegenation laws, and immigration restrictions of the 20th century." These policies overwhelmingly targeted minorities and people of color.

In addition to racist policies in the name of science, a lack of community involvement in research also has led to harms to certain minority groups. For example, in the 1990s, members of the Havasupai tribe, a small Native American group based in the Grand Canyon, approached researchers at Arizona State University (ASU) asking if there was anything that could be done to treat the high rates of diabetes in their community. Blood samples taken from several tribe members, who were told that it would be used to "study the causes of behavioral/medical disorders." Over the subsequent years, these samples were used in a variety of research projects beyond diabetes research. Some of these studies were about the tribe's geographic ancestral origins, suggesting narratives that were in direct conflict with the tribe's traditional stories of its origins in the Grand Canyon. Some worried that this research could be used to threaten their sovereign rights to their land. It was clear that many of the individuals whose blood samples were used in that research would not have consented if they had understood the scope of the projects that would be done. Ultimately, the Havasupai tribe sued ASU. As part of the settlement for the lawsuit, ASU returned the samples, some of the research that had been done was withdrawn, and the tribe received a large payment and other concessions from ASU. This story highlights how carefully executed research on topics that may seem benign to researchers can harm disadvantaged groups and sow distrust between researchers and the community.

Acknowledging the harm done to certain groups in the past in the name of science reminds us of the importance of careful communication of the implication of scientific research and intense vigilance that disadvantaged groups are not further harmed by this and other related work.

#### Could this research be used to harm certain groups (e.g., through discrimination or stigmatization)?

Yes, but excluding these groups from genetics research is also harmful. For this reason it is important to be aware of and mitigate potential harms.

As described above, genetics has a long history of being used to stigmatize certain groups. Although our results do not imply that phenotypic differences between groups are due to genetic or biological differences, we do anticipate that some racist individuals may mistakenly or willfully misinterpret our study to advance their ideological agendas. However, the exclusion of diverse groups from genetics research directly harms minority populations. When research is based only on one group, subsequent treatments and policies that are tested and implemented will most greatly benefit that population, exacerbating disparities. Remember that human populations are more genetically alike than we are different. Including a broader and more inclusive set of individuals helps support biological understanding for all groups, and does not imply a meaningful difference between them.

We have carried out a number of activities in an effort to maximize benefits and minimize risks from this research. See our response in "What has been done to reduce the potential harms of this research?" for more information.

#### What has been done to reduce the potential harms of this research?

We have adopted several strategies in an effort to reduce the potential harms of this research.

First, we have written this FAQ so that interested laypeople and the media can understand the value and the limitations of this work. We will treat this as a living document and welcome feedback from the community if any aspect of our analyses or their interpretation remain unclear.

Second, we also discussed this project and this FAQ with groups and individuals with diverse expertise and perspectives. For example, we met on several occasions with members of Shades@Broad, an identity-based affinity group whose mission is to advocate for and support the recruitment, development, and success of ethnic minorities at the Broad. We sought their comments, advice, and perspectives throughout the analysis, interpretation, and dissemination of results. We also obtained feedback on our research and this FAQ from researchers and clinicians who work with diverse communities across the US.

Third, Shawneequa Callier, a bioethics professor who specializes in the ethical, legal, and social implications of genetics research and racial categories reviewed is an author on this manuscript and provided feedback on this FAQ. Professor Callier advised us on ethical considerations surrounding this research.

Even with these efforts, it is still likely that some will misinterpret this work. As such occasions arise, we will attempt to correct the public record with firmness where appropriate.

### References


1. Lander, E. S. & Schork, N. J. Genetic dissection of complex traits. *Science* **265**, 2037–2048 (1994).
2. 1000 Genomes Project Consortium *et al.* A global reference for human genetic variation. *Nature* **526**, 68–74 (2015).
3. Li, J. Z. *et al.* Worldwide human relationships inferred from genome-wide patterns of variation. *Science* **319**, 1100–1104 (2008).
4. Purcell, S. *et al.* PLINK: a tool set for whole-genome association and population-based linkage analyses. *Am. J. Hum. Genet.* **81**, 559–575 (2007).
5. Manichaikul, A. *et al.* Robust relationship inference in genome-wide association studies. *Bioinformatics* **26**, 2867–2873 (2010).
6. Bycroft, C. *et al.* The UK Biobank resource with deep phenotyping and genomic data. *Nature* **562**, 203–209 (2018).
7. Gurdasani, D. *et al.* The African Genome Variation Project shapes medical genetics in Africa. *Nature* **517**, 327–332 (2015).
8. Majara, L. *et al.* Low generalizability of polygenic scores in African populations due to genetic and environmental diversity. *Cold Spring Harbor Laboratory* 2021.01.12.426453 (2021) doi:10.1101/2021.01.12.426453.
9. Atkinson, E. G. *et al.* Tractor uses local ancestry to enable the inclusion of admixed individuals in GWAS and to boost power. *Nat. Genet.* (2021) doi:10.1038/s41588-020-00766-y.
10. Mathieson, I. & Scally, A. What is ancestry? *PLoS Genet.* **16**, e1008624 (2020).
11. Lewis, A. C. F. *et al.* Getting genetic ancestry right for science and society. *Science* **376**, 250–252 (2022).
12. BBC ON THIS DAY. *BBC*.
13. Karczewski, K. J. *et al.* Systematic single-variant and gene-based association testing of thousands of phenotypes in 394,841 UK Biobank exomes. *Cell Genomics* **2**, 100168 (2022).
14. Palmer, D. S. *et al.* Analysis of genetic dominance in the UK Biobank. *Science* **379**, 1341–1348 (2023).

15. Millard, L. A. C., Davies, N. M., Gaunt, T. R., Davey Smith, G. & Tilling, K. Software Application Profile: PHESANT: a tool for performing automated phenome scans in UK Biobank. *Int. J. Epidemiol.* **47**, 29–35 (2018).

16. Denny, J. C. *et al.* Systematic comparison of phenome-wide association study of electronic medical record data and genome-wide association study data. *Nat. Biotechnol.* **31**, 1102–1110 (2013).

17. Zhou, W. *et al.* Efficiently controlling for case-control imbalance and sample relatedness in large-scale genetic association studies. *Nat. Genet.* **50**, 1335–1341 (2018).

18. Koenig, Z. *et al.* A harmonized public resource of deeply sequenced diverse human genomes. *bioRxiv* (2023) doi:10.1101/2023.01.23.525248.

19. Van der Auwera, G. A. & O'Connor, B. D. *Genomics in the Cloud: Using Docker, GATK, and WDL in Terra*. (O'Reilly Media, 2020).

20. Hofmeister, R. J., Ribeiro, D. M., Rubinacci, S. & Delaneau, O. Accurate rare variant phasing of whole-genome and whole-exome sequencing data in the UK Biobank. *Nat. Genet.* **55**, 1243–1249 (2023).

21. Maples, B. K., Gravel, S., Kenny, E. E. & Bustamante, C. D. RFMix: a discriminative modeling approach for rapid and robust local-ancestry inference. *Am. J. Hum. Genet.* **93**, 278–288 (2013).

22. Alexander, D. H., Novembre, J. & Lange, K. Fast model-based estimation of ancestry in unrelated individuals. *Genome Res.* **19**, 1655–1664 (2009).

23. McCaw, Z. R., Lane, J. M., Saxena, R., Redline, S. & Lin, X. Operating characteristics of the rank-based inverse normal transformation for quantitative trait analysis in genome-wide association studies. *Biometrics* **76**, 1262–1272 (2020).

24. Karczewski, K. J. *et al.* The mutational constraint spectrum quantified from variation in 141,456 humans. *Nature* **581**, 434–443 (2020).

25. Yang, J. *et al.* Genetic variance estimation with imputed variants finds negligible missing heritability for human height and body mass index. *Nat. Genet.* **47**, 1114–1120 (2015).

26. Gazal, S. *et al.* Linkage disequilibrium-dependent architecture of human complex traits shows action of negative selection. *Nat. Genet.* **49**, 1421–1427 (2017).

27. Bulik-Sullivan, B. *et al.* An atlas of genetic correlations across human diseases and traits. *Nat. Genet.* **47**, 1236–1241 (2015).
28. Finucane, H. K. *et al.* Partitioning heritability by functional annotation using genome-wide association summary statistics. *Nat. Genet.* **47**, 1228–1235 (2015).
29. Evans, L. M. *et al.* Comparison of methods that use whole genome data to estimate the heritability and genetic architecture of complex traits. *Nat. Genet.* **50**, 737–745 (2018).
30. Pazokitoroudi, A. *et al.* Efficient variance components analysis across millions of genomes. *Nat. Commun.* **11**, 4020 (2020).
31. de Bakker, P. I. W. *et al.* Practical aspects of imputation-driven meta-analysis of genome-wide association studies. *Hum. Mol. Genet.* **17**, R122–8 (2008).
32. Shi, H. *et al.* Population-specific causal disease effect sizes in functionally important regions impacted by selection. *Nat. Commun.* **12**, 1098 (2021).
33. Hou, K. *et al.* Causal effects on complex traits are similar for common variants across segments of different continental ancestries within admixed individuals. *Nat. Genet.* **55**, 549–558 (2023).
34. Zeng, J. *et al.* Signatures of negative selection in the genetic architecture of human complex traits. *Nat. Genet.* **50**, 746–753 (2018).
35. Malone, J. *et al.* Modeling sample variables with an Experimental Factor Ontology. *Bioinformatics* **26**, 1112–1118 (2010).
36. Buniello, A. *et al.* The NHGRI-EBI GWAS Catalog of published genome-wide association studies, targeted arrays and summary statistics 2019. *Nucleic Acids Res.* **47**, D1005–D1012 (2019).
37. Howrigan, D. Details and Considerations of the UK Biobank GWAS. Preprint at (2017).
38. Lek, M. *et al.* Analysis of protein-coding genetic variation in 60,706 humans. *Nature* **536**, 285–291 (2016).
39. Cui, R. *et al.* Improving fine-mapping by modeling infinitesimal effects. *Nat. Genet.* 1–8 (2023).
40. Kanai, M. *et al.* Insights from complex trait fine-mapping across diverse populations. *bioRxiv* (2021) doi:10.1101/2021.09.03.21262975.
41. Martin, A. R., Teferra, S., Möller, M., Hoal, E. G. & Daly, M. J. The critical needs and challenges for genetic

architecture studies in Africa. *Curr. Opin. Genet. Dev.* **53**, 113–120 (2018).

42. Huang, H. *et al.* Fine-mapping inflammatory bowel disease loci to single-variant resolution. *Nature* **547**, 173–178 (2017).
